# Intervention Characteristics and Mechanisms and their Relationship with the Influence of Social Prescribing: a Systematic Review

**DOI:** 10.1101/2023.11.17.23298673

**Authors:** Eveline M. Dubbeldeman, Jessica C. Kiefte-de Jong, Frank H. Ardesch, Mirte Boelens, Laura A. van der Velde, Sophie G.L. van der Steen, Miriam L. Heijnders, Mathilde R. Crone

## Abstract

**Introduction:** Social Prescribing (SP) is an integrated care program aimed to improve individuals’ health and wellbeing. Understanding the influence of SP and determining best practices and processes is challenging due to variability in its delivery, implementation, and intervention characteristics between different studies and countries. This study aimed to identify the intervention characteristics, mechanisms, and outcomes associated with SP research, and explore how these factors relate to the influence of SP on health and wellbeing, healthcare utilization, and care experiences.

**Method:** A comprehensive search was conducted in 12 databases, Google Scholar, and reference lists of relevant studies published from January 2010 up to April 2023. Searches were limited to literature written in English or Dutch. The methodological quality of the included studies was assessed using the Mixed Methods Appraisal Tool and the risk of bias was evaluated using the Cochrane RoB2 and the ROBINS-I. We coded all intervention characteristics, mechanisms, and relevant outcomes. Qualitative data were visually presented using Harvest Plots and qualitative data were narratively summarized.

**Results and discussion:** In total, 49 papers were included, of which seven qualitative, seventeen quantitative, and 25 mixed method studies. Moreover, the findings highlights the importance of social-related mechanisms, including loneliness and social connectedness, in contributing to the observed positive influence of SP on mental health and wellbeing. The observed outcomes seem to be influenced by various characteristics, including gender, age, the presence of a link worker, and the use of behavior change techniques. However, we should be cautious when interpreting these results due to limitations in study designs, such as the lack of controlled trials and statistical considerations. Further rigorous research is needed to comprehensively understand the impact and potential benefits of SP.

## 1. Introduction

Social Prescribing (SP) is an integrated care program that enables healthcare professionals (HCPs) to refer individuals who do not benefit from medical and/or psychological treatment to a range of voluntary, community, and social (VCS) services to support their health and wellbeing [1,2]. It is suggested that SP increases social networks, which in turn may contribute to the quadruple aim by 1) improving population health and wellbeing, 2) reducing or stabilizing costs in health care and other sectors, and 3) improving individuals’ and 4) HCPs’ experience with care [3]. Several processes underlie the effects of SP including social support, social connectedness, and loneliness reduction [4,5]. These processes, also called mechanisms are hypothesized to empower individuals with resilience, which in turn, reduces stress and improves wellbeing and other health-related outcomes. [6–9]. However, the extent to which SP affects such mechanisms and, in turn, quadruple aim outcomes are still unclear.

Since the implementation of SP in the UK, there is a growing interest in SP in other countries such as the Netherlands [10], Australia [11], and Scandinavian countries [12]. Although there is an overall aim within the different SP programs, the implementation and delivery varies per country, making it difficult to understanding the mechanisms for SP [13]. These variations in implementation are also reflected in the studies that evaluate SP, where there is a wide range of intervention characteristics (i.e. context, structure, and content of the interventions) [14–16]. Contextual characteristics refer to elements such as the delivery setting and target group. While most SP studies focus on general populations with psychosocial factors, some target specific groups like young people [17], individuals with type 2 diabetes [18], or families with preschool children [19]. Structural characteristics include type of referral pathways, collaboration between professionals, the intensity and duration of SP and its activities. Content characteristics include the available VCS-services and specific strategies used to implement and deliver the service. Due to variations in intervention and study characteristics, it is challenging to determine what contributes to the influence of SP.

### 1.1. Review objectives

To understand the influence of SP and clarify what constitutes the best SP practice and process, it is important to identify both the intervention characteristics as well as mechanisms that contribute to different health and wellbeing outcomes. With this systematic review, we aimed to 1) identify intervention characteristics, mechanisms, and quadruple aim outcomes in SP research and 2) explore how these characteristics and mechanisms relate to the influence of SP on the quadruple aim outcomes.

## 2. Methods

This systematic review was registered in PROSPERO on April 2022 (CRD42022319765). We followed the Preferred Reporting Items for Systematic Reviews and Meta-Analyses (PRISMA) guidelines (Appendix A) [20].

### 2.1. Search Strategy

We searched multiple databases including Medline, PubMed, Embase, Web of Science, Cochrane Library, PsycINFO, Emcare, Epistemonikos database, Academic Search Premier, Social Services Abstracts, Sociological Abstracts, ERIC, NHS economic evaluation database / Health Technology Assessment database on November 9, 2021 and April 6, 2023 (update). Since the target population for SP and outcomes of interest varied widely, we did not use the PICO method for search term specification. Instead, our search terms included keywords and phrases related to ‘social prescribing’, ‘arts on prescription’, ‘social intervention’, or (‘link worker’ and ‘primary care’) (Appendix B). Additionaly, on November 15, 2022 we searched the first 200 hits relating to ‘Social Prescribing’ and ‘Welzijn op Recept’ in Google Scholar to identify grey literature and we searched all references from the included papers [21]. The search strategy was developed in consultation with an information specialist from the Leiden University Medical Center.

### 2.2. Eligibility criteria

#### Population

We included papers primarily reporting on interventions targeting individuals seeking primary care for psychosocial problems, such as those outlined by Heijnders and Meijs (e.g., depression, anxiety, frequent healthcare visits, social isolation) [22]. Studies that exclusively included individuals with long-term conditions and did not account for the presence of psychosocial problems were excluded from the analysis. We also excluded papers that solely focused on individuals under eighteen.

#### Intervention

We included papers reporting on interventions primarily focusing on the transfer from the primary care to any VCS-based activities aimed at increasing individuals’ wellbeing. We excluded papers focusing on interventions in which individuals are referred to VCS organizations from another setting than primary care (e.g., exercise referral schemes for clinical depression or cardiac rehabilitation) and interventions in which the primary aim is not to increase individuals’ wellbeing (e.g. exercise referral schemes to increase physical activity in sedentary adults).

#### Comparator

No comparator criterium was applied since we included all types of empirical studies.

#### Outcome

We included papers reporting on outcomes related to the dimensions of the quadruple aim: 1) health and wellbeing, 2) health care use and costs, 3) HCPs’ and 4) participants’ experience with SP. Papers without the focus on at least the first two dimensions were excluded.

#### Type of study

We included papers reporting on (non-)randomized controlled trials (RCTs), observational studies, quasi-experimental designs, and qualitative studies written in English or Dutch. Studies in other languages were excluded due to language barriers of the authors. Since 2010, there has been an increased recognition and adoption of SP by HCPs’ and a growth of community-based organizations. Accordingly, we limited our inclusion criteria to papers published from 2010 onward.

### 2.3. Selection of papers

Duplicates were removed in Endnote (version 20). One researcher (EMD) conducted the title and abstract screening using the AI-aided screening tool ASReview [23], which combines machine learning with an active learning model to predict inclusion likelihood [24]. A dataset containing all references retrieved from the search was imported into ASReview. Based on a study examining active learning model performance [25], we selected the Naïve Bayes as the classifier to make relevancy predictions and the TF-IDF as the feature extraction technique (i.e., a technique to numerically represent textual content as feature vectors, which allows the classifier to predict relevancy). Certainty-based sampling was used as the query strategy to prioritize relevant papers. Screening was stopped after assessing 25% of all papers to minimize time spent on irrelevant papers [25,26]. EMD reviewed the full-text papers for inclusion, and a random 20% sample was cross-checked by two additional researchers (FHA and MRC). Any disagreements were resolved by consensus, a fourth researcher (MLH) was consulted in case of uncertainty about inclusion.

### 2.4. Data extraction

Data extraction was performed by five researchers independently. To ensure consistency and optimize the data extraction protocol, a random sample of seven studies was assessed by five researchers (EMD, FHA, SGLS, MB, and LAV) and the assessment of the remaining papers was divided among the researchers. Following Cochrane guidelines [27], a coding system was developed to code all intervention characteristics that could moderate the influence of SP. We extracted data on contextual (e.g., participants, delivery setting), structural (e.g., presence of a link worker, activity duration), and applied behavior change techniques (BCTs) as content characteristics. BCTs are strategies and methods to promote the adoption or modification of behaviors, such as goal-setting, self-monitoring, and social support (Appendix C) [28]. Outcomes related to the quadruple aim and mechanisms (e.g., social connectedness, self-confidence, knowlegde) were also extracted [4,5]. Statistical analysis details were coded as they varied between studies. Based on descriptive statistics or significance of inferential statistics, outcomes were classified as ‘improvement, ‘deterioration, or ‘no influence’. Additionally, information on citation details, study design, study method, country, and intervention name was extracted from each included paper. The percentage agreement was calculated and discrepancies were solved through discussion. MRC was consulted to solve any disagreement. Percent agreements for study characteristics, intervention characteristics, and outcomes were 94.2%, 89.2%, and 66.0% respectively (Appendix D).

### 2.5. Quality assessment and risk of bias

The Mixed Method Appraisal Tool version 2018 (MMAT) was used to appraise the quality of each included paper [29]. The MMAT is designed to critically appraise both qualitative, quantitative, and mixed method studies and is, therefore, suitable for systematic reviews with mixed studies. Every criterion is rated as ‘yes’ (1) or ‘no/can’t tell’ (0) for every applicable item. We rated each paper ‘low’, ‘moderate’, or ‘high’ quality based on the number of criteria met (1-2 = low, 3=moderate, 4-5=high). For mixed methods studies, the overall quality score was determined by the lowest score among the components [29]. Papers were not excluded based on their quality score, but scores were incorporated in the data synthesis. Percent agreement ranged from 68.0% (category mixed methods) to 92.0% (category qualitative studies) (Appendix D). Risk of bias was assessed for RCTs and non-randomized studies using the Cochrane RoB2 [30] and the ROBINS-I [31] respectively. Uncontrolled before-after and cross-sectional studies were considered at high risk of bias due to the lack of control groups, following the Cochrane Handbook for Systematic Reviews of Interventions [27].

### 2.6. Data Synthesis

Because of heterogeneity in the outcomes of the quadruple aim, we were not able to pool the results in a meta-analysis and meta-regression. We therefore used Harvest plots to graphically present the results [32]. Harvest plots allow for graphically displaying evidence from complex and diverse studies or results, wherein outcomes are non-identical or combining data through conventional meta-analysis is not feasible. Based on available data, we created plots for different outcomes related to the quadruple aim. Results were stratified for intervention characteristics and mechanisms, and we incorporated the quality assessment, the research methods, and the type of analysis used to assess the influence of SP (i.e., descriptive statistics and inferential statistics) in the plots. Plots were created using R Studio version 2022.02.3 [33]. We summarized qualitative data narratively to identify outcomes in papers reporting on qualitative methods and to explore the influence of SP on these outcomes.

## 3. Results

### 3.1 Study selection

In total, 6018 unique papers were retrieved. Title and abstract screening resulted in the exclusion of 5806 papers, and full-text screening resulted in the exclusion of 183 papers, resulting in 30 papers that met the inclusion criteria. After searching the reference lists manually and the search update, a total of 49 papers were included for data extraction (Figure 1). Appendix E provides a list of publications excluded in the full-text screening stage, including reasons for exclusion. The primary reasons for exclusion encompassed studies that did not specifically examine the influence of SP (e.g., process evaluations), lacked a primary care referral component, or involved interventions embedded within a mental health recovery program or an exercise referral program.

**Figure 1.**
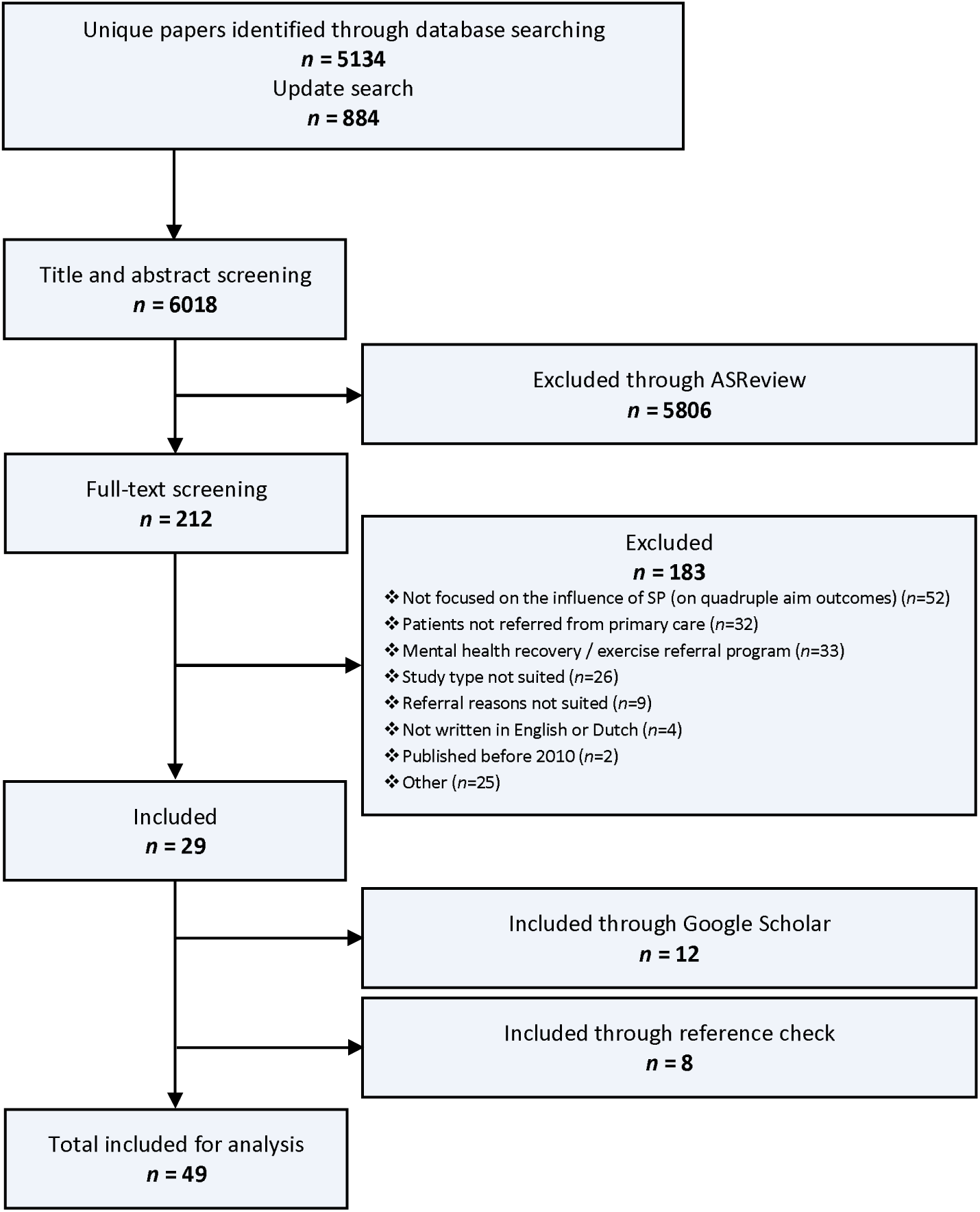
PRISMA flowchart.

### 3.2 Study characteristics

Seventeen used quantitative methods, mainly a before-after design [8,17,34–48], of which two also included a control group [40,48]. One paper described a RCT [41]. Seven papers reported on qualitative methods, using primarily interviews [10,49–54]. Furthermore, 25 papers reported on a mixed methods study in which mainly before-after designs (including one with a control group [55]) and interviews were used [2,6,55–77]. The majority of studies were performed in the UK (43, 87.8%). Other studies were performed in Australia [34,72], the Netherlands [10,48], Canada [49], and South Korea [38]. Characteristics of the included papers are shown in Table 1.

**Table 1.**
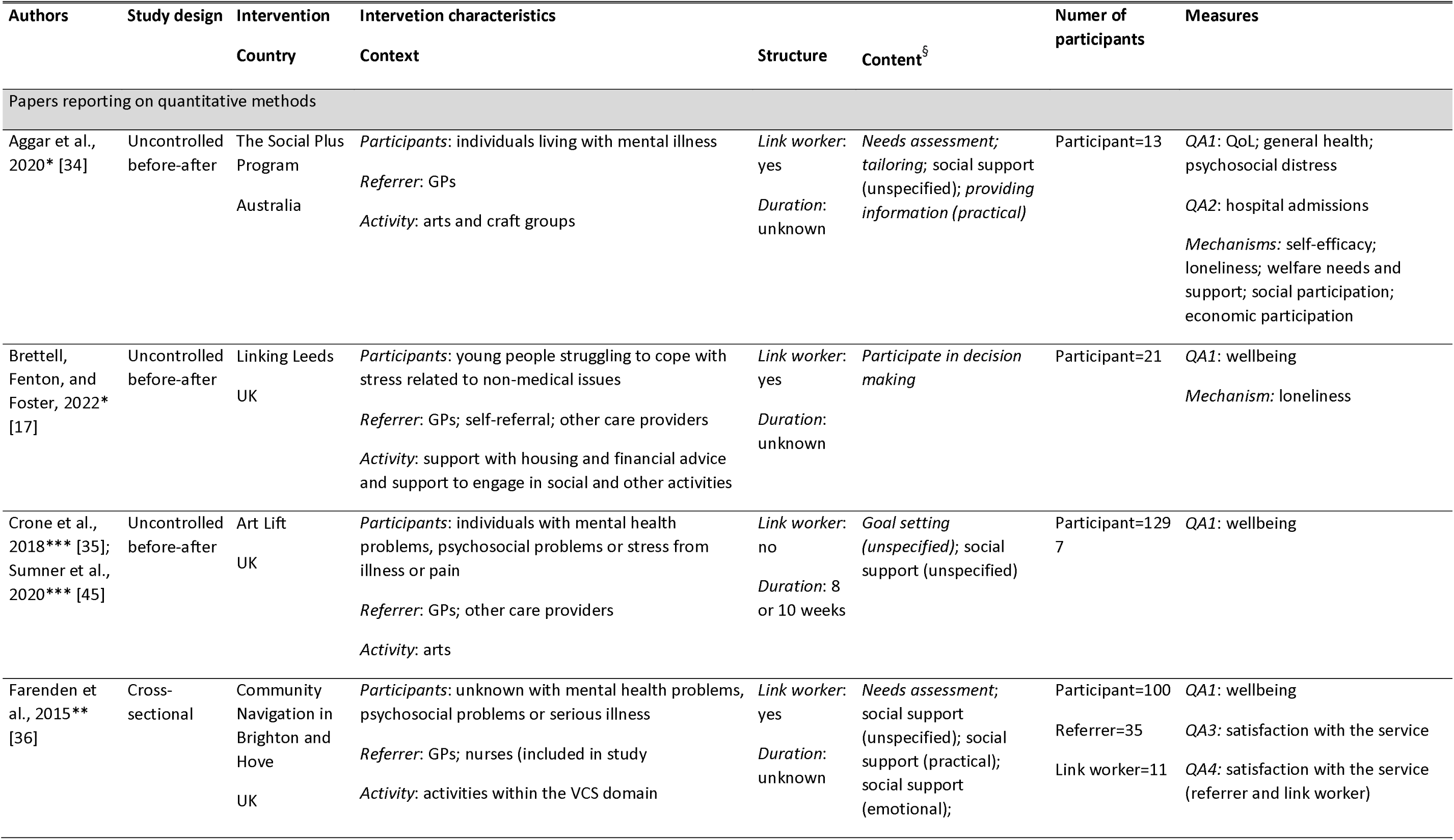
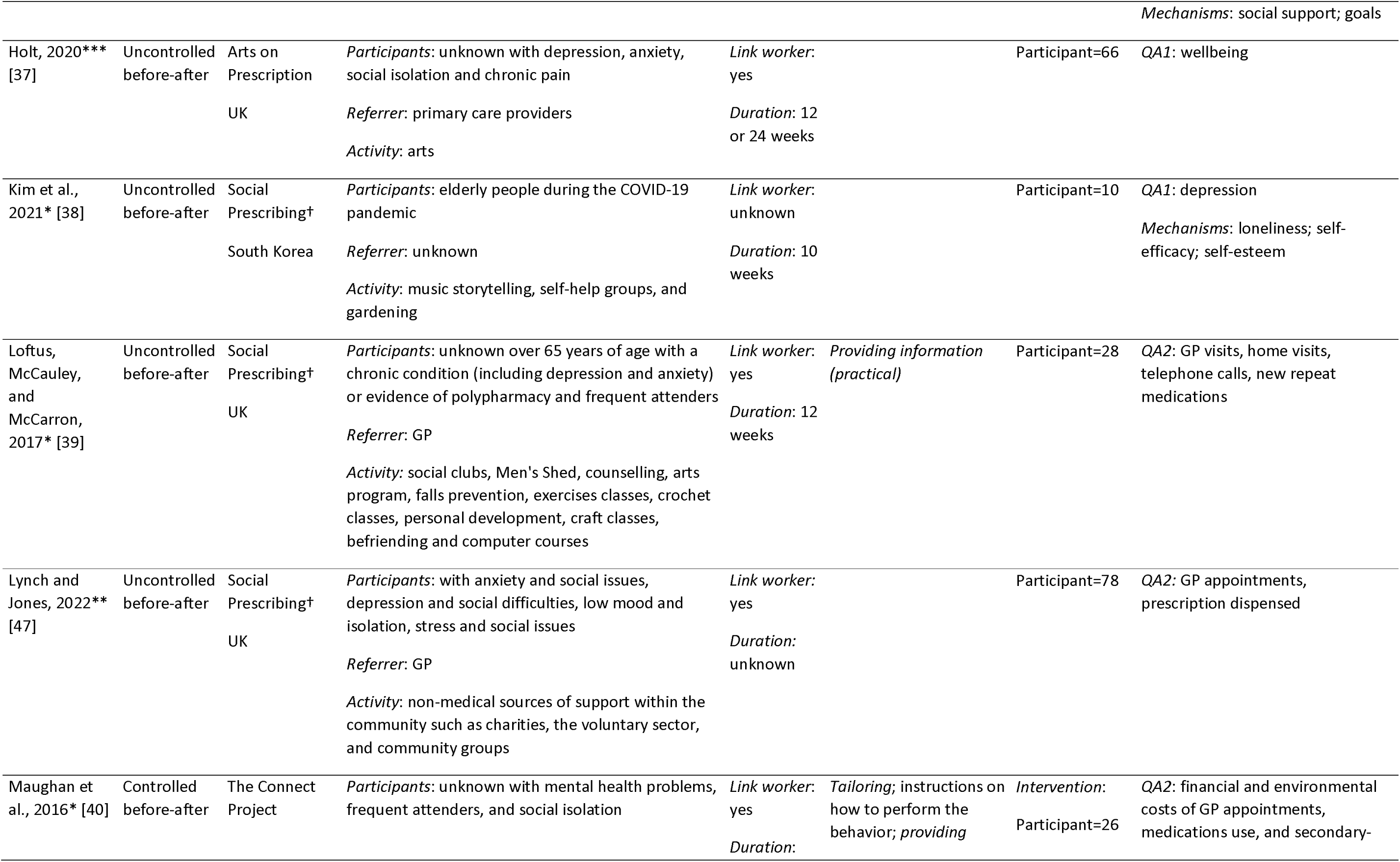
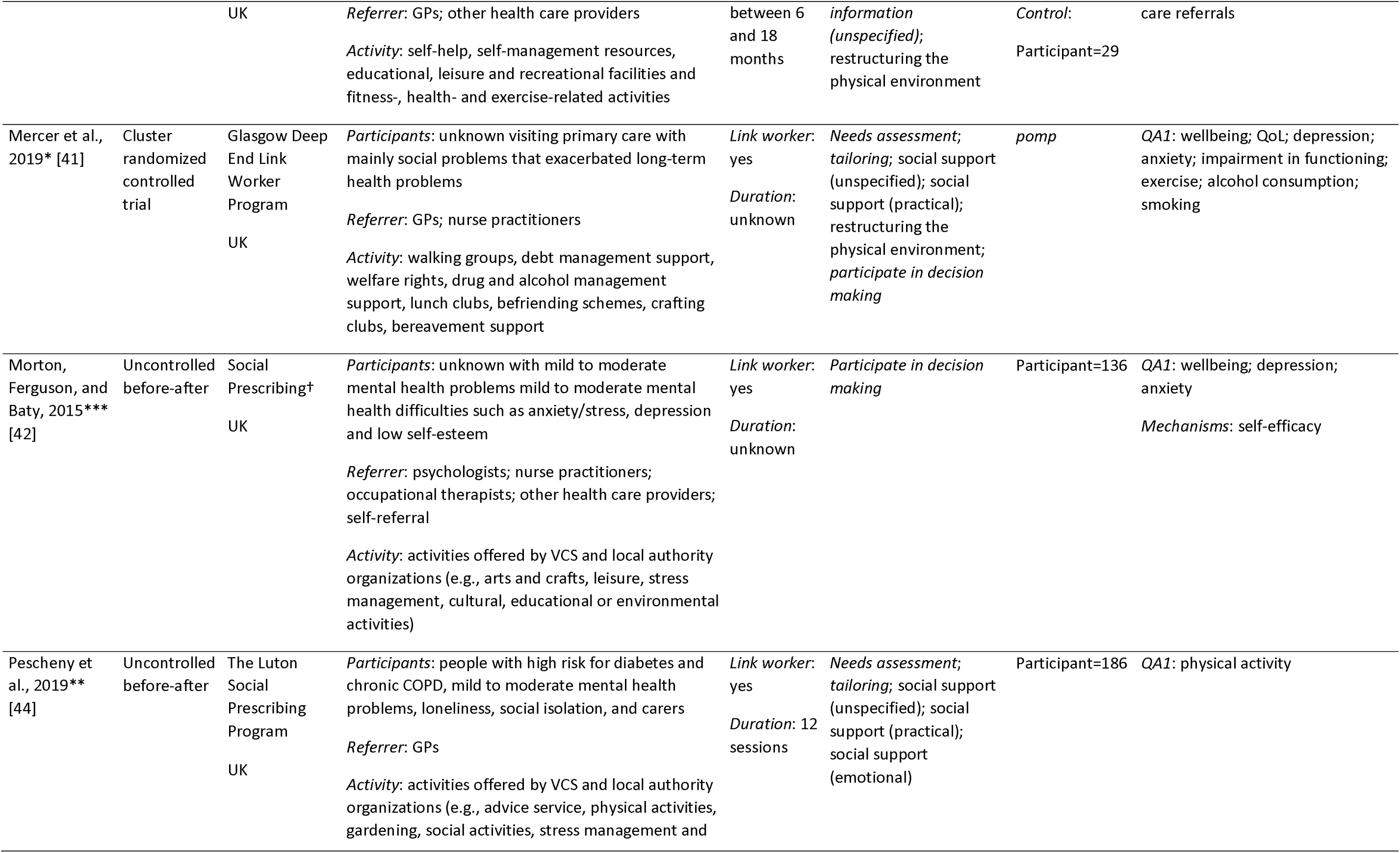
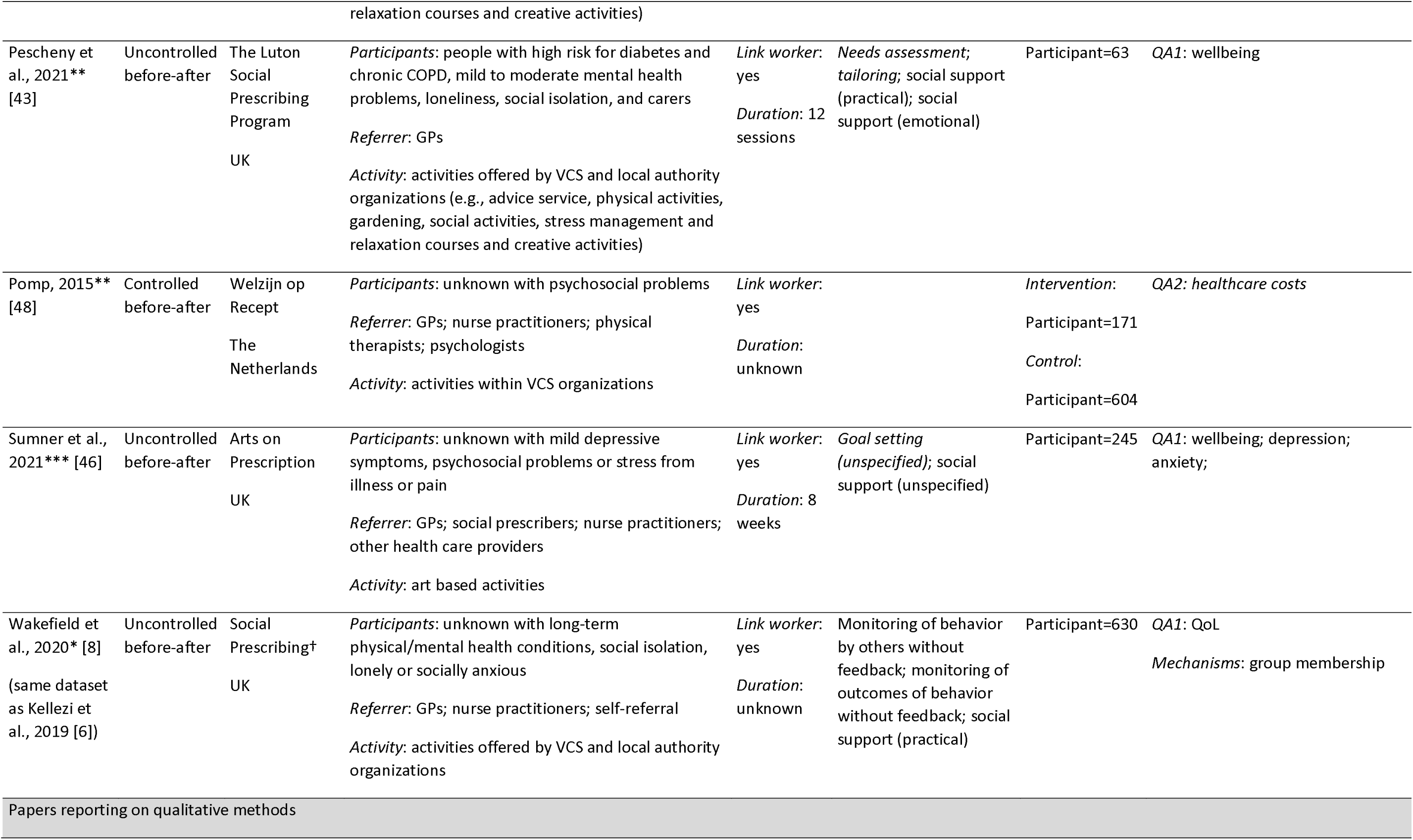
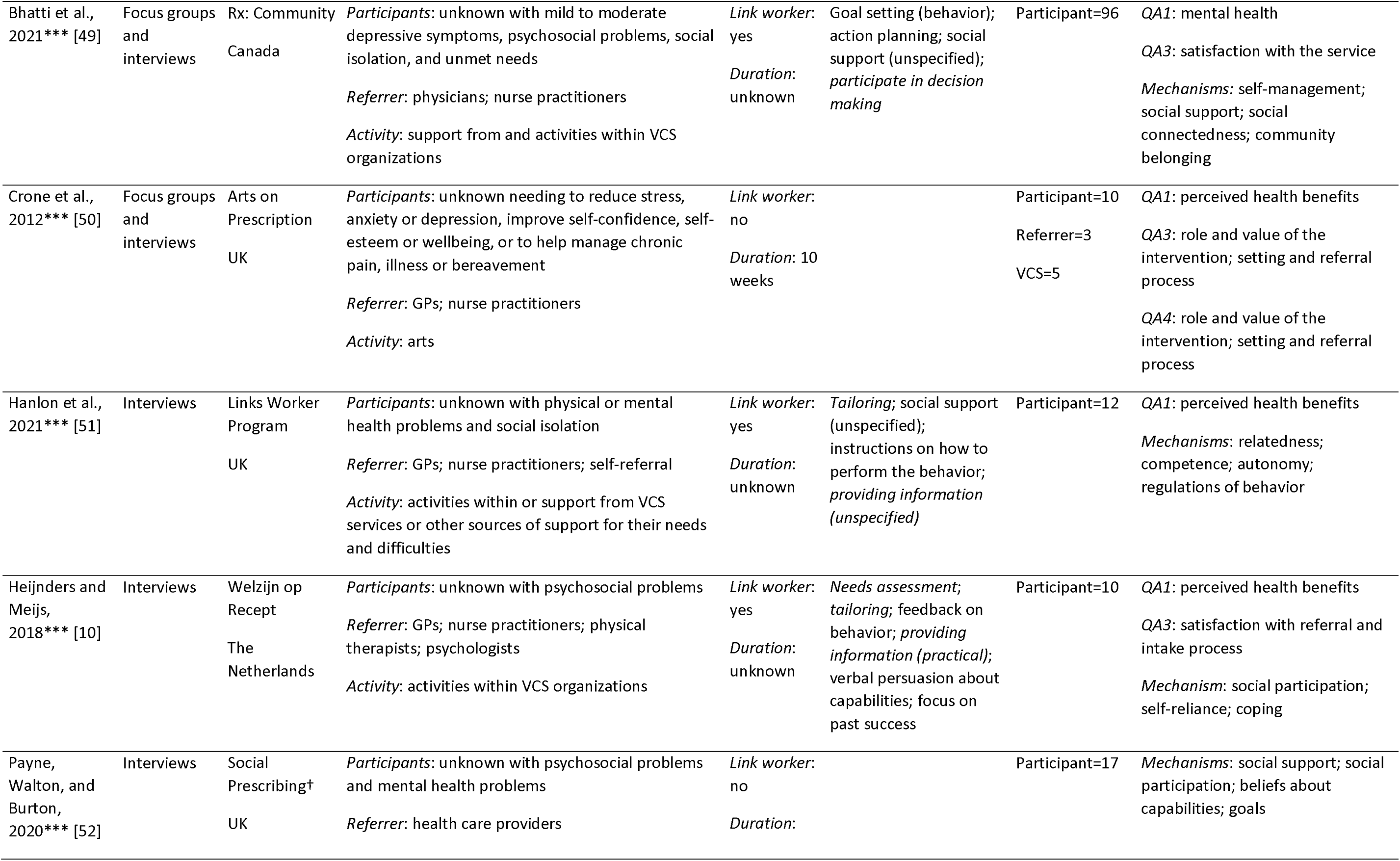
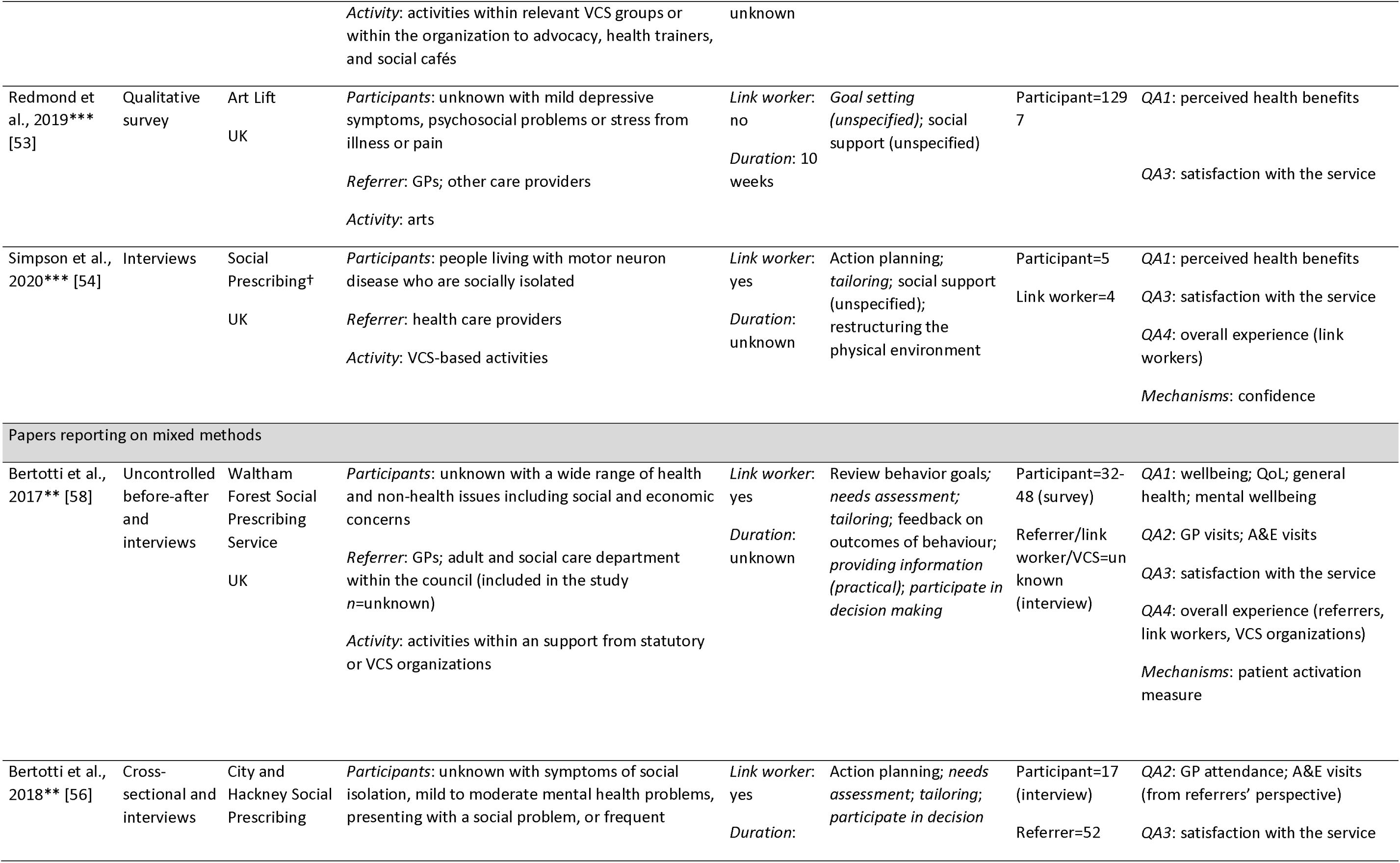
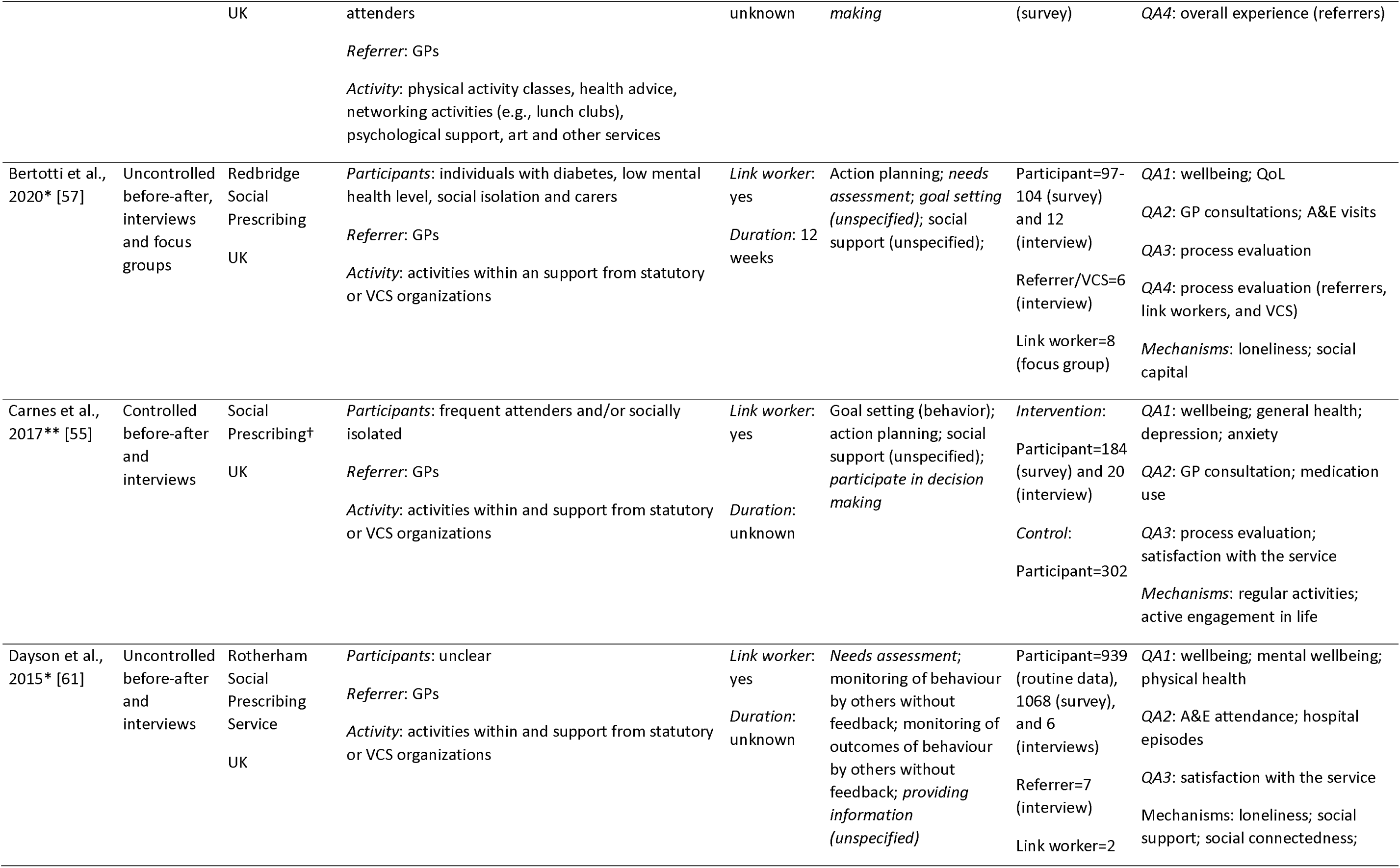
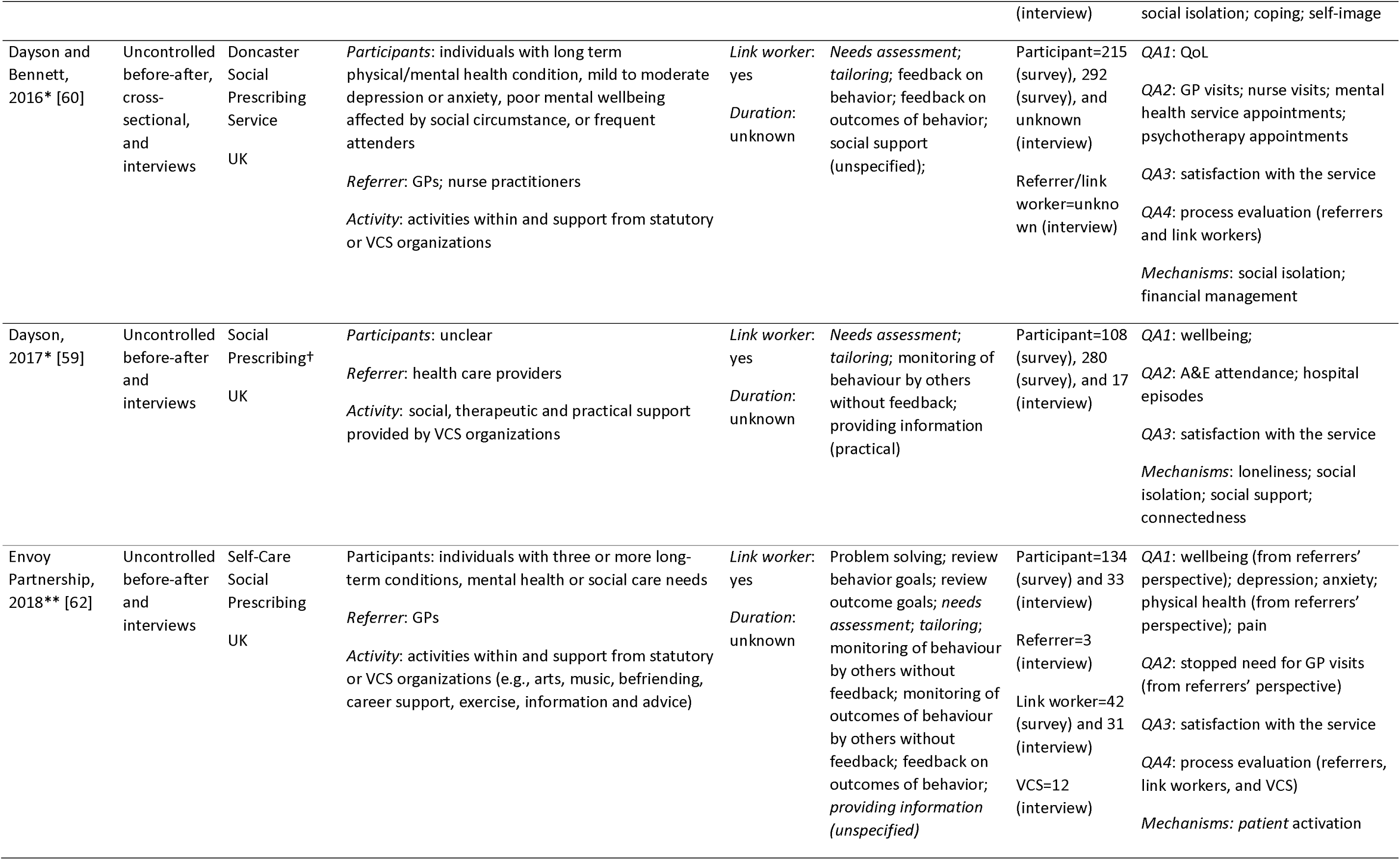
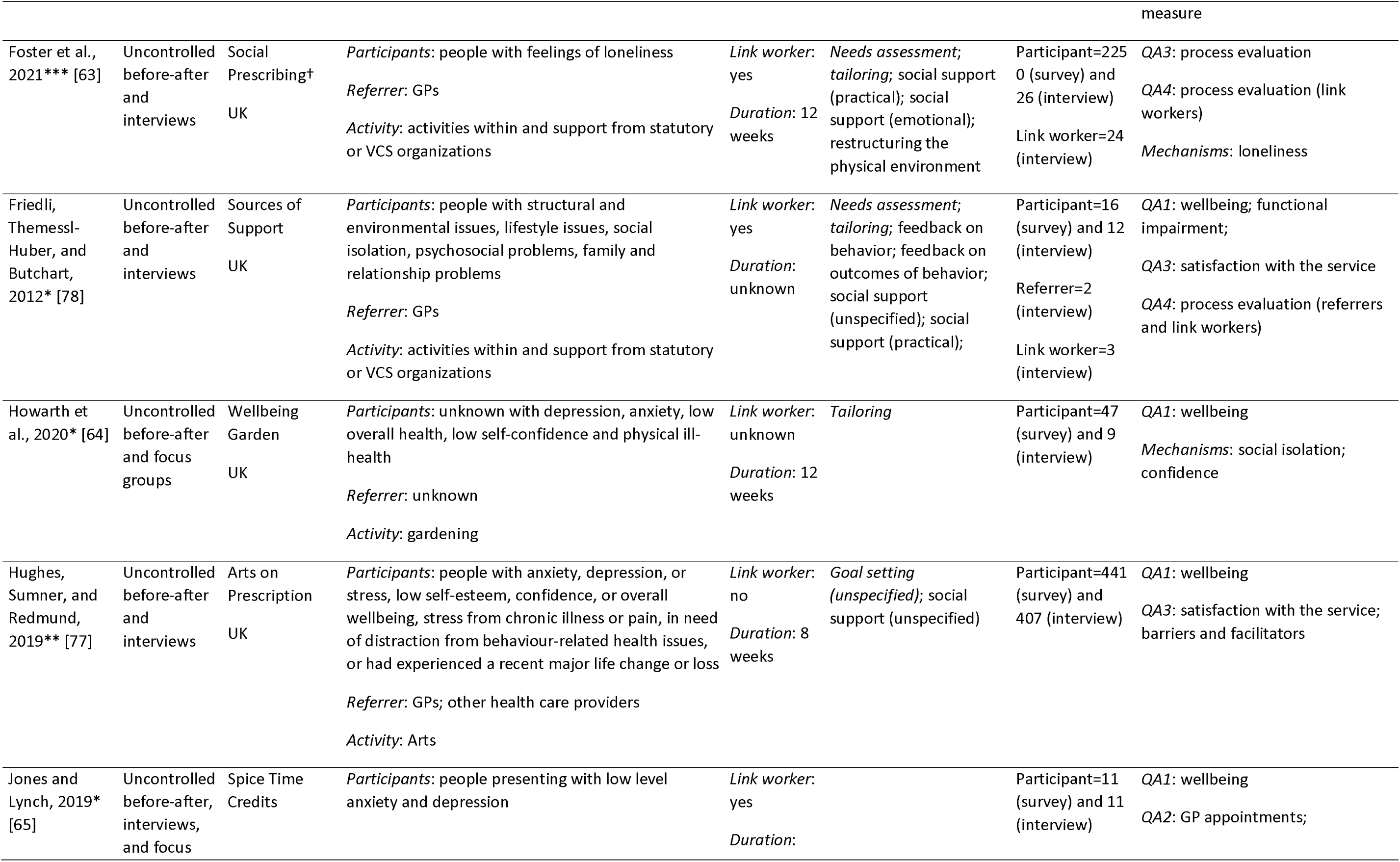
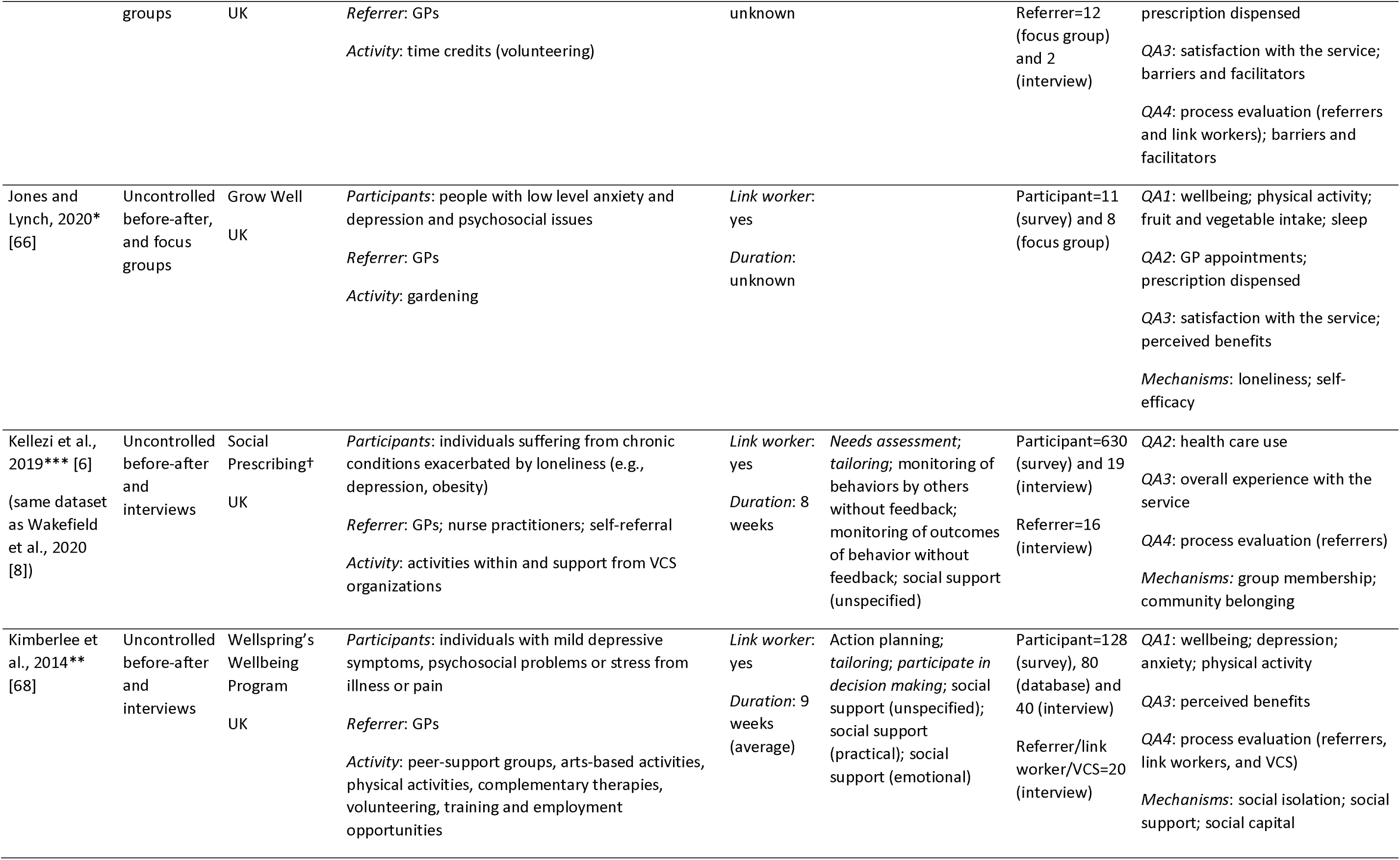
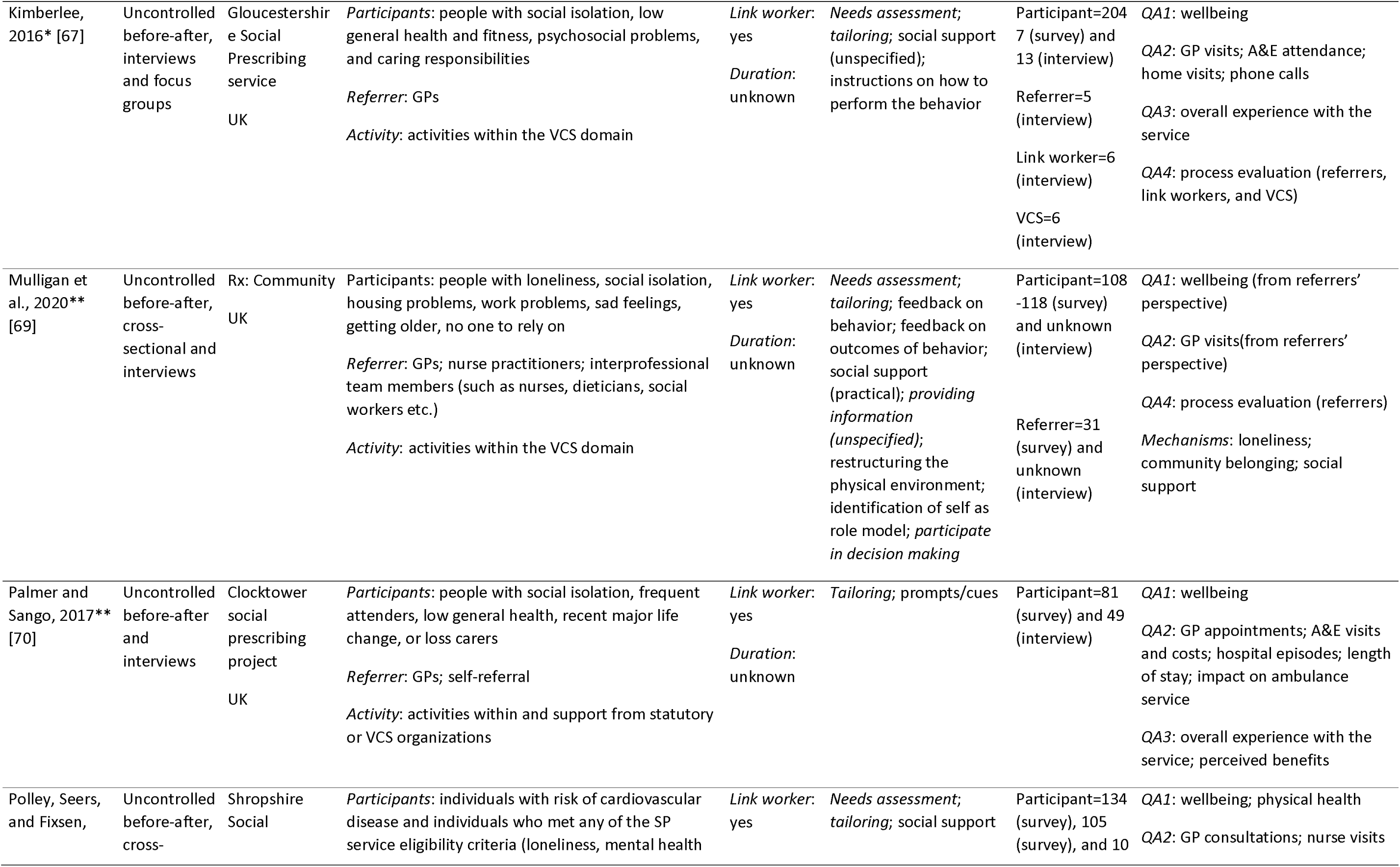
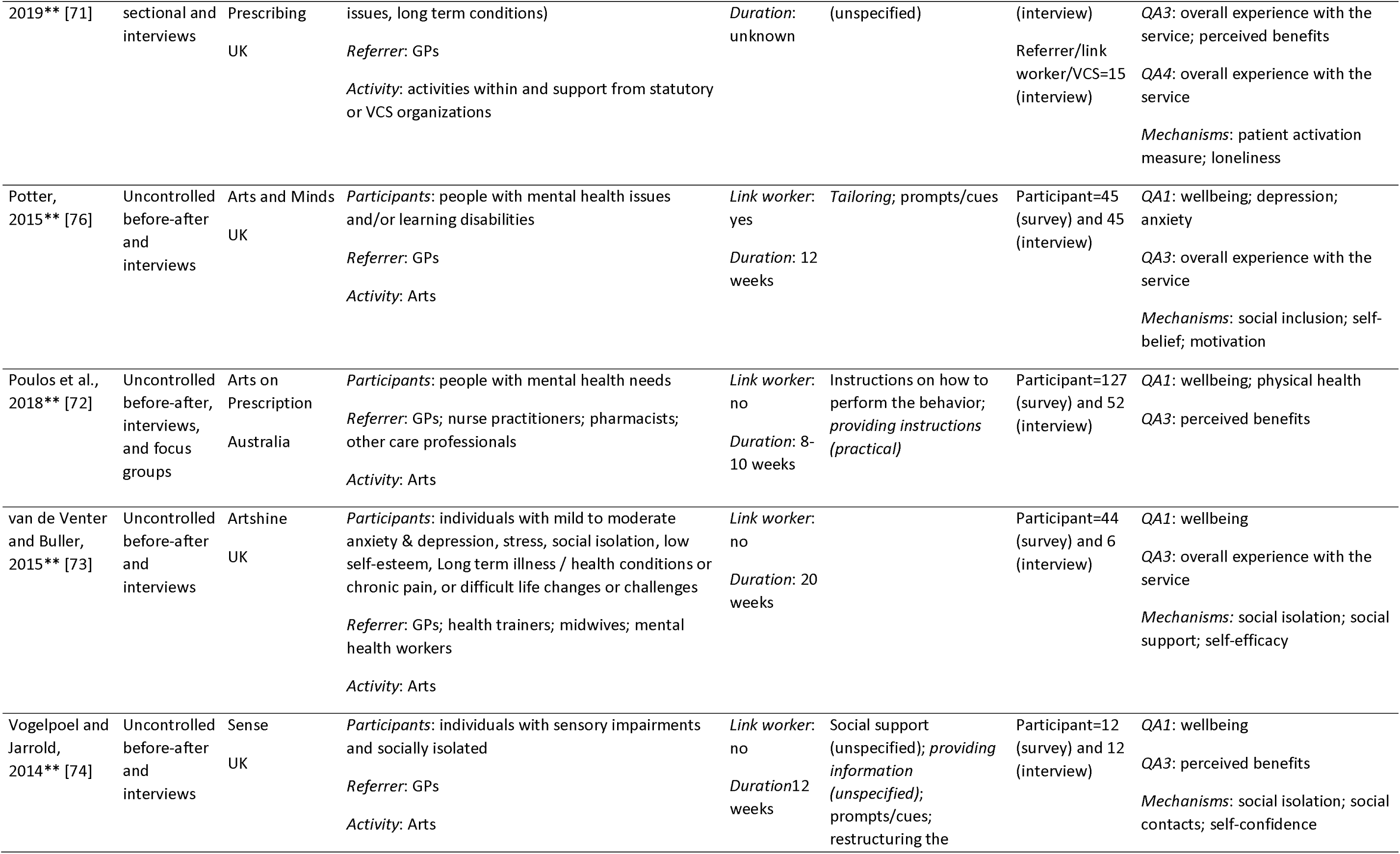
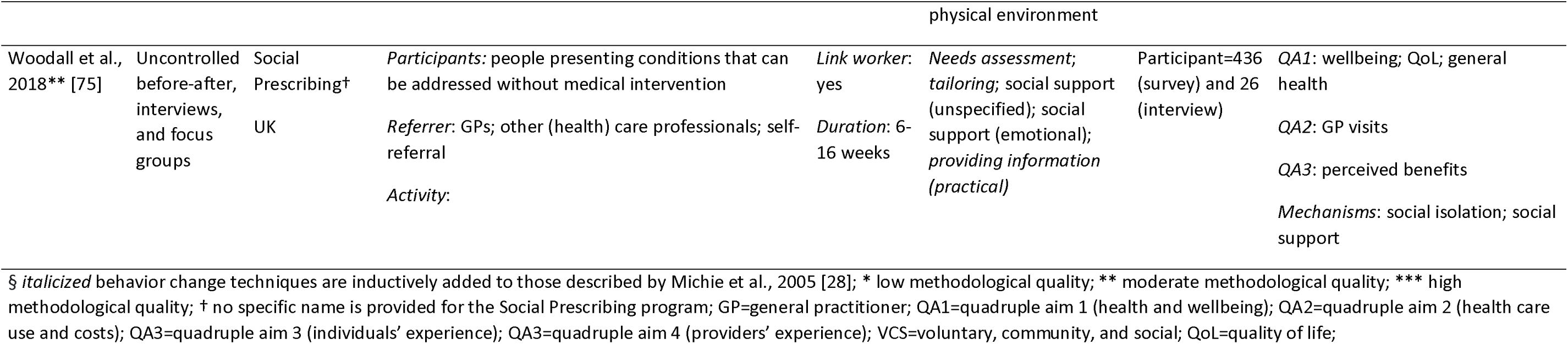
Characteristics of the included papers.

#### 3.2.1. Participant characteristics

For participants attending SP, baseline sample sizes varied between *n*=10 [38] and *n*=2250 [63] in papers reporting on quantitative methods and between *n*=5 [54] and *n*=1272 [53] in papers reporting on qualitative methods. All papers included in this review reported on a combined total baseline number of *n*=18631 participants (*n*=17084 receiving SP, *n*=1547 controls). The percentage of women ranged from 22% to 100% and participants’ baseline mean age ranged from 38 to 82 years. Within studies, reported referral reasons were diverse, with mild to moderate mental health problems, social isolation, frequent attenders, housing and financial problems, and significant life changes as commonly reported reasons. Only a few papers reported information on participants’ ethnicity, employment, living, and/or socioeconomic status [6,38,41,46,49,52,55,57,58,63,67–69,71,73,75–77]. Most of the participants were white, unemployed or retired, lived alone, and had a lower socioeconomic status. Sample sizes for SP service staff (i.e., referrers, link workers, and VCS staff) varied between *n*=4 [54] and *n*=42 [62]. Five studies did not report detailed information on participant and SP service staff sample sizes for the qualitative parts [56,58,60,61,69].

#### 3.2.2. Intervention characteristics

Based on the information provided in the papers, only 14.3% (*n*=7) stated that the referrer was trained or had SP-specific knowledge and skills [36,39,41,49,62,69,74]. In 73.5% (*n*=36) of the papers, a link worker was involved of which 32.7% (*n*=16) stated that the link worker was trained or had SP-specific knowledge and skills [10,36,37,39–43,55–57,63,68,69,71,75]. The most often used BCT was providing social support (53.1%, *n*=26), followed by tailoring (51.0%, *n*=25) and needs assessment (42.9%, *n*=21). Examples of social support were meeting for a cup of tea, attending or help with access to prescribed activities, or one-to-one support meetings (i.e., emotional, practical, and unspecified support).

### 3.3. Quality assessment and risk of bias

Of the papers reporting on quantitative methods, we rated five as high methodological quality, four as moderate, and eight as low. All seven papers reporting on qualitative methods were rated as high quality. Two papers reporting on mixed methods were rated as high, two as moderate, 21 as low quality. Overall methodological quality was low. Risk of bias was assessed for one cluster-RCT [41] and two controlled before-after studies [48,55]. Risk of bias was rated as high for the cluster-RCT and moderate for the latter two. The cluster-RCT had bias related to the timing of identification and recruitment of participants, as well as outcome measurement. The controlled before-after studies had bias related to confounding factors and selection of reported results. More detailed information is present in Appendix D.

### 3.4. Papers reporting on quantitative methods

#### 3.4.1 Outcome measures

The majority of studies measured participants’ wellbeing [17,35–37,41–43,45,46,55,57–59,61,62,64–78]. Other outcome measures were general and physical health, health behaviors [34,41,44,55,57,58,60,62,66,68,71,72,75,78], mental health [34,37,38,41,42,46,55,60,62,68,69,75,76], anxiety [41,42,46,55,57,60,62,68,75,76], and quality of life [8,34,41,57,58]. Wellbeing was most often measured using the (Short) Warwick Edinburgh Mental Wellbeing Scale [17,35,37,42,43,45,46,57,58,64–67,70,72–78]. Others used the ONS Personal Wellbeing Scale [17,57,68], the Measure Yourself Concerns and Wellbeing [57,71], the ICEpop CAPability measure for Adults [41], the Measure Yourself Medical Outcome Profile [55], or self-developed questionnaires [36,61,62,68]. A variety of instruments were used to measure general health such as the EuroQol Visual Analogue Scale [34,57,75], separate items from validated quality of life questionnaires [34,60], or self-developed questionnaires [55,58]. Polley et al., 2019 used physiological measures to measure physical health (i.e., blood pressure and body mass index) [71]. Validated questionnaires used were the Work and Social Adjustment Scale [41,78], the International Physical Activity Questionnaire [44,68], or separate items from validated quality of life questionnaires [60,62]. To measure health behaviors, self-reported measures such as physical exercise, fruit and vegetable intake, sleep, smoking, and alcohol consumption were most often used [41,66,72]. Mental health was measured using the Patient

Health Questionnaire [46,68,76], the Hospital Anxiety and Depression Scale [41,42,55], The Kessler Psychological Distress Scale [34], the Short Mood Scale [37], and the Geriatric Depression Scale [38]. Others used separate items from quality of life questionnaires [60,62,75] or a self-developed mental health measure [69]. Anxiety was measured using the Hospital Anxiety and Depression Scale [41,42,55], the General Anxiety Disorder scale [46,68,76], or separate items from validated (quality of life) questionnaires [57,60,62,75]. Different variations of the EQ-5D were most often used to measure quality of life [41,57,58]. Aggar et al., 2021 used the World Health Organization Quality of Life Brief Version [34] and Bertotti et al., 2020 used separate items of the ONS Personal Wellbeing scale [57] as quality of life related measures.

Regarding health care use and/or costs, the majority of papers analyzed primary health care use such as general practitioner (GP) visits [6,39,47,55,57–60,62,65–71,75], accident and emergency (A&E) visits [57,59–61,67,70], and GPs’ phone calls [39,67,68]. Secondary health care use, primarily hospital admissions, was analyzed in three papers [60,61,70]. Four papers analyzed medication use [39,47,55,66] and three analyzed health care costs [40,48,70]. The majority relied on self-reported measures to analyze the influence of SP on primary health care use and/or costs. Others used routine data [47,48,59,61,65–68,71] or providers’ perceived stopped need for health care use [62,69].

Only two papers reported outcomes related to providers’ and participants’ experience with SP and its process using self-developed questionnaires [36,71].

Several papers analyzed the influence of SP on outcomes considered as mechanisms. Social-related mechanisms such as loneliness, social connectedness, and social support were analyzed most frequently [6,8,17,34,36,38,55,57,60,63,64,66,68,69,71,75,76]. Other mechanisms measured were beliefs about capabilities (e.g., self-efficacy and confidence) [34,38,42,60,64,66,71,76] and participants’ and link workers’ knowledge and skills [36,60,76]. Loneliness was measured using the UCLA Loneliness Scale [17,34,38,63,66], the Campaign to End Loneliness Measurement tool [57,69,75], or the De Jong Gierveld Loneliness Scale [71]. Outcomes related to social connectedness (e.g., social isolation, social participation, community belonging) were measured using the Friendship scale [68], the Social Participation Scale for the Elderly Living Alone [38], the Social Inclusion Scale [76], or self-developed items (based on validated questionnaires) [6,8,34,55,57,60,64,69]. Social support was measured using self-developed items (based on validated questionnaires) [34,36,57,69].

#### 3.4.2 Influence of SP on the quadruple aim

Harvest plots were created for the most frequent assessed outcomes, namely 1) wellbeing, 2) mental health, 3) general and physical health, 4) primary health care use, and 5) social-related mechanisms.

Figure 2A through C shows the influence of SP on participants’ wellbeing, mental health, and general and physical health, respectively. In total, 25 papers reported improved wellbeing outcomes and only four papers reported no influence on participants’ wellbeing [41,55,57,58]. Sixty-two percent of the improvements were based on inferential statistics. None of the papers including a control group showed positive results [41,55]. Results related to mental health were generally positive. Ten papers reported improvements of which 70% were based on inferential statistics, while the remaining three - including those with a control group-reported no influences [34,41,55]. Results for general and physical health and health behaviors were mixed. About half of the papers reported positive results for general and physical health [34,57,62,68,71,75,78], while the other half - including those with a control group-reported no influence [34,41,55,58,60,71,72]. Health behavior measures such as smoking, alcohol consumption, exercise, and sleep were also mixed [41,44,55,60,66,72].

**Figure 2.**
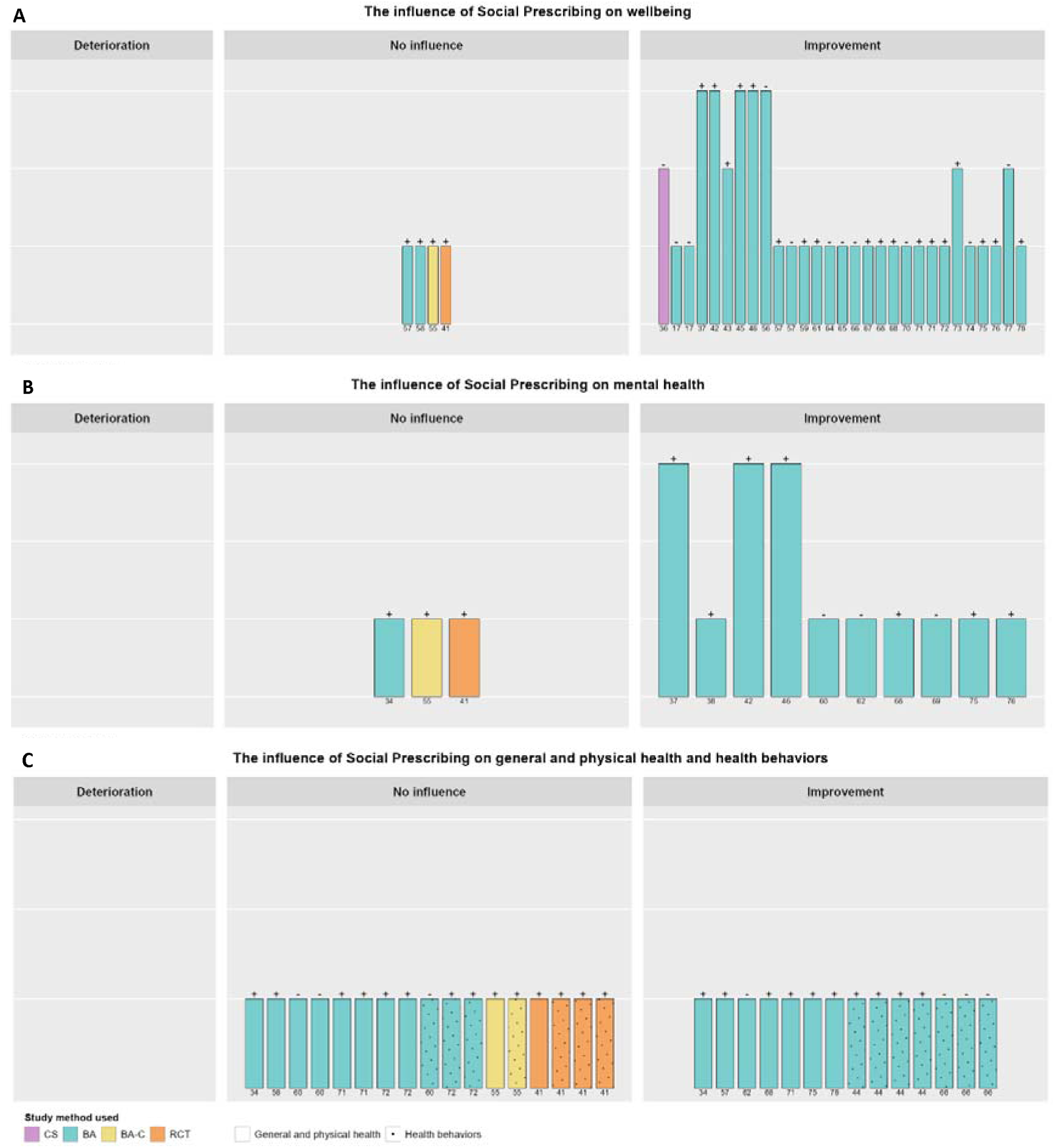
The influence of Social Prescribing on wellbeing (A), mental health (B), and general and physical health and health behaviors (C). The height of the bars represents the methodological quality; the symbols on top of the bars represents the statistical analysis: + = inferential statistics and - = descriptive statistics; the numbers below the bars represents the reference number; CS=cross-sectional; BA=before-after; BA-C=before-after with controls; RCT=randomized control trial.

Results on primary health care use were mixed (Figure 3). Thirteen papers showed a reduction in primary health care use and eight showed no changes. Palmer et al., 2017 showed an increase in GP visits [70]. In only 38.5% of the cases, inferential statistics were used to analyze changes in primary healthcare use.

**Figure 3.**
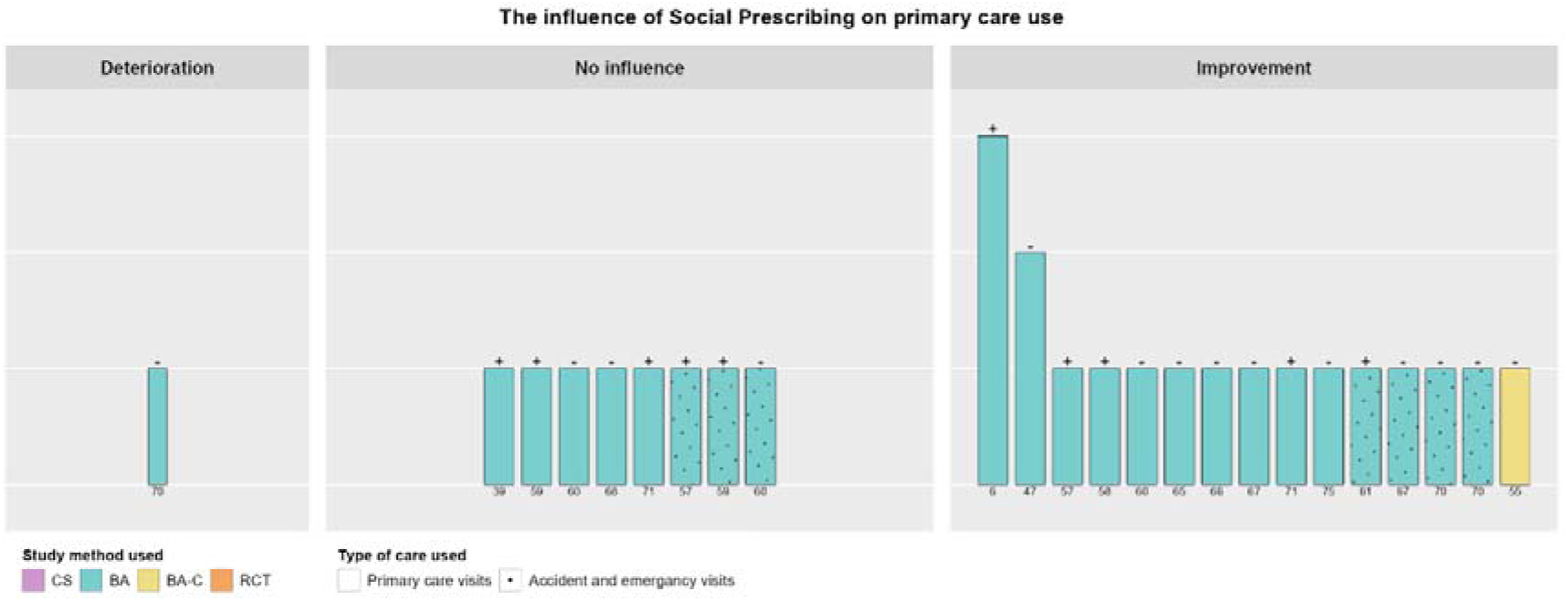
The influence of Social Prescribing on primary health care use. The height of the bars represents the methodological quality; the symbols on top of the bars represents the statistical analysis: + = inferential statistics and - = descriptive statistics; the numbers below the bars represents the reference number; CS=cross-sectional; BA=before-after; BA-C=before-after with controls; RCT=randomized control trial.

Papers that reported on the influence of SP on social-related mechanisms are shown in figure 4. Most papers reported positive outcomes [6,8,17,36,38,57,60,63,64,66,68,69,75,76]. In 44.0% of these cases, inferential statistics were used. In papers that reported no influence, all used inferential statistics to analyze the influence of SP on social-related mechanisms [34,55,57,71].

**Figure 4.**
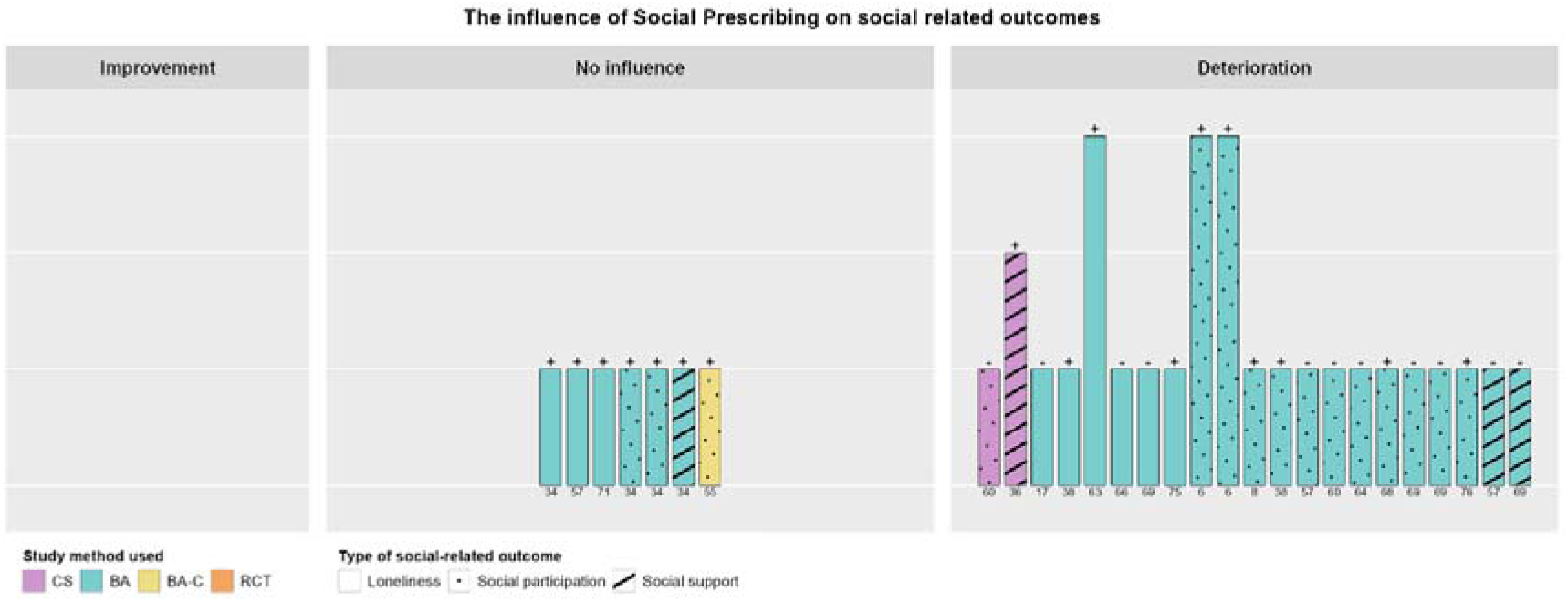
The influence of Social Prescribing on social-related mechanisms. The height of the bars represents the methodological quality; the symbols on top of the bars represents the statistical analysis: + = inferential statistics and - = descriptive statistics; the numbers below the bars represents the reference number; CS=cross-sectional; BA=before-after; BA-C=before-after with controls; RCT=randomized control trial.

#### 3.4.3 Influence of SP stratified for intervention characteristics and mechanisms

Stratified results for gender, age group, presence of a link worker, applied BCTs (i.e., social support, tailoring, and needs assessment), and social-related mechanisms are shown in Appendix F. Regarding participants’ wellbeing, results showed that participants with positive results on social-related mechanisms also showed positive results on wellbeing scores. This was also shown in results on mental health. Moreover, most improvements in mental health were found in studies with primarily women (i.e., >73%). General and physical health tended to be more often improved in participants between 50 and 65 years. Furthermore, SP seemed less effective on general and physical health for women and in interventions without a link worker or when (based on the information provided in the papers) needs assessments, tailoring, and social support were not incorporated in the interventions.

Regarding primary health care use, improvements were more often found in studies where both men and women were included and in studies where (based on the information in the papers) needs assessment was incorporated in the intervention.

Stratified results for social-related mechanisms did not show any differences.

### 3.5. Papers reporting on qualitative methods

#### 3.5.1. Influence of SP on health, wellbeing, and health care use

Participants reported a positive influence of SP on wellbeing [51,59–61,63,70,77], mental health [49,53,68], and general and physical health [6,10,53,61]. These improvements were also perceived by SP service staff (i.e., referrers, link workers, and VCS staff) [6,50,65]. Furthermore, few studies found that SP had a positive influence on health-related behaviors such as physical activity and smoking [10,71].

Only few studies discussed the influence on healthcare use. Some studies reported that GPs noticed that participants were making fewer GP appointments [65,67], while in other studies participants’ perspective on their healthcare use was mixed [56,60].

#### 3.5.2. Mechanisms

The most discussed outcomes considered as a mechanism were social-related, namely 1) from the participants’ environment (i.e., loneliness, social connectedness, and social support from peers) [6,10,49–53,59–61,66,70,73–77] and 2) from the SP service staff (i.e., social support) [6,10,49,51,52,56,59,60,63,67,71]. Regarding the social-related mechanisms from the participants’ environment, joining the activities offered within SP was of major influence in the social lives of the participants. Having a regular group to go to, establishing new friendships, engaging with others, and sharing experiences contributed to reduced feelings of loneliness, social isolation, and improved social connectedness, which in turn improved their wellbeing [10,51,71,77]. Social support from the SP service staff was considered as particularly important to participants. They described the importance of having someone to express themselves to; someone who was prepared to listen [6,49,51,56,60,61,71]. Referrers and link workers also informed participants about suitable services and facilitated access to these services,, providing some practical support [6,10,52,56,60,67]. Nevertheless, studies also discussed the challenges in the dose and duration of support, especially for participants with complex problems [6,10,63]. Foster et al. (2021) highlighted participants’ challenges to maintain improvements in loneliness after the support finished. In this study, participants indicated to miss the trustful relationship they had developed with the link worker, or they were reliant on them to attend activities [63].

SP also increased participants self-confidence, which can also be seen as a mechanism for the effects of SP. Participants confidence to join activities and interact with others was mediated by among others support from link workers and VCS staff [6,10,52,54,56,60,71]. Also, due to participating in various activities, participants gained confidence in for example, picking up old or exploring new hobbies, and engaging in volunteering, for example, as a health and wellbeing champion [6,50,51,61,63,73–75]. Participants’ confidence and ability to manage or cope with their (long-term) problems was also improved due to SP [10,49,53,59–61,78].

Another reported mechanism was participants’ optimism regarding SP as well as their future perspective. They were motivated to explore new things and were hopeful to benefit from SP [49,50,72,75–77]. Furthermore, participants regained perspective over their lives, developed a positive outlook on their present situation, and gained a sense of purpose [10,50–52,61,66,74–76].

#### 3.5.3 Participants’ experiences with the service

Most of the participants had positive overall experiences with SP [57,64,66,73]. However, the study of Carnes et al. (2017) reported that only half of the participants were satisfied with the SP service and the majority of participants in the studies by Bertotti et al. (2017, 2018, 2020) had poor experience with SP [55,57,58]. In the latter studies, participants argued that the time between the referral and the first contact (i.e., 2 months) was too long. Additionally, the study by Polley et al. (2019) demonstrated the value of communication between link workers and participants [71]. Shortly after receiving the referral letter, participants were called by the link worker to provide them with more information on SP. This was considered important, especially for those with low agency and those who were unlikely to proactively book an appointment by themselves. In several studies, participants reported challenges with engaging with the services because of their lack of knowledge about SP, psychological and physical problems, or aspects of participants’ situation or environment such as home, family, or financial barriers [56,57,63,65,66]. Challenges in engaging participants in SP were also reported by link workers [63].

#### 3.5.3 SP service staff experiences with the service

Overall, SP service staff acknowledged the potential of SP in improving participants’ health and wellbeing and reducing healthcare use [6,49,54,57,69,78]. However, some issues were discussed on the implementation of SP. First, skilled and knowledgeable referrers and link workers was a recurring topic [34,54,56–58,60,63,65,67,78]. Although referrers and link workers perceived the training on SP as important for its implementation [56,57,60,65], they lacked specific skills regarding for example, social security benefits and entitlements or how to deal with participants with complex needs [54,65,67]. Referrers also highlighted the value of link workers’ communication skills [60]. Knowledge on suitable sources of support within the VCS domain was also considered important [56,60,65,69,78]. Bertotti et al. (2018) argued that poor knowledge on VCS domains leads to obstacles to referral [56]. The importance of skilled and knowledgeable referrers and link workers was also confirmed by participants [63,67]. Second, the balance between demand and supply was also an issue [54,57,58]. This concerned both the capacity of the SP service as well as the capacity of services within the VCS domain. Other issues that inhibited the implementation of SP were referrers’ perceived high workload [65] and recruiting volunteers to support participants [63].

## 4. Discussion

With this systematic review, we aimed to identify intervention characteristics, mechanisms, and outcomes in SP research and to explore how these characteristics and mechanisms relate to the influence of SP on the quadruple aim outcomes. In total, 49 papers were included, of which seven qualitative, seventeen quantitative, and 25 mixed method studies. Quantitative data revealed that SP has a positive influence on the wellbeing and mental health of participants, as well as social-related mechanisms, such as loneliness, social connectedness, and social support. Notably, the participants with an improved wellbeing and mental health were also more likely to show improvements in social-related mechanisms. The qualitative data confirmed these findings by revealing the importance of social-related mechanisms on participant health and wellbeing and showed the importance of increased self-confidence and optimism. Additionally, according to the qualitative input, the quality of the SP service, including knowledgeable referrers and link workers with effective communication skills and sufficient capacity within the service and the VCS organizations, is a crucial part in facilitating positive experiences for participants and SP service staff. The quantitative findings, however, were primarily based on uncontrolled before-after studies and the analysis of controlled studies revealed no significant effect of SP on quadruple aim outcomes and mechanisms. Therefore, the findings in our study should be interpreted with caution.

The results in our review are consistent with recent evaluations on the influence of SP [79–82]. SP is a holistic approach that recognizes health as more than just the absence of disease, and acknowledges the importance of social networks, purpose, and meaning in overall (mental) wellbeing [14]. Our review supports this view by showing that participants who experienced improvements in social-related mechanisms also showed improvements in mental health and wellbeing outcomes. These social influences included both link workers and VCS services, as well as influences of participants’ personal environment and peers. Joining the activities offered within SP was of major influence in the social lives of the participants. Having a regular group to go to, establishing new friendships, engaging with others, and sharing experiences contributed to reduced feelings of loneliness and improved social connectedness, which in turn improved their wellbeing [10,51,71,77]. Mercer et al. (2019) highlighted the importance of social support from link workers [41]. Despite no overall significant differences between the intervention and control group, the authors showed that participants who saw the link worker three or more times had significant improvements in their health and wellbeing. However, we identified that some link worker reported that providing the right dose and duration of support was challenging. While some participants may require ongoing support to maintain behavior change and sustain the benefits of SP, others may require less intensive support once they have established connections with VCS services. Therefore, we advocate for the regular assessment of participants’ progress and requirements by link workers or referrers to determine the necessity of sustained support. In addition, researchers are urged to evaluate the frequency of link worker sessions.

In addition, this review also confirmed the importance of participants gaining a sense of purpose. A sense of purpose after SP can help to sustain the positive influences of the intervention and promote long-term wellbeing [83]. Some SP interventions offer the opportunity for participants to volunteer as health and wellbeing champions [36,49,69]. Health and wellbeing champions can contribute to promoting health and wellbeing within their communities. They can share their personal experiences with SP and perceived benefits, act as mentors or coaches, and provide guidance and encouragement to help other participants engage in the service and achieve their goals [84,85]. Furthermore, health and wellbeing champions can identify opportunities for collaboration and partnership within the community, leading to new resources and support for SPs [84].

Several physical activities are offered in SP such as walking groups, yoga, and gardening, as these activities have been shown to have numerous health benefits that involve both the body and mind. The influence of SP on general and physical health, however, was mixed, which is in line with a recent review [80]. This may be due to the challenges participants face with starting and adhering to the referred activities. Our review showed that improvements in general and physical health were more often present in interventions with needs assessments, tailoring, social support, and with the involvement of a link worker. Participants who perceive an intervention as personally relevant, tailored to their needs, and those who felt supported might be more likely to be motivated to participate and adhere to the intervention. Another potential reason for the mixed results in this review is the use of different outcome measures (e.g., physiological measures, behavioral measures, and physical activity), making it difficult to compare results between studies and provide an overall conclusion about the influence of SP on physical health.

Moreover, it is important to note that most participants in prior research were referred to SP programs to address psychosocial and mental health concerns, raising questions about the suitability of general and physical health outcomes to draw conclusions about the influence of SP. To evaluate the influence of SP, research should include tailored measures in addition to fixed measures. For example, goal setting is an important component of SP as it enables participants to establish and work towards specific objectives, as confirmed by Cooper et al. (2022) [81]. SP prioritizes participant-centeredness, and the achievement of participant goals serves as a direct reflection of how well the intervention aligns with their needs and preferences. Considering the anticipated positive influence of long-term goal achievement on health-related outcomes [86], it is worth considering incorporating goal achievement as an intermediate outcome measure in future SP research.

### Strengths and limitations

To our knowledge, this systematic review is the first to use Harvest Plots to evaluate the influence of SP on different outcomes. Harvest Plots are valuable for evaluating SP given the diverse range of outcomes. They allow for a comprehensive assessment of evidence to identify strongly influenced outcomes and those needing further investigation. Furthermore, Harvest Plots help to detect patterns and inconsistencies in the data that guide clinical decisions and future research. Furthermore, we used several strategies to rigorously search for relevant literature such as ASReview, grey literature search, and handsearching.

Nevertheless, some limitations of this systematic review and the included studies should be noted. First, the lack of controlled studies in SP research makes it difficult to determine the influence of SP on health outcomes, as observed changes may be due to other factors, which is also highlighted by several other reviews [81,87,88]. Additionally, a great amount of the studies reporting positive outcomes were based on descriptive statistics, which raises questions about the significance of the results. It is noteworthy that nearly none of the controlled studies reported significant improvements in outcomes evaluated in this review: only Carnes et al. (2017) showed significant improvements in GP consultations rates [55]. However, these findings should be interpreted with care due to the large number of controls and potential influence of regression to the mean rather than the effectiveness of the intervention. The authors acknowledged the challenges associated with incorporating an appropriate control group and randomization [41,55]. Including a control group in SP research is challenging due to the individualization of SP and the difficulty in standardizing them [89]. Additionally, SP is often implemented in real-world settings, such as community centers, making it difficult to control for confounding variables and ensure fidelity to the intervention protocol. Alternative methods to improve the practicality of randomized designs should be considered such as pragmatic trials or trial of intervention principles (meaning that the theoretical principles of the intervention are tested) [90].

Next, although the studies included in this review successfully reached individuals who were suitable for SP based on their employment, marital status, and socioeconomic status, they generally failed to include ethnic minority groups. Evidence suggests that these groups are at higher risk of experiencing psychosocial problems [91]. Such individuals may face multiple barriers to accessing healthcare services, which can limit their ability to access SP. Additionally, they may not recognize the potential benefits of SP or the intervention may not be designed to tailor to the specific language and cultural needs of individuals from ethnic minority groups [92]. Although reducing health inequalities is one of the core principles of SP [93], Moscrop argues that SP may exacerbate health inequalities by excluding those who are most in need and may not have the resources to engage with SP [94]. To address this issue, it is important to ensure that SP is culturally sensitive, inclusive, and accessible to all individuals, regardless of their ethnicity.

Finally, the data extraction process for intervention characteristics was solely based on information reported in the included papers, without additional insights from authors or websites. Therefore, the accuracy and completeness of our findings may be limited. Reporting intervention core principles is essential for researchers to comprehensively evaluate intervention effectiveness, assess fidelity, as well as the underlying mechanisms for producing its effects. Such information is crucial in determining the intervention’s efficacy and for improving its design in future studies. Also, most studies did not provide detailed information on the type, dosage, and duration of activities offered as part of SP. Consequently, we were unable to examine the influence of these factors on outcomes. As activities offered in SP can differ between countries, it would be valuable to investigate the influence of specific activities. For instance, in the Netherlands, basic needs concerns such as financial and housing issues are not considered as criteria for a referral to SP and thus advice on these issues is not offered [95], whereas, in the UK, SP also includes activities such as providing debt and financial advice, adult literacy classes, and job center support [96]. Therefore, it is important to assess the potential benefits of expanding the scope of SP to encompass a wider range of challenges.

## Conclusion

The quantitative findings suggest a positive influence of SP on participants’ mental health and wellbeing. Moreover, the findings highlights the importance of social-related mechanisms, including loneliness and social connectedness, in contributing to the observed positive influence of SP on mental health and wellbeing. These findings were supported by qualitative data, highlighting the importance of social influences and increased self-confidence and optimism resulting from SP participation. The study emphasizes the need for high-quality SP services with knowledgeable referrers and link workers, effective communication skills, and sufficient capacity to facilitate positive experiences for participants and SP service staff. We should, however, be cautious when interpreting these results due to limitations in study design, such as the lack of controlled trials, appropriate outcome measures, and statistical considerations, as well as the suboptimal reach of the target group. Therefore, while SP demonstrates promise in improving health and wellbeing outcomes, more rigorous and comprehensive research is needed to fully understand its influence and potential benefits.

## Supplementary Material

Appendix A: PRISMA reporting guidelines

Appendix B: Search Strategy for different databases

Appendix C: Behavioral Change Techniques Taxonomy (v1)

Appendix D: Results of the 1) Percent Agreement, 2) Quality Assessment Mixed Method Appraisal Tool, and 3) Risk of Bias Assessment Cochrane RoB2 and ROBINS-I

Appendix E: Overview of publications excluded in the full-text screening stage, including reasons for exclusion

Appendix F: Stratified Harvest Plots for wellbeing, mental health, general and physical health, primary care use, and social-related mechanisms.

## Supporting information

AppendixA_PRISMA

AppendixB_SearchStrategy

AppendixC_BCTTaxonomy_v1

AppendixD_PA-QC-RoB

AppendixE_ExcludedReferences

AppendixF_StratifiedResults

## Abbreviations

BCT: Behavioral Change Technique
HCP: Health Care Provider
SP: Social Prescribing
VCS: Voluntary, Community, and Social

## Declarations

### Data availability

The data used to support the findings of this systematic review are available from the corresponding author upon request.

## Acknowledgement

We would like to thank J.W. Schoones for his assistance in conducting the literature search.

## References

1. South, J., et al., Can social prescribing provide the missing link? Primary Health Care Research & Development, 2008. 9(4): p. 310–318.

2. Friedli, L., et al., Social prescribing for mental health—A guide to commissioning and delivery. Vol. 9. Care Services Improvement Partnership. 2008.

3. Bodenheimer, T. and C. Sinsky, From triple to quadruple aim: care of the patient requires care of the provider. The Annals of Family Medicine, 2014. 12(6): p. 573–576.

4. Carey, R.N., et al., Behavior change techniques and their mechanisms of action: a synthesis of links described in published intervention literature. Annals of Behavioral Medicine, 2019. 53(8): p. 693–707.

5. Connell, L.E., et al., Links between behavior change techniques and mechanisms of action: An expert consensus study. Annals of Behavioral Medicine, 2019. 53(8): p. 708–720.

6. Kellezi, B., et al., The social cure of social prescribing: a mixed-methods study on the benefits of social connectedness on quality and effectiveness of care provision. BMJ Open, 2019. 9(11): p. e033137.

7. Wakefield, J.R., et al., When groups help and when groups harm: Origins, developments, and future directions of the “Social Cure” perspective of group dynamics. Social and Personality Psychology Compass, 2019. 13(3): p. e12440.

8. Wakefield, J.R.H., et al., Social Prescribing as ‘Social Cure’: A longitudinal study of the health benefits of social connectedness within a Social Prescribing pathway. Journal of Health Psychology, 2020: p. 1359105320944991.

9. Haslam, C., et al., The new psychology of health: Unlocking the social cure. 2018: Routledge.

10. Heijnders, M.L. and J.J. Meijs, ’Welzijn op Recept’ (Social Prescribing): a helping hand in re-establishing social contacts - an explorative qualitative study. Primary Health Care Research & Development, 2018. 19(3): p. 223–231.

11. Zurynski, Y., A. Vedovi, and K.-l. Smith. Social prescribing: a rapid literature review to inform primary care policy in Australia. in Consumers’ Health Forum of Australia. 2020.

12. Jensen, A., et al., Arts on prescription in Scandinavia: a review of current practice and future possibilities. Perspectives in public health, 2017. 137(5): p. 268–274.

13. Baska, A., et al., Social prescribing and lifestyle medicine—a remedy to chronic health problems? International Journal of Environmental Research and Public Health, 2021. 18(19): p. 10096.

14. Bickerdike, L., et al., Social prescribing: less rhetoric and more reality. A systematic review of the evidence. BMJ open, 2017. 7(4): p. e013384.

15. Pilkington, K., M. Loef, and M. Polley, Searching for real-world effectiveness of health care innovations: scoping study of social prescribing for diabetes. Journal of medical Internet research, 2017. 19(2): p. e6431.

16. Kimberlee, R., Developing a social prescribing approach for Bristol. Bristol CCG, 2013.

17. Brettell, M., C. Fenton, and E. Foster, Linking Leeds: A Social Prescribing Service for Children and Young People. International Journal of Environmental Research and Public Health, 2022. 19(3): p. 1426.

18. Wildman, J. and J.M. Wildman, Evaluation of a Community Health Worker Social Prescribing Program Among UK Patients With Type 2 Diabetes. JAMA network open, 2021. 4(9): p. e2126236–e2126236.

19. Burns, J., et al., Linking families with pre-school children from healthcare services to community resources: a systematic review protocol. Systematic reviews, 2017. 6(1): p. 1–7.

20. Moher, D., et al., Preferred reporting items for systematic reviews and meta-analyses: the PRISMA statement. PLoS medicine, 2009. 6(7): p. e1000097.

21. Haddaway, N.R., et al., The role of Google Scholar in evidence reviews and its applicability to grey literature searching. PloS one, 2015. 10(9): p. e0138237.

22. Heijnders, M. and J.J. Meijs, Handboek Welzijn op Recept: Zorg en welzijn maken samen het verschil. 2019: Springer Nature.

23. Van de Schoot, R., et al., ASReview: Active learning for systematic reviews (Version v0.19). Zenodo. Retrieved from, 2020. 10.

24. van de Schoot, R., et al., An open source machine learning framework for efficient and transparent systematic reviews. Nature Machine Intelligence, 2021. 3(2): p. 125–133.

25. Ferdinands, G., et al., Active learning for screening prioritization in systematic reviews-A simulation study. 2020.

26. Yu, Z. and T. Menzies, Fast2: An intelligent assistant for finding relevant papers. Expert Systems with Applications, 2019. 120: p. 57–71.

27. Higgins, J.P., et al., Cochrane handbook for systematic reviews of interventions. 2019: John Wiley & Sons.

28. Michie, S., et al., Making psychological theory useful for implementing evidence based practice: a consensus approach. BMJ Quality & Safety, 2005. 14(1): p. 26–33.

29. Hong, Q.N., et al., The Mixed Methods Appraisal Tool (MMAT) version 2018 for information professionals and researchers. Education for information, 2018. 34(4): p. 285–291.

30. Sterne, J.A., et al., RoB 2: a revised tool for assessing risk of bias in randomised trials. bmj, 2019. 366.

31. Sterne, J.A., et al., ROBINS-I: a tool for assessing risk of bias in non-randomised studies of interventions. bmj, 2016. 355.

32. Ogilvie, D., et al., The harvest plot: a method for synthesising evidence about the differential effects of interventions. BMC medical research methodology, 2008. 8(1): p. 1–7.

33. RStudio Team, RStudio: Integrated Development Environment for R. 2022: RStudio, PBC, Boston, MA.

34. Aggar, C., et al., Social Prescribing for Individuals Living with Mental Illness in an Australian Community Setting: A Pilot Study. Community Mental Health Journal, 2021. 57(1): p. 189–195.

35. Crone, D.M., et al., ’Artlift’ arts-on-referral intervention in UK primary care: updated findings from an ongoing observational study. European Journal of Public Health, 2018. 28(3): p. 404–409.

36. Farenden, C., et al., Community navigation in Brighton & Hove. Evaluation of a social prescribing pilot. Hove: Brighton & Hove Impetus, 2015.

37. Holt, N.J., Tracking momentary experience in the evaluation of arts-on-prescription services: using mood changes during art workshops to predict global wellbeing change. Perspectives in Public Health, 2020. 140(5): p. 270–276.

38. Kim, J.E., et al., Effects of social prescribing pilot project for the elderly in rural area of South Korea during COVID-19 pandemic. Health Science Reports, 2021. 4(3): p. e320.

39. Loftus, A.M., F. McCauley, and M.O. McCarron, Impact of social prescribing on general practice workload and polypharmacy. Public Health, 2017. 148: p. 96–101.

40. Maughan, D.L., et al., Primary-care-based social prescribing for mental health: an analysis of financial and environmental sustainability. Primary Health Care Research & Development, 2016. 17(2): p. 114–21.

41. Mercer, S.W., et al., Effectiveness of Community-Links Practitioners in Areas of High Socioeconomic Deprivation. Annals of Family Medicine, 2019. 17(6): p. 518–525.

42. Morton, L., M. Ferguson, and F. Baty, Improving wellbeing and self-efficacy by social prescription. Public health, 2015. 3(129): p. 286–289.

43. Pescheny, J.V., et al., The impact of the Luton social prescribing programme on mental well-being: a quantitative before-and-after study. Journal of Public Health, 2021. 43(1): p. e69–e76.

44. Pescheny, J.V., et al., The impact of the Luton social prescribing programme on energy expenditure: a quantitative before-and-after study. BMJ Open, 2019. 9(6): p. e026862.

45. Sumner, R.C., et al., Factors associated with attendance, engagement and wellbeing change in an arts on prescription intervention. Journal of Public Health, 2020. 42(1): p. e88–e95.

46. Sumner, R.C., et al., Arts on prescription: observed changes in anxiety, depression, and well-being across referral cycles. Public Health, 2021. 192: p. 49–55.

47. Lynch, M. and C.R. Jones, Social prescribing for frequent attenders in primary care: An economic analysis. Frontiers in Public Health, 2022: p. 3580.

48. Pomp, M., Effectstudy Welbeing on Prescription [Effectstudie Welzijn op Recept]. 2015.

49. Bhatti, S., et al., Using self-determination theory to understand the social prescribing process: a qualitative study. Bjgp Open, 2021. 5(2).

50. Crone, D., et al., ‘It helps me make sense of the world’: the role of an art intervention for promoting health and wellbeing in primary care—perspectives of patients, health professionals and artists. Journal of Public Health, 2012. 20(5): p. 519–524.

51. Hanlon, P., et al., Does Self-Determination Theory help explain the impact of social prescribing? A qualitative analysis of patients’ experiences of the Glasgow ‘Deep-End’ Community Links Worker Intervention. Chronic Illness, 2021. 17(3): p. 173–188.

52. Payne, K., E. Walton, and C. Burton, Steps to benefit from social prescription: a qualitative interview study. British Journal of General Practice, 2020. 70(690): p. e36–e44.

53. Redmond, M., et al., ’Light in dark places’: exploring qualitative data from a longitudinal study using creative arts as a form of social prescribing. Arts & Health, 2019. 11(3): p. 232–245.

54. Simpson, S., et al., Supporting access to activities to enhance well-being and reduce social isolation in people living with motor neurone disease. Health & Social Care in the Community, 2020. 28(6): p. 2282–2289.

55. Carnes, D., et al., The impact of a social prescribing service on patients in primary care: a mixed methods evaluation. BMC Health Services Research, 2017. 17(1): p. 835.

56. Bertotti, M., et al., A realist evaluation of social prescribing: an exploration into the context and mechanisms underpinning a pathway linking primary care with the voluntary sector. Primary Health Care Research & Development, 2018. 19(3): p. 232–245.

57. Bertotti, M., C. Frostick, and O. Temirov, An evaluation of Social Prescribing in the London Borough of Redbridge: final evaluation report. 2020.

58. Bertotti, M., et al., The social prescribing service in the London Borough of Waltham Forest: final evaluation report. 2017.

59. Dayson, C., Evaluating social innovations and their contribution to social value: the benefits of a ‘blended value’ approach. Policy and Politics, 2017. 45(3): p. 395–411.

60. Dayson, C. and E. Bennett, Evaluation of Doncaster Social Prescribing Service: understanding outcomes and impact. 2016.

61. Dayson, C. and E. Bennett, Evaluation of the Rotherham mental health social prescribing pilot. 2015.

62. Envoy Partnership, Self-Care social prescribing. 2018, Kensington & Chelsea Social Council and NHS West London Clinical Commissioning Group: London.

63. Foster, A., et al., Impact of social prescribing to address loneliness: A mixed methods evaluation of a national social prescribing programme. Health & Social Care in the Community, 2021. 29(5): p. 1439–1449.

64. Howarth, M., et al., Social prescribing: a ‘natural’ community-based solution. British Journal of Community Nursing, 2020. 25(6): p. 294–298.

65. Jones, C. and M. Lynch, Spice time credits social prescribing pilot evaluation. Final Report, 2019.

66. Jones, C. and M. Lynch, Grow well social prescribing pilot evaluation. Final Report, 2020.

67. Kimberlee, R., Gloucestershire clinical commissioning group’s social prescribing service: evaluation report. 2016.

68. Kimberlee, R., et al., Measuring the economic impact of the wellspring healthy living centre’s social prescribing wellbeing programme for low level mental health issues encountered by GP services. Project Report. South West Forum, UK. 2013. POV_ Final_ Report_ March_, 2014.

69. Mulligan, K., et al., Social prescribing in Ontario, final report. Toronto: Alliance for Healthier Communities, 2020.

70. Palmer, D., et al., Social prescribing in Bexley: pilot evaluation report. 2017: Mind in Bexley.

71. Polley, M., H. Seers, and A. Fixsen, Evaluation report of the social prescribing demonstrator site in Shropshire–Final Report.University of Westminster, 2019.

72. Poulos, R.G., et al., Arts on prescription for community-dwelling older people with a range of health and wellness needs. Health & Social Care in the Community, 2019. 27(2): p. 483–492.

73. van de Venter, E. and A.M. Buller, Arts on referral interventions: a mixed-methods study investigating factors associated with differential changes in mental well-being. Journal of Public Health, 2015. 37(1): p. 143–150.

74. Vogelpoel, N. and K. Jarrold, Social prescription and the role of participatory arts programmes for older people with sensory impairments. Journal of Integrated Care, 2014. 22(2): p. 39-+.

75. Woodall, J., et al., Understanding the effectiveness and mechanisms of a social prescribing service: a mixed method analysis. BMC Health Services Research, 2018. 18(1): p. 604.

76. Potter S, Arts on Prescription 2014-15: Evaluation report. 2015.

77. Hughes, S., et al., Understanding well-being outcomes in primary care arts on referral interventions: a mixed method study. European Journal for Person Centered Healthcare, 2019. 7(3): p. 1768.

78. Friedli, L., M. Themessl-Huber, and M. Butchart, Evaluation of Dundee equally well sources of support: social prescribing in Maryfield. Evaluation report four, 2012.

79. Vidovic, D., G.Y. Reinhardt, and C. Hammerton, Can social prescribing foster individual and community well-being? A systematic review of the evidence. International journal of environmental research and public health, 2021. 18(10): p. 5276.

80. Costa, A., et al., Effectiveness of social prescribing programs in the primary health-care context: a systematic literature review. Sustainability, 2021. 13(5): p. 2731.

81. Cooper, M., et al., Effectiveness and active ingredients of social prescribing interventions targeting mental health: a systematic review. BMJ open, 2022. 12(7): p. e060214.

82. Napierala, H., et al., Social Prescribing: Systematic Review of the Effectiveness of Psychosocial Community Referral Interventions in Primary Care. International Journal of Integrated Care, 2022. 22(3).

83. Hill, P.L., et al., The value of a purposeful life: Sense of purpose predicts greater income and net worth. Journal of research in personality, 2016. 65: p. 38–42.

84. Pescheny, J.V., Y. Pappas, and G. Randhawa, Facilitators and barriers of implementing and delivering social prescribing services: a systematic review. BMC Health Services Research, 2018. 18(1): p. 1–14.

85. Brandling, J. and W. House, Social prescribing in general practice: adding meaning to medicine. British Journal of General Practice, 2009. 59(563): p. 454–456.

86. Tabaei-Aghdaei, Z., J.R. McColl-Kennedy, and L.V. Coote, Goal setting and health-related outcomes in chronic diseases: a systematic review and meta-analysis of the literature from 2000 to 2020. Medical Care Research and Review, 2023. 80(2): p. 145–164.

87. Percival, A., et al., Systematic review of social prescribing and older adults: where to from here? Family Medicine and Community Health, 2022. 10(Suppl 1): p. e001829.

88. Chatterjee, H.J., et al., Non-clinical community interventions: a systematised review of social prescribing schemes. Arts & Health, 2018. 10(2): p. 97–123.

89. Husk, K., et al., Social prescribing: where is the evidence? 2019, British Journal of General Practice. p. 6–7.

90. Mohr, D.C., et al., Trials of intervention principles: evaluation methods for evolving behavioral intervention technologies. Journal of medical Internet research, 2015. 17(7): p. e4391.

91. Williams, D.R. and S.A. Mohammed, Discrimination and racial disparities in health: evidence and needed research. Journal of behavioral medicine, 2009. 32: p. 20–47.

92. Netto, G., et al., How can health promotion interventions be adapted for minority ethnic communities? Five principles for guiding the development of behavioural interventions. Health promotion international, 2010. 25(2): p. 248–257.

93. UK Government. Social prescribing: applying All Our Health. 2019 Januari 27th, 2022 [cited 2023 April 19th]; Available from: https://www.gov.uk/government/publications/social-prescribing-applying-all-our-health/social-prescribing-applying-all-our-health.

94. Moscrop, A., Social prescribing is no remedy for health inequalities. 2023, British Medical Journal Publishing Group.

95. Landelijk Netwerk Welzijn op Recept. Welzijn op Recept. 2021 [cited 2023 April 19th]; Available from: www.welzijnoprecept.n;/.

96. Sandhu, S., et al., Intervention components of link worker social prescribing programmes: a scoping review. Health & Social Care in the Community, 2022.

