## AppendixB_SearchStrategy for "Intervention Characteristics and Mechanisms and their Relationship with the Influence of Social Prescribing: a Systematic Review"

**Appendix B**. Search strategy

Databases:

**MEDLINE via OVID**

<http://gateway.ovid.com/ovidweb.cgi?T=JS&MODE=ovid&NEWS=n&PAGE=main&D=medall>

**(**("social prescribing".mp OR "social prescri*".mp OR "community prescribing".mp OR "community prescri*".mp OR "community referral".mp OR "community referrals".mp OR "community referr*".mp OR "social referral".mp OR "social referrals".mp OR "social referr*".mp OR "linking scheme".mp OR "linking schemes".mp OR "referral scheme".mp OR "referral schemes".mp) **OR** ("social" **ADJ1** "prescri*").mp OR ("community" **ADJ1** "prescri*").mp **OR** (("social intervention".mp OR "social interventions".mp OR "welfare intervention".mp OR "welfare interventions".mp) AND ("link worker".mp OR "link workers".mp OR "link work*".mp OR "community navigators".mp OR "community navigator".mp OR "community liaisons".mp OR "community liaison".mp OR "well being coach".mp OR "wellbeing coach".mp OR "well being coaches".mp OR "wellbeing coaches".mp OR "welfare coordinators".mp OR "welfare coordinator".mp) AND (exp "Primary Health Care"/ OR "Primary Health Care".mp OR "Primary Healthcare".mp OR "Primary Care".mp OR exp "General Practice"/ OR "Family Practice".mp OR "General Practice".mp OR exp "General Practitioners"/ OR "General Practitioner".mp OR "General Practitioners".mp OR "GP".ti,ab OR exp "Physicians, Family"/ OR "Family Physician".mp OR "Family Physicians".mp OR exp "Physical Therapists"/ OR "Physical Therapist".mp OR "Physical Therapists".mp OR "Physiotherapist".mp OR "Physiotherapists".mp OR exp "Physical Therapy Specialty"/ OR exp "Physical Therapy Modalities"/ OR exp "Social Workers"/ OR "Social Worker".mp OR "Social Workers".mp OR exp "Social Work"/ OR "Social Work".mp OR "health centres".mp OR "health centre".mp OR "health centers".mp OR "health center".mp OR "health care centres".mp OR "health care centre".mp OR "health care centers".mp OR "health care center".mp OR "healthcare centres".mp OR "healthcare centre".mp OR "healthcare centers".mp OR "healthcare center".mp OR "health services".mp OR "health service".mp OR "health care services".mp OR "health care service".mp OR "healthcare services".mp OR "healthcare service".mp OR "social care".mp)) **OR** "Arts on Prescription".mp OR "Books on Prescription".mp OR "Education on Prescription".mp OR "exercise on prescription".mp OR "exercise referral".mp **OR** ((("referral" OR "referr*" OR "prescription" OR "prescrib*" OR "prescrip*") **ADJ1** ("adult learning" OR "aqua therapy" OR "art" OR "art therapy" OR "arts" OR "befriending" OR "bibliotherap*" OR "bibliotherapy" OR "books" OR "community education group" OR "community education groups" OR "computerised cbt" OR "computerised cognitive behavioural therapy" OR "computerized cbt" OR "computerized cognitive behavioural therapy" OR "creativity" OR "cycling" OR "dance" OR "dance class" OR "dance classes" OR "dancing" OR "education" OR "exercise" OR "exercise class" OR "exercise classes" OR "exercising " OR "fishing" OR "fishing club" OR "fishing clubs" OR "gardening" OR "gardening club" OR "gardening clubs" OR "green gym" OR "group activities" OR "group activity" OR "guided walk" OR "guided walking" OR "guided walks" OR "gym" OR "gym-based activities" OR "gym-based activity" OR "gymnastic" OR "gymnastics" OR "health walk" OR "health walks" OR "knit club" OR "knit clubs" OR "knitting" OR "knitting club" OR "knitting clubs" OR "learning" OR "learning new skill" OR "learning new skills" OR "mutual aid" OR "natter club" OR "natter clubs" OR "physical activities" OR "physical activity" OR "self-help group" OR "self-help groups" OR "self-help reading" OR "signposting guidance" OR "signposting information" OR "supported education" OR "supported employment" OR "swimming" OR "team sport" OR "team sports" OR "time bank" OR "time banks" OR "volunteer" OR "volunteering" OR "volunteers" OR "walk" OR "walking" OR "walks").mp) AND (exp "Primary Health Care"/ OR "Primary Health Care".mp OR "Primary Healthcare".mp OR "Primary Care".mp OR exp "General Practice"/ OR "Family Practice".mp OR "General Practice".mp OR exp "General Practitioners"/ OR "General Practitioner".mp OR "General Practitioners".mp OR "GP".ti,ab OR exp "Physicians, Family"/ OR "Family Physician".mp OR "Family Physicians".mp OR exp "Physical Therapists"/ OR "Physical Therapist".mp OR "Physical Therapists".mp OR "Physiotherapist".mp OR "Physiotherapists".mp OR exp "Physical Therapy Specialty"/ OR exp "Physical Therapy Modalities"/ OR exp "Social Workers"/ OR "Social Worker".mp OR "Social Workers".mp OR exp "Social Work"/ OR "Social Work".mp OR "health centres".mp OR "health centre".mp OR "health centers".mp OR "health center".mp OR "health care centres".mp OR "health care centre".mp OR "health care centers".mp OR "health care center".mp OR "healthcare centres".mp OR "healthcare centre".mp OR "healthcare centers".mp OR "healthcare center".mp OR "health services".mp OR "health service".mp OR "health care services".mp OR "health care service".mp OR "healthcare services".mp OR "healthcare service".mp OR "social care".mp)) **OR** (("referral" OR "referr*" OR "prescription" OR "prescrib*" OR "prescrip*") **ADJ1** ("adult learning" OR "aqua therapy" OR "art" OR "art therapy" OR "arts" OR "befriending" OR "bibliotherap*" OR "bibliotherapy" OR "books" OR "community education group" OR "community education groups" OR "computerised cbt" OR "computerised cognitive behavioural therapy" OR "computerized cbt" OR "computerized cognitive behavioural therapy" OR "creativity" OR "cycling" OR "dance" OR "dance class" OR "dance classes" OR "dancing" OR "education" OR "exercise" OR "exercise class" OR "exercise classes" OR "exercising " OR "fishing" OR "fishing club" OR "fishing clubs" OR "gardening" OR "gardening club" OR "gardening clubs" OR "green gym" OR "group activities" OR "group activity" OR "guided walk" OR "guided walking" OR "guided walks" OR "gym" OR "gym-based activities" OR "gym-based activity" OR "gymnastic" OR "gymnastics" OR "health walk" OR "health walks" OR "knit club" OR "knit clubs" OR "knitting" OR "knitting club" OR "knitting clubs" OR "learning" OR "learning new skill" OR "learning new skills" OR "mutual aid" OR "natter club" OR "natter clubs" OR "physical activities" OR "physical activity" OR "self-help group" OR "self-help groups" OR "self-help reading" OR "signposting guidance" OR "signposting information" OR "supported education" OR "supported employment" OR "swimming" OR "team sport" OR "team sports" OR "time bank" OR "time banks" OR "volunteer" OR "volunteering" OR "volunteers" OR "walk" OR "walking" OR "walks")).ti**)**

AND (exp "review"/ OR "systematic review"/ OR "review".mp OR "overview".mp OR "review*".mp OR "overview*".mp OR "appraisal".mp OR "synthesis".mp)

**PubMed**

<http://www.ncbi.nlm.nih.gov/pubmed?otool=leiden>

**(**("social prescribing"[tw] OR "social prescri*"[tw] OR "community prescribing"[tw] OR "community prescri*"[tw] OR "community referral"[tw] OR "community referrals"[tw] OR "community referr*"[tw] OR "social referral"[tw] OR "social referrals"[tw] OR "social referr*"[tw] OR "linking scheme"[tw] OR "linking schemes"[tw] OR "referral scheme"[tw] OR "referral schemes"[tw]) OR (("social intervention"[tw] OR "social interventions"[tw] OR "welfare intervention"[tw] OR "welfare interventions"[tw]) AND ("link worker"[tw] OR "link workers"[tw] OR "link work*"[tw] OR "community navigators"[tw] OR "community navigator"[tw] OR "community liaisons"[tw] OR "community liaison"[tw] OR "well being coach"[tw] OR "wellbeing coach"[tw] OR "well being coaches"[tw] OR "wellbeing coaches"[tw] OR "welfare coordinators"[tw] OR "welfare coordinator"[tw]) AND ("Primary Health Care"[mesh] OR "Primary Health Care"[tw] OR "Primary Healthcare"[tw] OR "Primary Care"[tw] OR "General Practice"[mesh] OR "Family Practice"[tw] OR "General Practice"[tw] OR "General Practitioners"[mesh] OR "General Practitioner"[tw] OR "General Practitioners"[tw] OR "GP".ti,ab OR "Physicians, Family"[mesh] OR "Family Physician"[tw] OR "Family Physicians"[tw] OR "Physical Therapists"[mesh] OR "Physical Therapist"[tw] OR "Physical Therapists"[tw] OR "Physiotherapist"[tw] OR "Physiotherapists"[tw] OR "Physical Therapy Specialty"[mesh] OR "Physical Therapy Modalities"[mesh] OR "Social Workers"[mesh] OR "Social Worker"[tw] OR "Social Workers"[tw] OR "Social Work"[mesh] OR "Social Work"[tw] OR "health centres"[tw] OR "health centre"[tw] OR "health centers"[tw] OR "health center"[tw] OR "health care centres"[tw] OR "health care centre"[tw] OR "health care centers"[tw] OR "health care center"[tw] OR "healthcare centres"[tw] OR "healthcare centre"[tw] OR "healthcare centers"[tw] OR "healthcare center"[tw] OR "health services"[tw] OR "health service"[tw] OR "health care services"[tw] OR "health care service"[tw] OR "healthcare services"[tw] OR "healthcare service"[tw] OR "social care"[tw])) **OR** "Arts on Prescription"[tw] OR "Books on Prescription"[tw] OR "Education on Prescription"[tw] OR "exercise on prescription"[tw] OR "exercise referral"[tw] **OR** ((("referral"[ti] OR "referr*"[ti] OR "prescription"[ti] OR "prescrib*"[ti] OR "prescrip*"[ti]) **AND** ("adult learning"[ti] OR "aqua therapy"[ti] OR "art"[ti] OR "art therapy"[ti] OR "arts"[ti] OR "befriending"[ti] OR "bibliotherap*"[ti] OR "bibliotherapy"[ti] OR "books"[ti] OR "community education group"[ti] OR "community education groups"[ti] OR "computerised cbt"[ti] OR "computerised cognitive behavioural therapy"[ti] OR "computerized cbt"[ti] OR "computerized cognitive behavioural therapy"[ti] OR "creativity"[ti] OR "cycling"[ti] OR "dance"[ti] OR "dance class"[ti] OR "dance classes"[ti] OR "dancing"[ti] OR "education"[ti] OR "exercise"[ti] OR "exercise class"[ti] OR "exercise classes"[ti] OR "exercising "[ti] OR "fishing"[ti] OR "fishing club"[ti] OR "fishing clubs"[ti] OR "gardening"[ti] OR "gardening club"[ti] OR "gardening clubs"[ti] OR "green gym"[ti] OR "group activities"[ti] OR "group activity"[ti] OR "guided walk"[ti] OR "guided walking"[ti] OR "guided walks"[ti] OR "gym"[ti] OR "gym-based activities"[ti] OR "gym-based activity"[ti] OR "gymnastic"[ti] OR "gymnastics"[ti] OR "health walk"[ti] OR "health walks"[ti] OR "knit club"[ti] OR "knit clubs"[ti] OR "knitting"[ti] OR "knitting club"[ti] OR "knitting clubs"[ti] OR "learning"[ti] OR "learning new skill"[ti] OR "learning new skills"[ti] OR "mutual aid"[ti] OR "natter club"[ti] OR "natter clubs"[ti] OR "physical activities"[ti] OR "physical activity"[ti] OR "self-help group"[ti] OR "self-help groups"[ti] OR "self-help reading"[ti] OR "signposting guidance"[ti] OR "signposting information"[ti] OR "supported education"[ti] OR "supported employment"[ti] OR "swimming"[ti] OR "team sport"[ti] OR "team sports"[ti] OR "time bank"[ti] OR "time banks"[ti] OR "volunteer"[ti] OR "volunteering"[ti] OR "volunteers"[ti] OR "walk"[ti] OR "walking"[ti] OR "walks"[ti])) AND ("Primary Health Care"[mesh] OR "Primary Health Care"[tw] OR "Primary Healthcare"[tw] OR "Primary Care"[tw] OR "General Practice"[mesh] OR "Family Practice"[tw] OR "General Practice"[tw] OR "General Practitioners"[mesh] OR "General Practitioner"[tw] OR "General Practitioners"[tw] OR "GP".ti,ab OR "Physicians, Family"[mesh] OR "Family Physician"[tw] OR "Family Physicians"[tw] OR "Physical Therapists"[mesh] OR "Physical Therapist"[tw] OR "Physical Therapists"[tw] OR "Physiotherapist"[tw] OR "Physiotherapists"[tw] OR "Physical Therapy Specialty"[mesh] OR "Physical Therapy Modalities"[mesh] OR "Social Workers"[mesh] OR "Social Worker"[tw] OR "Social Workers"[tw] OR "Social Work"[mesh] OR "Social Work"[tw] OR "health centres"[tw] OR "health centre"[tw] OR "health centers"[tw] OR "health center"[tw] OR "health care centres"[tw] OR "health care centre"[tw] OR "health care centers"[tw] OR "health care center"[tw] OR "healthcare centres"[tw] OR "healthcare centre"[tw] OR "healthcare centers"[tw] OR "healthcare center"[tw] OR "health services"[tw] OR "health service"[tw] OR "health care services"[tw] OR "health care service"[tw] OR "healthcare services"[tw] OR "healthcare service"[tw] OR "social care"[tw]))**)**

AND ("review"[pt] OR "systematic review"[pt] OR "systematic"[sb] OR "review"[tw] OR "overview"[tw] OR "review*"[tw] OR "overview*"[tw] OR "appraisal"[tw] OR "synthesis"[tw] OR "Meta-Analysis"[pt] OR "Meta-Analy*"[tw] OR "Metaanaly*"[tw])

**Embase**

<http://ovidsp.ovid.com/ovidweb.cgi?T=JS&PAGE=main&MODE=ovid&D=oemezd>

**(**("social prescribing".ti,ab OR "social prescri*".ti,ab OR "community prescribing".ti,ab OR "community prescri*".ti,ab OR "community referral".ti,ab OR "community referrals".ti,ab OR "community referr*".ti,ab OR "social referral".ti,ab OR "social referrals".ti,ab OR "social referr*".ti,ab OR "linking scheme".ti,ab OR "linking schemes".ti,ab OR "referral scheme".ti,ab OR "referral schemes".ti,ab) **OR** ("social" **ADJ1** "prescri*").ti,ab OR ("community" **ADJ1** "prescri*").ti,ab **OR** (("social intervention".ti,ab OR "social interventions".ti,ab OR "welfare intervention".ti,ab OR "welfare interventions".ti,ab) AND ("link worker".ti,ab OR "link workers".ti,ab OR "link work*".ti,ab OR "community navigators".ti,ab OR "community navigator".ti,ab OR "community liaisons".ti,ab OR "community liaison".ti,ab OR "well being coach".ti,ab OR "wellbeing coach".ti,ab OR "well being coaches".ti,ab OR "wellbeing coaches".ti,ab OR "welfare coordinators".ti,ab OR "welfare coordinator".ti,ab) AND (exp *"Primary Health Care"/ OR "Primary Health Care".ti,ab OR "Primary Healthcare".ti,ab OR "Primary Care".ti,ab OR exp *"General Practice"/ OR "Family Practice".ti,ab OR "General Practice".ti,ab OR exp *"General Practitioner"/ OR "General Practitioner".ti,ab OR "General Practitioners".ti,ab OR "GP".ti,ab OR "Family Physician".ti,ab OR "Family Physicians".ti,ab OR exp *"Physiotherapist"/ OR "Physical Therapist".ti,ab OR "Physical Therapists".ti,ab OR "Physiotherapist".ti,ab OR "Physiotherapists".ti,ab OR exp *"Physiotherapy"/ OR exp *"Social Worker"/ OR "Social Worker".ti,ab OR "Social Workers".ti,ab OR exp *"Social Work"/ OR "Social Work".ti,ab OR exp *"health center"/ OR "health centres".ti,ab OR "health centre".ti,ab OR "health centers".ti,ab OR "health center".ti,ab OR "health care centres".ti,ab OR "health care centre".ti,ab OR "health care centers".ti,ab OR "health care center".ti,ab OR "healthcare centres".ti,ab OR "healthcare centre".ti,ab OR "healthcare centers".ti,ab OR "healthcare center".ti,ab OR "health services".ti,ab OR "health service".ti,ab OR "health care services".ti,ab OR "health care service".ti,ab OR "healthcare services".ti,ab OR "healthcare service".ti,ab OR "social care".ti,ab)) **OR** "Arts on Prescription".ti,ab OR "Books on Prescription".ti,ab OR "Education on Prescription".ti,ab OR "exercise on prescription".ti,ab OR "exercise referral".ti,ab **OR** ((("referral" OR "referr*" OR "prescription" OR "prescrib*" OR "prescrip*") **ADJ1** ("adult learning" OR "aqua therapy" OR "art" OR "art therapy" OR "arts" OR "befriending" OR "bibliotherap*" OR "bibliotherapy" OR "books" OR "community education group" OR "community education groups" OR "computerised cbt" OR "computerised cognitive behavioural therapy" OR "computerized cbt" OR "computerized cognitive behavioural therapy" OR "creativity" OR "cycling" OR "dance" OR "dance class" OR "dance classes" OR "dancing" OR "education" OR "exercise" OR "exercise class" OR "exercise classes" OR "exercising " OR "fishing" OR "fishing club" OR "fishing clubs" OR "gardening" OR "gardening club" OR "gardening clubs" OR "green gym" OR "group activities" OR "group activity" OR "guided walk" OR "guided walking" OR "guided walks" OR "gym" OR "gym-based activities" OR "gym-based activity" OR "gymnastic" OR "gymnastics" OR "health walk" OR "health walks" OR "knit club" OR "knit clubs" OR "knitting" OR "knitting club" OR "knitting clubs" OR "learning" OR "learning new skill" OR "learning new skills" OR "mutual aid" OR "natter club" OR "natter clubs" OR "physical activities" OR "physical activity" OR "self-help group" OR "self-help groups" OR "self-help reading" OR "signposting guidance" OR "signposting information" OR "supported education" OR "supported employment" OR "swimming" OR "team sport" OR "team sports" OR "time bank" OR "time banks" OR "volunteer" OR "volunteering" OR "volunteers" OR "walk" OR "walking" OR "walks").ti,ab) AND (exp *"Primary Health Care"/ OR "Primary Health Care".ti,ab OR "Primary Healthcare".ti,ab OR "Primary Care".ti,ab OR exp *"General Practice"/ OR "Family Practice".ti,ab OR "General Practice".ti,ab OR exp *"General Practitioner"/ OR "General Practitioner".ti,ab OR "General Practitioners".ti,ab OR "GP".ti,ab OR "Family Physician".ti,ab OR "Family Physicians".ti,ab OR exp *"Physiotherapist"/ OR "Physical Therapist".ti,ab OR "Physical Therapists".ti,ab OR "Physiotherapist".ti,ab OR "Physiotherapists".ti,ab OR exp *"Physiotherapy"/ OR exp *"Social Worker"/ OR "Social Worker".ti,ab OR "Social Workers".ti,ab OR exp *"Social Work"/ OR "Social Work".ti,ab OR exp *"health center"/ OR "health centres".ti,ab OR "health centre".ti,ab OR "health centers".ti,ab OR "health center".ti,ab OR "health care centres".ti,ab OR "health care centre".ti,ab OR "health care centers".ti,ab OR "health care center".ti,ab OR "healthcare centres".ti,ab OR "healthcare centre".ti,ab OR "healthcare centers".ti,ab OR "healthcare center".ti,ab OR "health services".ti,ab OR "health service".ti,ab OR "health care services".ti,ab OR "health care service".ti,ab OR "healthcare services".ti,ab OR "healthcare service".ti,ab OR "social care".ti,ab)) **OR** (("referral" OR "referr*" OR "prescription" OR "prescrib*" OR "prescrip*") **ADJ1** ("adult learning" OR "aqua therapy" OR "art" OR "art therapy" OR "arts" OR "befriending" OR "bibliotherap*" OR "bibliotherapy" OR "books" OR "community education group" OR "community education groups" OR "computerised cbt" OR "computerised cognitive behavioural therapy" OR "computerized cbt" OR "computerized cognitive behavioural therapy" OR "creativity" OR "cycling" OR "dance" OR "dance class" OR "dance classes" OR "dancing" OR "education" OR "exercise" OR "exercise class" OR "exercise classes" OR "exercising " OR "fishing" OR "fishing club" OR "fishing clubs" OR "gardening" OR "gardening club" OR "gardening clubs" OR "green gym" OR "group activities" OR "group activity" OR "guided walk" OR "guided walking" OR "guided walks" OR "gym" OR "gym-based activities" OR "gym-based activity" OR "gymnastic" OR "gymnastics" OR "health walk" OR "health walks" OR "knit club" OR "knit clubs" OR "knitting" OR "knitting club" OR "knitting clubs" OR "learning" OR "learning new skill" OR "learning new skills" OR "mutual aid" OR "natter club" OR "natter clubs" OR "physical activities" OR "physical activity" OR "self-help group" OR "self-help groups" OR "self-help reading" OR "signposting guidance" OR "signposting information" OR "supported education" OR "supported employment" OR "swimming" OR "team sport" OR "team sports" OR "time bank" OR "time banks" OR "volunteer" OR "volunteering" OR "volunteers" OR "walk" OR "walking" OR "walks")).ti**)**

AND (exp "review"/ OR "systematic review"/ OR "review".mp OR "overview".mp OR "review*".mp OR "overview*".mp OR "appraisal".mp OR "synthesis".mp)

NOT conference review.pt

NOT (conference review or conference abstract).pt

AND (conference abstract).pt

**Web of Science**

<http://isiknowledge.com/wos>

**(**ti=("social prescribing" OR "social prescri*" OR "community prescribing" OR "community prescri*" OR "community referral" OR "community referrals" OR "community referr*" OR "social referral" OR "social referrals" OR "social referr*" OR "linking scheme" OR "linking schemes" OR "referral scheme" OR "referral schemes") OR ab=("social prescribing" OR "social prescri*" OR "community prescribing" OR "community prescri*" OR "community referral" OR "community referrals" OR "community referr*" OR "social referral" OR "social referrals" OR "social referr*" OR "linking scheme" OR "linking schemes" OR "referral scheme" OR "referral schemes") **OR** ti=(("social" **NEAR/1** "prescri*") OR ("community" **NEAR/1** "prescri*")) **OR** ts=(("social intervention" OR "social interventions" OR "welfare intervention" OR "welfare interventions") AND ("link worker" OR "link workers" OR "link work*" OR "community navigators" OR "community navigator" OR "community liaisons" OR "community liaison" OR "well being coach" OR "wellbeing coach" OR "well being coaches" OR "wellbeing coaches" OR "welfare coordinators" OR "welfare coordinator") AND ("Primary Health Care" OR "Primary Health Care" OR "Primary Healthcare" OR "Primary Care" OR "General Practice" OR "Family Practice" OR "General Practice" OR "General Practitioner" OR "General Practitioner" OR "General Practitioners" OR "GP" OR "Family Physician" OR "Family Physicians" OR "Physiotherapist" OR "Physical Therapist" OR "Physical Therapists" OR "Physiotherapist" OR "Physiotherapists" OR "Physiotherapy" OR "Social Worker" OR "Social Worker" OR "Social Workers" OR "Social Work" OR "Social Work" OR "health center" OR "health centres" OR "health centre" OR "health centers" OR "health center" OR "health care centres" OR "health care centre" OR "health care centers" OR "health care center" OR "healthcare centres" OR "healthcare centre" OR "healthcare centers" OR "healthcare center" OR "health services" OR "health service" OR "health care services" OR "health care service" OR "healthcare services" OR "healthcare service" OR "social care")) **OR** ti=("Arts on Prescription" OR "Books on Prescription" OR "Education on Prescription" OR "exercise on prescription" OR "exercise referral") OR ab=("Arts on Prescription" OR "Books on Prescription" OR "Education on Prescription" OR "exercise on prescription" OR "exercise referral") **OR** ti=((("referral" OR "referr*" OR "prescription" OR "prescrib*" OR "prescrip*") **NEAR/1** ("adult learning" OR "aqua therapy" OR "art" OR "art therapy" OR "arts" OR "befriending" OR "bibliotherap*" OR "bibliotherapy" OR "books" OR "community education group" OR "community education groups" OR "computerised cbt" OR "computerised cognitive behavioural therapy" OR "computerized cbt" OR "computerized cognitive behavioural therapy" OR "creativity" OR "cycling" OR "dance" OR "dance class" OR "dance classes" OR "dancing" OR "education" OR "exercise" OR "exercise class" OR "exercise classes" OR "exercising " OR "fishing" OR "fishing club" OR "fishing clubs" OR "gardening" OR "gardening club" OR "gardening clubs" OR "green gym" OR "group activities" OR "group activity" OR "guided walk" OR "guided walking" OR "guided walks" OR "gym" OR "gym-based activities" OR "gym-based activity" OR "gymnastic" OR "gymnastics" OR "health walk" OR "health walks" OR "knit club" OR "knit clubs" OR "knitting" OR "knitting club" OR "knitting clubs" OR "learning" OR "learning new skill" OR "learning new skills" OR "mutual aid" OR "natter club" OR "natter clubs" OR "physical activities" OR "physical activity" OR "self-help group" OR "self-help groups" OR "self-help reading" OR "signposting guidance" OR "signposting information" OR "supported education" OR "supported employment" OR "swimming" OR "team sport" OR "team sports" OR "time bank" OR "time banks" OR "volunteer" OR "volunteering" OR "volunteers" OR "walk" OR "walking" OR "walks")) AND ("Primary Health Care" OR "Primary Health Care" OR "Primary Healthcare" OR "Primary Care" OR "General Practice" OR "Family Practice" OR "General Practice" OR "General Practitioner" OR "General Practitioner" OR "General Practitioners" OR "GP" OR "Family Physician" OR "Family Physicians" OR "Physiotherapist" OR "Physical Therapist" OR "Physical Therapists" OR "Physiotherapist" OR "Physiotherapists" OR "Physiotherapy" OR "Social Worker" OR "Social Worker" OR "Social Workers" OR "Social Work" OR "Social Work" OR "health center" OR "health centres" OR "health centre" OR "health centers" OR "health center" OR "health care centres" OR "health care centre" OR "health care centers" OR "health care center" OR "healthcare centres" OR "healthcare centre" OR "healthcare centers" OR "healthcare center" OR "health services" OR "health service" OR "health care services" OR "health care service" OR "healthcare services" OR "healthcare service" OR "social care")) **OR** ti=(("referral" OR "referral*" OR "prescription" OR "prescrib*" OR "prescrip*") **NEAR/1** ("adult learning" OR "aqua therapy" OR "art" OR "art therapy" OR "arts" OR "befriending" OR "bibliotherap*" OR "bibliotherapy" OR "books" OR "community education group" OR "community education groups" OR "computerised cbt" OR "computerised cognitive behavioural therapy" OR "computerized cbt" OR "computerized cognitive behavioural therapy" OR "creativity" OR "cycling" OR "dance" OR "dance class" OR "dance classes" OR "dancing" OR "education" OR "exercise" OR "exercise class" OR "exercise classes" OR "exercising " OR "fishing" OR "fishing club" OR "fishing clubs" OR "gardening" OR "gardening club" OR "gardening clubs" OR "green gym" OR "group activities" OR "group activity" OR "guided walk" OR "guided walking" OR "guided walks" OR "gym" OR "gym-based activities" OR "gym-based activity" OR "gymnastic" OR "gymnastics" OR "health walk" OR "health walks" OR "knit club" OR "knit clubs" OR "knitting" OR "knitting club" OR "knitting clubs" OR "learning" OR "learning new skill" OR "learning new skills" OR "mutual aid" OR "natter club" OR "natter clubs" OR "physical activities" OR "physical activity" OR "self-help group" OR "self-help groups" OR "self-help reading" OR "signposting guidance" OR "signposting information" OR "supported education" OR "supported employment" OR "swimming" OR "team sport" OR "team sports" OR "time bank" OR "time banks" OR "volunteer" OR "volunteering" OR "volunteers" OR "walk" OR "walking" OR "walks"))**)**

AND TS=("review" OR "systematic review" OR "review" OR "overview" OR "review*" OR "overview*" OR "appraisal" OR "synthesis")

**Cochrane**

<https://www.cochranelibrary.com/advanced-search/search-manager>

**(**

("social prescribing" OR "social prescri*" OR "community prescribing" OR "community prescri*" OR "community referral" OR "community referrals" OR "community referr*" OR "social referral" OR "social referrals" OR "social referr*" OR "linking scheme" OR "linking schemes" OR "referral scheme" OR "referral schemes"):ti,ab,kw

**OR**

(("social" **NEAR/1** "prescribing") OR ("community" **NEAR/1** "prescribing")):ti,ab,kw

**OR**

(("social" **NEAR/1** "prescription") OR ("community" **NEAR/1** "prescription")):ti,ab,kw

**OR**

(("social intervention" OR "social interventions" OR "welfare intervention" OR "welfare interventions") AND ("link worker" OR "link workers" OR "link work*" OR "community navigators" OR "community navigator" OR "community liaisons" OR "community liaison" OR "well being coach" OR "wellbeing coach" OR "well being coaches" OR "wellbeing coaches" OR "welfare coordinators" OR "welfare coordinator") AND ("Primary Health Care" OR "Primary Health Care" OR "Primary Healthcare" OR "Primary Care" OR "General Practice" OR "Family Practice" OR "General Practice" OR "General Practitioner" OR "General Practitioner" OR "General Practitioners" OR "GP" OR "Family Physician" OR "Family Physicians" OR "Physiotherapist" OR "Physical Therapist" OR "Physical Therapists" OR "Physiotherapist" OR "Physiotherapists" OR "Physiotherapy" OR "Social Worker" OR "Social Worker" OR "Social Workers" OR "Social Work" OR "Social Work" OR "health center" OR "health centres" OR "health centre" OR "health centers" OR "health center" OR "health care centres" OR "health care centre" OR "health care centers" OR "health care center" OR "healthcare centres" OR "healthcare centre" OR "healthcare centers" OR "healthcare center" OR "health services" OR "health service" OR "health care services" OR "health care service" OR "healthcare services" OR "healthcare service" OR "social care")):ti,ab,kw

**OR**

("Arts on Prescription" OR "Books on Prescription" OR "Education on Prescription" OR "exercise on prescription" OR "exercise referral"):ti,kw

**OR**

(("Arts on Prescription" OR "Books on Prescription" OR "Education on Prescription" OR "exercise on prescription" OR "exercise referral") AND ("Primary Health Care" OR "Primary Health Care" OR "Primary Healthcare" OR "Primary Care" OR "General Practice" OR "Family Practice" OR "General Practice" OR "General Practitioner" OR "General Practitioner" OR "General Practitioners" OR "GP" OR "Family Physician" OR "Family Physicians" OR "Physiotherapist" OR "Physical Therapist" OR "Physical Therapists" OR "Physiotherapist" OR "Physiotherapists" OR "Physiotherapy" OR "Social Worker" OR "Social Worker" OR "Social Workers" OR "Social Work" OR "Social Work" OR "health center" OR "health centres" OR "health centre" OR "health centers" OR "health center" OR "health care centres" OR "health care centre" OR "health care centers" OR "health care center" OR "healthcare centres" OR "healthcare centre" OR "healthcare centers" OR "healthcare center" OR "health services" OR "health service" OR "health care services" OR "health care service" OR "healthcare services" OR "healthcare service" OR "social care")):ti,ab,kw

**OR**

((("referral" OR "referr*" OR "prescription" OR "prescrib*" OR "prescrip*") **NEAR/1** ("adult learning" OR "aqua therapy" OR "art" OR "art therapy" OR "arts" OR "befriending" OR "bibliotherap*" OR "bibliotherapy" OR "books" OR "community education group" OR "community education groups" OR "computerised cbt" OR "computerised cognitive behavioural therapy" OR "computerized cbt" OR "computerized cognitive behavioural therapy" OR "creativity" OR "cycling" OR "dance" OR "dance class" OR "dance classes" OR "dancing" OR "education" OR "exercise" OR "exercise class" OR "exercise classes" OR "exercising " OR "fishing" OR "fishing club" OR "fishing clubs" OR "gardening" OR "gardening club" OR "gardening clubs" OR "green gym" OR "group activities" OR "group activity" OR "guided walk" OR "guided walking" OR "guided walks" OR "gym" OR "gym-based activities" OR "gym-based activity" OR "gymnastic" OR "gymnastics" OR "health walk" OR "health walks" OR "knit club" OR "knit clubs" OR "knitting" OR "knitting club" OR "knitting clubs" OR "learning" OR "learning new skill" OR "learning new skills" OR "mutual aid" OR "natter club" OR "natter clubs" OR "physical activities" OR "physical activity" OR "self-help group" OR "self-help groups" OR "self-help reading" OR "signposting guidance" OR "signposting information" OR "supported education" OR "supported employment" OR "swimming" OR "team sport" OR "team sports" OR "time bank" OR "time banks" OR "volunteer" OR "volunteering" OR "volunteers" OR "walk" OR "walking" OR "walks")) AND ("Primary Health Care" OR "Primary Health Care" OR "Primary Healthcare" OR "Primary Care" OR "General Practice" OR "Family Practice" OR "General Practice" OR "General Practitioner" OR "General Practitioner" OR "General Practitioners" OR "GP" OR "Family Physician" OR "Family Physicians" OR "Physiotherapist" OR "Physical Therapist" OR "Physical Therapists" OR "Physiotherapist" OR "Physiotherapists" OR "Physiotherapy" OR "Social Worker" OR "Social Worker" OR "Social Workers" OR "Social Work" OR "Social Work" OR "health center" OR "health centres" OR "health centre" OR "health centers" OR "health center" OR "health care centres" OR "health care centre" OR "health care centers" OR "health care center" OR "healthcare centres" OR "healthcare centre" OR "healthcare centers" OR "healthcare center" OR "health services" OR "health service" OR "health care services" OR "health care service" OR "healthcare services" OR "healthcare service" OR "social care")):ti,ab,kw

**OR**

(("referral" OR "referr*" OR "prescription" OR "prescrib*" OR "prescrip*") **NEAR/1** ("adult learning" OR "aqua therapy" OR "art" OR "art therapy" OR "arts" OR "befriending" OR "bibliotherap*" OR "bibliotherapy" OR "books" OR "community education group" OR "community education groups" OR "computerised cbt" OR "computerised cognitive behavioural therapy" OR "computerized cbt" OR "computerized cognitive behavioural therapy" OR "creativity" OR "cycling" OR "dance" OR "dance class" OR "dance classes" OR "dancing" OR "education" OR "exercise" OR "exercise class" OR "exercise classes" OR "exercising " OR "fishing" OR "fishing club" OR "fishing clubs" OR "gardening" OR "gardening club" OR "gardening clubs" OR "green gym" OR "group activities" OR "group activity" OR "guided walk" OR "guided walking" OR "guided walks" OR "gym" OR "gym-based activities" OR "gym-based activity" OR "gymnastic" OR "gymnastics" OR "health walk" OR "health walks" OR "knit club" OR "knit clubs" OR "knitting" OR "knitting club" OR "knitting clubs" OR "learning" OR "learning new skill" OR "learning new skills" OR "mutual aid" OR "natter club" OR "natter clubs" OR "physical activities" OR "physical activity" OR "self-help group" OR "self-help groups" OR "self-help reading" OR "signposting guidance" OR "signposting information" OR "supported education" OR "supported employment" OR "swimming" OR "team sport" OR "team sports" OR "time bank" OR "time banks" OR "volunteer" OR "volunteering" OR "volunteers" OR "walk" OR "walking" OR "walks")):ti

**)**

**Emcare** <http://ovidsp.ovid.com/ovidweb.cgi?T=JS&NEWS=n&CSC=Y&PAGE=main&D=emcr>

**(**("social prescribing".ti,ab OR "social prescri*".ti,ab OR "community prescribing".ti,ab OR "community prescri*".ti,ab OR "community referral".ti,ab OR "community referrals".ti,ab OR "community referr*".ti,ab OR "social referral".ti,ab OR "social referrals".ti,ab OR "social referr*".ti,ab OR "linking scheme".ti,ab OR "linking schemes".ti,ab OR "referral scheme".ti,ab OR "referral schemes".ti,ab) **OR** ("social" **ADJ1** "prescri*").ti,ab OR ("community" **ADJ1** "prescri*").ti,ab **OR** (("social intervention".ti,ab OR "social interventions".ti,ab OR "welfare intervention".ti,ab OR "welfare interventions".ti,ab) AND ("link worker".ti,ab OR "link workers".ti,ab OR "link work*".ti,ab OR "community navigators".ti,ab OR "community navigator".ti,ab OR "community liaisons".ti,ab OR "community liaison".ti,ab OR "well being coach".ti,ab OR "wellbeing coach".ti,ab OR "well being coaches".ti,ab OR "wellbeing coaches".ti,ab OR "welfare coordinators".ti,ab OR "welfare coordinator".ti,ab) AND (exp *"Primary Health Care"/ OR "Primary Health Care".ti,ab OR "Primary Healthcare".ti,ab OR "Primary Care".ti,ab OR exp *"General Practice"/ OR "Family Practice".ti,ab OR "General Practice".ti,ab OR exp *"General Practitioner"/ OR "General Practitioner".ti,ab OR "General Practitioners".ti,ab OR "GP".ti,ab OR "Family Physician".ti,ab OR "Family Physicians".ti,ab OR exp *"Physiotherapist"/ OR "Physical Therapist".ti,ab OR "Physical Therapists".ti,ab OR "Physiotherapist".ti,ab OR "Physiotherapists".ti,ab OR exp *"Physiotherapy"/ OR exp *"Social Worker"/ OR "Social Worker".ti,ab OR "Social Workers".ti,ab OR exp *"Social Work"/ OR "Social Work".ti,ab OR exp *"health center"/ OR "health centres".ti,ab OR "health centre".ti,ab OR "health centers".ti,ab OR "health center".ti,ab OR "health care centres".ti,ab OR "health care centre".ti,ab OR "health care centers".ti,ab OR "health care center".ti,ab OR "healthcare centres".ti,ab OR "healthcare centre".ti,ab OR "healthcare centers".ti,ab OR "healthcare center".ti,ab OR "health services".ti,ab OR "health service".ti,ab OR "health care services".ti,ab OR "health care service".ti,ab OR "healthcare services".ti,ab OR "healthcare service".ti,ab OR "social care".ti,ab)) **OR** "Arts on Prescription".ti,ab OR "Books on Prescription".ti,ab OR "Education on Prescription".ti,ab OR "exercise on prescription".ti,ab OR "exercise referral".ti,ab **OR** ((("referral" OR "referr*" OR "prescription" OR "prescrib*" OR "prescrip*") **ADJ1** ("adult learning" OR "aqua therapy" OR "art" OR "art therapy" OR "arts" OR "befriending" OR "bibliotherap*" OR "bibliotherapy" OR "books" OR "community education group" OR "community education groups" OR "computerised cbt" OR "computerised cognitive behavioural therapy" OR "computerized cbt" OR "computerized cognitive behavioural therapy" OR "creativity" OR "cycling" OR "dance" OR "dance class" OR "dance classes" OR "dancing" OR "education" OR "exercise" OR "exercise class" OR "exercise classes" OR "exercising " OR "fishing" OR "fishing club" OR "fishing clubs" OR "gardening" OR "gardening club" OR "gardening clubs" OR "green gym" OR "group activities" OR "group activity" OR "guided walk" OR "guided walking" OR "guided walks" OR "gym" OR "gym-based activities" OR "gym-based activity" OR "gymnastic" OR "gymnastics" OR "health walk" OR "health walks" OR "knit club" OR "knit clubs" OR "knitting" OR "knitting club" OR "knitting clubs" OR "learning" OR "learning new skill" OR "learning new skills" OR "mutual aid" OR "natter club" OR "natter clubs" OR "physical activities" OR "physical activity" OR "self-help group" OR "self-help groups" OR "self-help reading" OR "signposting guidance" OR "signposting information" OR "supported education" OR "supported employment" OR "swimming" OR "team sport" OR "team sports" OR "time bank" OR "time banks" OR "volunteer" OR "volunteering" OR "volunteers" OR "walk" OR "walking" OR "walks").ti,ab) AND (exp *"Primary Health Care"/ OR "Primary Health Care".ti,ab OR "Primary Healthcare".ti,ab OR "Primary Care".ti,ab OR exp *"General Practice"/ OR "Family Practice".ti,ab OR "General Practice".ti,ab OR exp *"General Practitioner"/ OR "General Practitioner".ti,ab OR "General Practitioners".ti,ab OR "GP".ti,ab OR "Family Physician".ti,ab OR "Family Physicians".ti,ab OR exp *"Physiotherapist"/ OR "Physical Therapist".ti,ab OR "Physical Therapists".ti,ab OR "Physiotherapist".ti,ab OR "Physiotherapists".ti,ab OR exp *"Physiotherapy"/ OR exp *"Social Worker"/ OR "Social Worker".ti,ab OR "Social Workers".ti,ab OR exp *"Social Work"/ OR "Social Work".ti,ab OR exp *"health center"/ OR "health centres".ti,ab OR "health centre".ti,ab OR "health centers".ti,ab OR "health center".ti,ab OR "health care centres".ti,ab OR "health care centre".ti,ab OR "health care centers".ti,ab OR "health care center".ti,ab OR "healthcare centres".ti,ab OR "healthcare centre".ti,ab OR "healthcare centers".ti,ab OR "healthcare center".ti,ab OR "health services".ti,ab OR "health service".ti,ab OR "health care services".ti,ab OR "health care service".ti,ab OR "healthcare services".ti,ab OR "healthcare service".ti,ab OR "social care".ti,ab)) **OR** (("referral" OR "referr*" OR "prescription" OR "prescrib*" OR "prescrip*") **ADJ1** ("adult learning" OR "aqua therapy" OR "art" OR "art therapy" OR "arts" OR "befriending" OR "bibliotherap*" OR "bibliotherapy" OR "books" OR "community education group" OR "community education groups" OR "computerised cbt" OR "computerised cognitive behavioural therapy" OR "computerized cbt" OR "computerized cognitive behavioural therapy" OR "creativity" OR "cycling" OR "dance" OR "dance class" OR "dance classes" OR "dancing" OR "education" OR "exercise" OR "exercise class" OR "exercise classes" OR "exercising " OR "fishing" OR "fishing club" OR "fishing clubs" OR "gardening" OR "gardening club" OR "gardening clubs" OR "green gym" OR "group activities" OR "group activity" OR "guided walk" OR "guided walking" OR "guided walks" OR "gym" OR "gym-based activities" OR "gym-based activity" OR "gymnastic" OR "gymnastics" OR "health walk" OR "health walks" OR "knit club" OR "knit clubs" OR "knitting" OR "knitting club" OR "knitting clubs" OR "learning" OR "learning new skill" OR "learning new skills" OR "mutual aid" OR "natter club" OR "natter clubs" OR "physical activities" OR "physical activity" OR "self-help group" OR "self-help groups" OR "self-help reading" OR "signposting guidance" OR "signposting information" OR "supported education" OR "supported employment" OR "swimming" OR "team sport" OR "team sports" OR "time bank" OR "time banks" OR "volunteer" OR "volunteering" OR "volunteers" OR "walk" OR "walking" OR "walks")).ti**)**

AND (exp "review"/ OR "systematic review"/ OR "review".mp OR "overview".mp OR "review*".mp OR "overview*".mp OR "appraisal".mp OR "synthesis".mp))

**Epistemonikos database**

<https://www.epistemonikos.org/en/>

("social prescribing" OR "social prescri*" OR "community prescribing" OR "community prescri*" OR "community referral" OR "community referrals" OR "community referr*" OR "social referral" OR "social referrals" OR "social referr*" OR "linking scheme" OR "linking schemes" OR "referral scheme" OR "referral schemes")

**PsycINFO**

<http://search.ebscohost.com/login.aspx?authtype=ip,uid&profile=lumc&defaultdb=psyh>

Limit to Academic Journals

4 queries

(TI("social prescribing" OR "social prescri*" OR "community prescribing" OR "community prescri*" OR "community referral" OR "community referrals" OR "community referr*" OR "social referral" OR "social referrals" OR "social referr*" OR "linking scheme" OR "linking schemes" OR "referral scheme" OR "referral schemes") OR SU("social prescribing" OR "social prescri*" OR "community prescribing" OR "community prescri*" OR "community referral" OR "community referrals" OR "community referr*" OR "social referral" OR "social referrals" OR "social referr*" OR "linking scheme" OR "linking schemes" OR "referral scheme" OR "referral schemes") OR MA("social prescribing" OR "social prescri*" OR "community prescribing" OR "community prescri*" OR "community referral" OR "community referrals" OR "community referr*" OR "social referral" OR "social referrals" OR "social referr*" OR "linking scheme" OR "linking schemes" OR "referral scheme" OR "referral schemes") OR AB("social prescribing" OR "social prescri*" OR "community prescribing" OR "community prescri*" OR "community referral" OR "community referrals" OR "community referr*" OR "social referral" OR "social referrals" OR "social referr*" OR "linking scheme" OR "linking schemes" OR "referral scheme" OR "referral schemes"))

**(**TI("social prescribing" OR "social prescri*" OR "community prescribing" OR "community prescri*" OR "community referral" OR "community referrals" OR "community referr*" OR "social referral" OR "social referrals" OR "social referr*" OR "linking scheme" OR "linking schemes" OR "referral scheme" OR "referral schemes") **OR** TI(("social" **N1** "prescri*") OR ("community" **N1** "prescri*")) **OR** TI(("social intervention" OR "social interventions" OR "welfare intervention" OR "welfare interventions") AND ("link worker" OR "link workers" OR "link work*" OR "community navigators" OR "community navigator" OR "community liaisons" OR "community liaison" OR "well being coach" OR "wellbeing coach" OR "well being coaches" OR "wellbeing coaches" OR "welfare coordinators" OR "welfare coordinator") AND ("Primary Health Care" OR "Primary Health Care" OR "Primary Healthcare" OR "Primary Care" OR "General Practice" OR "Family Practice" OR "General Practice" OR "General Practitioner" OR "General Practitioner" OR "General Practitioners" OR "GP" OR "Family Physician" OR "Family Physicians" OR "Physiotherapist" OR "Physical Therapist" OR "Physical Therapists" OR "Physiotherapist" OR "Physiotherapists" OR "Physiotherapy" OR "Social Worker" OR "Social Worker" OR "Social Workers" OR "Social Work" OR "Social Work" OR "health center" OR "health centres" OR "health centre" OR "health centers" OR "health center" OR "health care centres" OR "health care centre" OR "health care centers" OR "health care center" OR "healthcare centres" OR "healthcare centre" OR "healthcare centers" OR "healthcare center" OR "health services" OR "health service" OR "health care services" OR "health care service" OR "healthcare services" OR "healthcare service" OR "social care")) **OR** TI("Arts on Prescription" OR "Books on Prescription" OR "Education on Prescription" OR "exercise on prescription" OR "exercise referral") **OR** TI((("referral" OR "referral*" OR "prescription" OR "prescrib*" OR "prescrip*") **N1** ("adult learning" OR "aqua therapy" OR "art" OR "art therapy" OR "arts" OR "befriending" OR "bibliotherap*" OR "bibliotherapy" OR "books" OR "community education group" OR "community education groups" OR "computerised cbt" OR "computerised cognitive behavioural therapy" OR "computerized cbt" OR "computerized cognitive behavioural therapy" OR "creativity" OR "cycling" OR "dance" OR "dance class" OR "dance classes" OR "dancing" OR "education" OR "exercise" OR "exercise class" OR "exercise classes" OR "exercising " OR "fishing" OR "fishing club" OR "fishing clubs" OR "gardening" OR "gardening club" OR "gardening clubs" OR "green gym" OR "group activities" OR "group activity" OR "guided walk" OR "guided walking" OR "guided walks" OR "gym" OR "gym-based activities" OR "gym-based activity" OR "gymnastic" OR "gymnastics" OR "health walk" OR "health walks" OR "knit club" OR "knit clubs" OR "knitting" OR "knitting club" OR "knitting clubs" OR "learning" OR "learning new skill" OR "learning new skills" OR "mutual aid" OR "natter club" OR "natter clubs" OR "physical activities" OR "physical activity" OR "self-help group" OR "self-help groups" OR "self-help reading" OR "signposting guidance" OR "signposting information" OR "supported education" OR "supported employment" OR "swimming" OR "team sport" OR "team sports" OR "time bank" OR "time banks" OR "volunteer" OR "volunteering" OR "volunteers" OR "walk" OR "walking" OR "walks")) AND ("Primary Health Care" OR "Primary Health Care" OR "Primary Healthcare" OR "Primary Care" OR "General Practice" OR "Family Practice" OR "General Practice" OR "General Practitioner" OR "General Practitioner" OR "General Practitioners" OR "GP" OR "Family Physician" OR "Family Physicians" OR "Physiotherapist" OR "Physical Therapist" OR "Physical Therapists" OR "Physiotherapist" OR "Physiotherapists" OR "Physiotherapy" OR "Social Worker" OR "Social Worker" OR "Social Workers" OR "Social Work" OR "Social Work" OR "health center" OR "health centres" OR "health centre" OR "health centers" OR "health center" OR "health care centres" OR "health care centre" OR "health care centers" OR "health care center" OR "healthcare centres" OR "healthcare centre" OR "healthcare centers" OR "healthcare center" OR "health services" OR "health service" OR "health care services" OR "health care service" OR "healthcare services" OR "healthcare service" OR "social care")) **OR** TI(("referral" OR "referral*" OR "prescription" OR "prescrib*" OR "prescrip*") **N1** ("adult learning" OR "aqua therapy" OR "art" OR "art therapy" OR "arts" OR "befriending" OR "bibliotherap*" OR "bibliotherapy" OR "books" OR "community education group" OR "community education groups" OR "computerised cbt" OR "computerised cognitive behavioural therapy" OR "computerized cbt" OR "computerized cognitive behavioural therapy" OR "creativity" OR "cycling" OR "dance" OR "dance class" OR "dance classes" OR "dancing" OR "education" OR "exercise" OR "exercise class" OR "exercise classes" OR "exercising " OR "fishing" OR "fishing club" OR "fishing clubs" OR "gardening" OR "gardening club" OR "gardening clubs" OR "green gym" OR "group activities" OR "group activity" OR "guided walk" OR "guided walking" OR "guided walks" OR "gym" OR "gym-based activities" OR "gym-based activity" OR "gymnastic" OR "gymnastics" OR "health walk" OR "health walks" OR "knit club" OR "knit clubs" OR "knitting" OR "knitting club" OR "knitting clubs" OR "learning" OR "learning new skill" OR "learning new skills" OR "mutual aid" OR "natter club" OR "natter clubs" OR "physical activities" OR "physical activity" OR "self-help group" OR "self-help groups" OR "self-help reading" OR "signposting guidance" OR "signposting information" OR "supported education" OR "supported employment" OR "swimming" OR "team sport" OR "team sports" OR "time bank" OR "time banks" OR "volunteer" OR "volunteering" OR "volunteers" OR "walk" OR "walking" OR "walks"))**)**

**(**SU("social prescribing" OR "social prescri*" OR "community prescribing" OR "community prescri*" OR "community referral" OR "community referrals" OR "community referr*" OR "social referral" OR "social referrals" OR "social referr*" OR "linking scheme" OR "linking schemes" OR "referral scheme" OR "referral schemes") **OR** SU(("social" **N1** "prescri*") OR ("community" **N1** "prescri*")) **OR** SU(("social intervention" OR "social interventions" OR "welfare intervention" OR "welfare interventions") AND ("link worker" OR "link workers" OR "link work*" OR "community navigators" OR "community navigator" OR "community liaisons" OR "community liaison" OR "well being coach" OR "wellbeing coach" OR "well being coaches" OR "wellbeing coaches" OR "welfare coordinators" OR "welfare coordinator") AND ("Primary Health Care" OR "Primary Health Care" OR "Primary Healthcare" OR "Primary Care" OR "General Practice" OR "Family Practice" OR "General Practice" OR "General Practitioner" OR "General Practitioner" OR "General Practitioners" OR "GP" OR "Family Physician" OR "Family Physicians" OR "Physiotherapist" OR "Physical Therapist" OR "Physical Therapists" OR "Physiotherapist" OR "Physiotherapists" OR "Physiotherapy" OR "Social Worker" OR "Social Worker" OR "Social Workers" OR "Social Work" OR "Social Work" OR "health center" OR "health centres" OR "health centre" OR "health centers" OR "health center" OR "health care centres" OR "health care centre" OR "health care centers" OR "health care center" OR "healthcare centres" OR "healthcare centre" OR "healthcare centers" OR "healthcare center" OR "health services" OR "health service" OR "health care services" OR "health care service" OR "healthcare services" OR "healthcare service" OR "social care")) **OR** SU("Arts on Prescription" OR "Books on Prescription" OR "Education on Prescription" OR "exercise on prescription" OR "exercise referral") **OR** SU((("referral" OR "referral*" OR "prescription" OR "prescrib*" OR "prescrip*") **N1** ("adult learning" OR "aqua therapy" OR "art" OR "art therapy" OR "arts" OR "befriending" OR "bibliotherap*" OR "bibliotherapy" OR "books" OR "community education group" OR "community education groups" OR "computerised cbt" OR "computerised cognitive behavioural therapy" OR "computerized cbt" OR "computerized cognitive behavioural therapy" OR "creativity" OR "cycling" OR "dance" OR "dance class" OR "dance classes" OR "dancing" OR "education" OR "exercise" OR "exercise class" OR "exercise classes" OR "exercising " OR "fishing" OR "fishing club" OR "fishing clubs" OR "gardening" OR "gardening club" OR "gardening clubs" OR "green gym" OR "group activities" OR "group activity" OR "guided walk" OR "guided walking" OR "guided walks" OR "gym" OR "gym-based activities" OR "gym-based activity" OR "gymnastic" OR "gymnastics" OR "health walk" OR "health walks" OR "knit club" OR "knit clubs" OR "knitting" OR "knitting club" OR "knitting clubs" OR "learning" OR "learning new skill" OR "learning new skills" OR "mutual aid" OR "natter club" OR "natter clubs" OR "physical activities" OR "physical activity" OR "self-help group" OR "self-help groups" OR "self-help reading" OR "signposting guidance" OR "signposting information" OR "supported education" OR "supported employment" OR "swimming" OR "team sport" OR "team sports" OR "time bank" OR "time banks" OR "volunteer" OR "volunteering" OR "volunteers" OR "walk" OR "walking" OR "walks")) AND ("Primary Health Care" OR "Primary Health Care" OR "Primary Healthcare" OR "Primary Care" OR "General Practice" OR "Family Practice" OR "General Practice" OR "General Practitioner" OR "General Practitioner" OR "General Practitioners" OR "GP" OR "Family Physician" OR "Family Physicians" OR "Physiotherapist" OR "Physical Therapist" OR "Physical Therapists" OR "Physiotherapist" OR "Physiotherapists" OR "Physiotherapy" OR "Social Worker" OR "Social Worker" OR "Social Workers" OR "Social Work" OR "Social Work" OR "health center" OR "health centres" OR "health centre" OR "health centers" OR "health center" OR "health care centres" OR "health care centre" OR "health care centers" OR "health care center" OR "healthcare centres" OR "healthcare centre" OR "healthcare centers" OR "healthcare center" OR "health services" OR "health service" OR "health care services" OR "health care service" OR "healthcare services" OR "healthcare service" OR "social care"))**)**

**(**AB("social prescribing" OR "social prescri*" OR "community prescribing" OR "community prescri*" OR "community referral" OR "community referrals" OR "community referr*" OR "social referral" OR "social referrals" OR "social referr*" OR "linking scheme" OR "linking schemes" OR "referral scheme" OR "referral schemes") **OR** AB(("social" **N1** "prescri*") OR ("community" **N1** "prescri*")) **OR** AB(("social intervention" OR "social interventions" OR "welfare intervention" OR "welfare interventions") AND ("link worker" OR "link workers" OR "link work*" OR "community navigators" OR "community navigator" OR "community liaisons" OR "community liaison" OR "well being coach" OR "wellbeing coach" OR "well being coaches" OR "wellbeing coaches" OR "welfare coordinators" OR "welfare coordinator") AND ("Primary Health Care" OR "Primary Health Care" OR "Primary Healthcare" OR "Primary Care" OR "General Practice" OR "Family Practice" OR "General Practice" OR "General Practitioner" OR "General Practitioner" OR "General Practitioners" OR "GP" OR "Family Physician" OR "Family Physicians" OR "Physiotherapist" OR "Physical Therapist" OR "Physical Therapists" OR "Physiotherapist" OR "Physiotherapists" OR "Physiotherapy" OR "Social Worker" OR "Social Worker" OR "Social Workers" OR "Social Work" OR "Social Work" OR "health center" OR "health centres" OR "health centre" OR "health centers" OR "health center" OR "health care centres" OR "health care centre" OR "health care centers" OR "health care center" OR "healthcare centres" OR "healthcare centre" OR "healthcare centers" OR "healthcare center" OR "health services" OR "health service" OR "health care services" OR "health care service" OR "healthcare services" OR "healthcare service" OR "social care")) **OR** AB("Arts on Prescription" OR "Books on Prescription" OR "Education on Prescription" OR "exercise on prescription" OR "exercise referral") **OR** AB(AB(("referral" OR "referral*" OR "prescription" OR "prescrib*" OR "prescrip*") **N1** ("adult learning" OR "aqua therapy" OR "art" OR "art therapy" OR "arts" OR "befriending" OR "bibliotherap*" OR "bibliotherapy" OR "books" OR "community education group" OR "community education groups" OR "computerised cbt" OR "computerised cognitive behavioural therapy" OR "computerized cbt" OR "computerized cognitive behavioural therapy" OR "creativity" OR "cycling" OR "dance" OR "dance class" OR "dance classes" OR "dancing" OR "education" OR "exercise" OR "exercise class" OR "exercise classes" OR "exercising " OR "fishing" OR "fishing club" OR "fishing clubs" OR "gardening" OR "gardening club" OR "gardening clubs" OR "green gym" OR "group activities" OR "group activity" OR "guided walk" OR "guided walking" OR "guided walks" OR "gym" OR "gym-based activities" OR "gym-based activity" OR "gymnastic" OR "gymnastics" OR "health walk" OR "health walks" OR "knit club" OR "knit clubs" OR "knitting" OR "knitting club" OR "knitting clubs" OR "learning" OR "learning new skill" OR "learning new skills" OR "mutual aid" OR "natter club" OR "natter clubs" OR "physical activities" OR "physical activity" OR "self-help group" OR "self-help groups" OR "self-help reading" OR "signposting guidance" OR "signposting information" OR "supported education" OR "supported employment" OR "swimming" OR "team sport" OR "team sports" OR "time bank" OR "time banks" OR "volunteer" OR "volunteering" OR "volunteers" OR "walk" OR "walking" OR "walks")) AND TI("Primary Health Care" OR "Primary Health Care" OR "Primary Healthcare" OR "Primary Care" OR "General Practice" OR "Family Practice" OR "General Practice" OR "General Practitioner" OR "General Practitioner" OR "General Practitioners" OR "GP" OR "Family Physician" OR "Family Physicians" OR "Physiotherapist" OR "Physical Therapist" OR "Physical Therapists" OR "Physiotherapist" OR "Physiotherapists" OR "Physiotherapy" OR "Social Worker" OR "Social Worker" OR "Social Workers" OR "Social Work" OR "Social Work" OR "health center" OR "health centres" OR "health centre" OR "health centers" OR "health center" OR "health care centres" OR "health care centre" OR "health care centers" OR "health care center" OR "healthcare centres" OR "healthcare centre" OR "healthcare centers" OR "healthcare center" OR "health services" OR "health service" OR "health care services" OR "health care service" OR "healthcare services" OR "healthcare service" OR "social care"))**)**

AND

(TI("review" OR "systematic review" OR "review" OR "overview" OR "review*" OR "overview*" OR "appraisal" OR "synthesis") OR SU("review" OR "systematic review" OR "review" OR "overview" OR "review*" OR "overview*" OR "appraisal" OR "synthesis") OR MA("review" OR "systematic review" OR "review" OR "overview" OR "review*" OR "overview*" OR "appraisal" OR "synthesis") OR AB("review" OR "systematic review" OR "review" OR "overview" OR "review*" OR "overview*" OR "appraisal" OR "synthesis") OR MR("review" OR "systematic review" OR "review" OR "overview" OR "review*" OR "overview*" OR "appraisal" OR "synthesis"))

**Academic Search Premier**

<http://search.ebscohost.com/login.aspx?authtype=ip,uid&profile=lumc&defaultdb=aph>

5 queries

(TI("social prescribing" OR "social prescri*" OR "community prescribing" OR "community prescri*" OR "community referral" OR "community referrals" OR "community referr*" OR "social referral" OR "social referrals" OR "social referr*" OR "linking scheme" OR "linking schemes" OR "referral scheme" OR "referral schemes") OR SU("social prescribing" OR "social prescri*" OR "community prescribing" OR "community prescri*" OR "community referral" OR "community referrals" OR "community referr*" OR "social referral" OR "social referrals" OR "social referr*" OR "linking scheme" OR "linking schemes" OR "referral scheme" OR "referral schemes") OR KW("social prescribing" OR "social prescri*" OR "community prescribing" OR "community prescri*" OR "community referral" OR "community referrals" OR "community referr*" OR "social referral" OR "social referrals" OR "social referr*" OR "linking scheme" OR "linking schemes" OR "referral scheme" OR "referral schemes"))

**(**TI("social prescribing" OR "social prescri*" OR "community prescribing" OR "community prescri*" OR "community referral" OR "community referrals" OR "community referr*" OR "social referral" OR "social referrals" OR "social referr*" OR "linking scheme" OR "linking schemes" OR "referral scheme" OR "referral schemes") **OR** TI(("social" **N1** "prescri*") OR ("community" **N1** "prescri*")) **OR** TI(("social intervention" OR "social interventions" OR "welfare intervention" OR "welfare interventions") AND ("link worker" OR "link workers" OR "link work*" OR "community navigators" OR "community navigator" OR "community liaisons" OR "community liaison" OR "well being coach" OR "wellbeing coach" OR "well being coaches" OR "wellbeing coaches" OR "welfare coordinators" OR "welfare coordinator") AND ("Primary Health Care" OR "Primary Health Care" OR "Primary Healthcare" OR "Primary Care" OR "General Practice" OR "Family Practice" OR "General Practice" OR "General Practitioner" OR "General Practitioner" OR "General Practitioners" OR "GP" OR "Family Physician" OR "Family Physicians" OR "Physiotherapist" OR "Physical Therapist" OR "Physical Therapists" OR "Physiotherapist" OR "Physiotherapists" OR "Physiotherapy" OR "Social Worker" OR "Social Worker" OR "Social Workers" OR "Social Work" OR "Social Work" OR "health center" OR "health centres" OR "health centre" OR "health centers" OR "health center" OR "health care centres" OR "health care centre" OR "health care centers" OR "health care center" OR "healthcare centres" OR "healthcare centre" OR "healthcare centers" OR "healthcare center" OR "health services" OR "health service" OR "health care services" OR "health care service" OR "healthcare services" OR "healthcare service" OR "social care")) **OR** TI("Arts on Prescription" OR "Books on Prescription" OR "Education on Prescription" OR "exercise on prescription" OR "exercise referral") **OR** TI((("referral" OR "referral*" OR "prescription" OR "prescrib*" OR "prescrip*") **N1** ("adult learning" OR "aqua therapy" OR "art" OR "art therapy" OR "arts" OR "befriending" OR "bibliotherap*" OR "bibliotherapy" OR "books" OR "community education group" OR "community education groups" OR "computerised cbt" OR "computerised cognitive behavioural therapy" OR "computerized cbt" OR "computerized cognitive behavioural therapy" OR "creativity" OR "cycling" OR "dance" OR "dance class" OR "dance classes" OR "dancing" OR "education" OR "exercise" OR "exercise class" OR "exercise classes" OR "exercising " OR "fishing" OR "fishing club" OR "fishing clubs" OR "gardening" OR "gardening club" OR "gardening clubs" OR "green gym" OR "group activities" OR "group activity" OR "guided walk" OR "guided walking" OR "guided walks" OR "gym" OR "gym-based activities" OR "gym-based activity" OR "gymnastic" OR "gymnastics" OR "health walk" OR "health walks" OR "knit club" OR "knit clubs" OR "knitting" OR "knitting club" OR "knitting clubs" OR "learning" OR "learning new skill" OR "learning new skills" OR "mutual aid" OR "natter club" OR "natter clubs" OR "physical activities" OR "physical activity" OR "self-help group" OR "self-help groups" OR "self-help reading" OR "signposting guidance" OR "signposting information" OR "supported education" OR "supported employment" OR "swimming" OR "team sport" OR "team sports" OR "time bank" OR "time banks" OR "volunteer" OR "volunteering" OR "volunteers" OR "walk" OR "walking" OR "walks")) AND ("Primary Health Care" OR "Primary Health Care" OR "Primary Healthcare" OR "Primary Care" OR "General Practice" OR "Family Practice" OR "General Practice" OR "General Practitioner" OR "General Practitioner" OR "General Practitioners" OR "GP" OR "Family Physician" OR "Family Physicians" OR "Physiotherapist" OR "Physical Therapist" OR "Physical Therapists" OR "Physiotherapist" OR "Physiotherapists" OR "Physiotherapy" OR "Social Worker" OR "Social Worker" OR "Social Workers" OR "Social Work" OR "Social Work" OR "health center" OR "health centres" OR "health centre" OR "health centers" OR "health center" OR "health care centres" OR "health care centre" OR "health care centers" OR "health care center" OR "healthcare centres" OR "healthcare centre" OR "healthcare centers" OR "healthcare center" OR "health services" OR "health service" OR "health care services" OR "health care service" OR "healthcare services" OR "healthcare service" OR "social care")) **OR** TI(("referral" OR "referral*" OR "prescription" OR "prescrib*" OR "prescrip*") **N1** ("adult learning" OR "aqua therapy" OR "art" OR "art therapy" OR "arts" OR "befriending" OR "bibliotherap*" OR "bibliotherapy" OR "books" OR "community education group" OR "community education groups" OR "computerised cbt" OR "computerised cognitive behavioural therapy" OR "computerized cbt" OR "computerized cognitive behavioural therapy" OR "creativity" OR "cycling" OR "dance" OR "dance class" OR "dance classes" OR "dancing" OR "education" OR "exercise" OR "exercise class" OR "exercise classes" OR "exercising " OR "fishing" OR "fishing club" OR "fishing clubs" OR "gardening" OR "gardening club" OR "gardening clubs" OR "green gym" OR "group activities" OR "group activity" OR "guided walk" OR "guided walking" OR "guided walks" OR "gym" OR "gym-based activities" OR "gym-based activity" OR "gymnastic" OR "gymnastics" OR "health walk" OR "health walks" OR "knit club" OR "knit clubs" OR "knitting" OR "knitting club" OR "knitting clubs" OR "learning" OR "learning new skill" OR "learning new skills" OR "mutual aid" OR "natter club" OR "natter clubs" OR "physical activities" OR "physical activity" OR "self-help group" OR "self-help groups" OR "self-help reading" OR "signposting guidance" OR "signposting information" OR "supported education" OR "supported employment" OR "swimming" OR "team sport" OR "team sports" OR "time bank" OR "time banks" OR "volunteer" OR "volunteering" OR "volunteers" OR "walk" OR "walking" OR "walks"))**)**

**(**SU("social prescribing" OR "social prescri*" OR "community prescribing" OR "community prescri*" OR "community referral" OR "community referrals" OR "community referr*" OR "social referral" OR "social referrals" OR "social referr*" OR "linking scheme" OR "linking schemes" OR "referral scheme" OR "referral schemes") **OR** SU(("social intervention" OR "social interventions" OR "welfare intervention" OR "welfare interventions") AND ("link worker" OR "link workers" OR "link work*" OR "community navigators" OR "community navigator" OR "community liaisons" OR "community liaison" OR "well being coach" OR "wellbeing coach" OR "well being coaches" OR "wellbeing coaches" OR "welfare coordinators" OR "welfare coordinator") AND ("Primary Health Care" OR "Primary Health Care" OR "Primary Healthcare" OR "Primary Care" OR "General Practice" OR "Family Practice" OR "General Practice" OR "General Practitioner" OR "General Practitioner" OR "General Practitioners" OR "GP" OR "Family Physician" OR "Family Physicians" OR "Physiotherapist" OR "Physical Therapist" OR "Physical Therapists" OR "Physiotherapist" OR "Physiotherapists" OR "Physiotherapy" OR "Social Worker" OR "Social Worker" OR "Social Workers" OR "Social Work" OR "Social Work" OR "health center" OR "health centres" OR "health centre" OR "health centers" OR "health center" OR "health care centres" OR "health care centre" OR "health care centers" OR "health care center" OR "healthcare centres" OR "healthcare centre" OR "healthcare centers" OR "healthcare center" OR "health services" OR "health service" OR "health care services" OR "health care service" OR "healthcare services" OR "healthcare service" OR "social care")) **OR** SU("Arts on Prescription" OR "Books on Prescription" OR "Education on Prescription" OR "exercise on prescription" OR "exercise referral") **OR** SU((("referral" OR "referral*" OR "prescription" OR "prescrib*" OR "prescrip*") **N1** ("adult learning" OR "aqua therapy" OR "art" OR "art therapy" OR "arts" OR "befriending" OR "bibliotherap*" OR "bibliotherapy" OR "books" OR "community education group" OR "community education groups" OR "computerised cbt" OR "computerised cognitive behavioural therapy" OR "computerized cbt" OR "computerized cognitive behavioural therapy" OR "creativity" OR "cycling" OR "dance" OR "dance class" OR "dance classes" OR "dancing" OR "education" OR "exercise" OR "exercise class" OR "exercise classes" OR "exercising " OR "fishing" OR "fishing club" OR "fishing clubs" OR "gardening" OR "gardening club" OR "gardening clubs" OR "green gym" OR "group activities" OR "group activity" OR "guided walk" OR "guided walking" OR "guided walks" OR "gym" OR "gym-based activities" OR "gym-based activity" OR "gymnastic" OR "gymnastics" OR "health walk" OR "health walks" OR "knit club" OR "knit clubs" OR "knitting" OR "knitting club" OR "knitting clubs" OR "learning" OR "learning new skill" OR "learning new skills" OR "mutual aid" OR "natter club" OR "natter clubs" OR "physical activities" OR "physical activity" OR "self-help group" OR "self-help groups" OR "self-help reading" OR "signposting guidance" OR "signposting information" OR "supported education" OR "supported employment" OR "swimming" OR "team sport" OR "team sports" OR "time bank" OR "time banks" OR "volunteer" OR "volunteering" OR "volunteers" OR "walk" OR "walking" OR "walks")) AND ("Primary Health Care" OR "Primary Health Care" OR "Primary Healthcare" OR "Primary Care" OR "General Practice" OR "Family Practice" OR "General Practice" OR "General Practitioner" OR "General Practitioner" OR "General Practitioners" OR "GP" OR "Family Physician" OR "Family Physicians" OR "Physiotherapist" OR "Physical Therapist" OR "Physical Therapists" OR "Physiotherapist" OR "Physiotherapists" OR "Physiotherapy" OR "Social Worker" OR "Social Worker" OR "Social Workers" OR "Social Work" OR "Social Work" OR "health center" OR "health centres" OR "health centre" OR "health centers" OR "health center" OR "health care centres" OR "health care centre" OR "health care centers" OR "health care center" OR "healthcare centres" OR "healthcare centre" OR "healthcare centers" OR "healthcare center" OR "health services" OR "health service" OR "health care services" OR "health care service" OR "healthcare services" OR "healthcare service" OR "social care"))**)**

**(**KW("social prescribing" OR "social prescri*" OR "community prescribing" OR "community prescri*" OR "community referral" OR "community referrals" OR "community referr*" OR "social referral" OR "social referrals" OR "social referr*" OR "linking scheme" OR "linking schemes" OR "referral scheme" OR "referral schemes") **OR** KW(("social intervention" OR "social interventions" OR "welfare intervention" OR "welfare interventions") AND ("link worker" OR "link workers" OR "link work*" OR "community navigators" OR "community navigator" OR "community liaisons" OR "community liaison" OR "well being coach" OR "wellbeing coach" OR "well being coaches" OR "wellbeing coaches" OR "welfare coordinators" OR "welfare coordinator") AND ("Primary Health Care" OR "Primary Health Care" OR "Primary Healthcare" OR "Primary Care" OR "General Practice" OR "Family Practice" OR "General Practice" OR "General Practitioner" OR "General Practitioner" OR "General Practitioners" OR "GP" OR "Family Physician" OR "Family Physicians" OR "Physiotherapist" OR "Physical Therapist" OR "Physical Therapists" OR "Physiotherapist" OR "Physiotherapists" OR "Physiotherapy" OR "Social Worker" OR "Social Worker" OR "Social Workers" OR "Social Work" OR "Social Work" OR "health center" OR "health centres" OR "health centre" OR "health centers" OR "health center" OR "health care centres" OR "health care centre" OR "health care centers" OR "health care center" OR "healthcare centres" OR "healthcare centre" OR "healthcare centers" OR "healthcare center" OR "health services" OR "health service" OR "health care services" OR "health care service" OR "healthcare services" OR "healthcare service" OR "social care")) **OR** KW("Arts on Prescription" OR "Books on Prescription" OR "Education on Prescription" OR "exercise on prescription" OR "exercise referral") **OR** KW((("referral" OR "referral*" OR "prescription" OR "prescrib*" OR "prescrip*") **N1** ("adult learning" OR "aqua therapy" OR "art" OR "art therapy" OR "arts" OR "befriending" OR "bibliotherap*" OR "bibliotherapy" OR "books" OR "community education group" OR "community education groups" OR "computerised cbt" OR "computerised cognitive behavioural therapy" OR "computerized cbt" OR "computerized cognitive behavioural therapy" OR "creativity" OR "cycling" OR "dance" OR "dance class" OR "dance classes" OR "dancing" OR "education" OR "exercise" OR "exercise class" OR "exercise classes" OR "exercising " OR "fishing" OR "fishing club" OR "fishing clubs" OR "gardening" OR "gardening club" OR "gardening clubs" OR "green gym" OR "group activities" OR "group activity" OR "guided walk" OR "guided walking" OR "guided walks" OR "gym" OR "gym-based activities" OR "gym-based activity" OR "gymnastic" OR "gymnastics" OR "health walk" OR "health walks" OR "knit club" OR "knit clubs" OR "knitting" OR "knitting club" OR "knitting clubs" OR "learning" OR "learning new skill" OR "learning new skills" OR "mutual aid" OR "natter club" OR "natter clubs" OR "physical activities" OR "physical activity" OR "self-help group" OR "self-help groups" OR "self-help reading" OR "signposting guidance" OR "signposting information" OR "supported education" OR "supported employment" OR "swimming" OR "team sport" OR "team sports" OR "time bank" OR "time banks" OR "volunteer" OR "volunteering" OR "volunteers" OR "walk" OR "walking" OR "walks")) AND ("Primary Health Care" OR "Primary Health Care" OR "Primary Healthcare" OR "Primary Care" OR "General Practice" OR "Family Practice" OR "General Practice" OR "General Practitioner" OR "General Practitioner" OR "General Practitioners" OR "GP" OR "Family Physician" OR "Family Physicians" OR "Physiotherapist" OR "Physical Therapist" OR "Physical Therapists" OR "Physiotherapist" OR "Physiotherapists" OR "Physiotherapy" OR "Social Worker" OR "Social Worker" OR "Social Workers" OR "Social Work" OR "Social Work" OR "health center" OR "health centres" OR "health centre" OR "health centers" OR "health center" OR "health care centres" OR "health care centre" OR "health care centers" OR "health care center" OR "healthcare centres" OR "healthcare centre" OR "healthcare centers" OR "healthcare center" OR "health services" OR "health service" OR "health care services" OR "health care service" OR "healthcare services" OR "healthcare service" OR "social care"))**)**

TI((("referral" OR "referral*" OR "prescription" OR "prescrib*" OR "prescrip*") **N1** ("adult learning" OR "aqua therapy" OR "art" OR "art therapy" OR "arts" OR "befriending" OR "bibliotherap*" OR "bibliotherapy" OR "books" OR "community education group" OR "community education groups" OR "computerised cbt" OR "computerised cognitive behavioural therapy" OR "computerized cbt" OR "computerized cognitive behavioural therapy" OR "creativity" OR "cycling" OR "dance" OR "dance class" OR "dance classes" OR "dancing" OR "education" OR "exercise" OR "exercise class" OR "exercise classes" OR "exercising " OR "fishing" OR "fishing club" OR "fishing clubs" OR "gardening" OR "gardening club" OR "gardening clubs" OR "green gym" OR "group activities" OR "group activity" OR "guided walk" OR "guided walking" OR "guided walks" OR "gym" OR "gym-based activities" OR "gym-based activity" OR "gymnastic" OR "gymnastics" OR "health walk" OR "health walks" OR "knit club" OR "knit clubs" OR "knitting" OR "knitting club" OR "knitting clubs" OR "learning" OR "learning new skill" OR "learning new skills" OR "mutual aid" OR "natter club" OR "natter clubs" OR "physical activities" OR "physical activity" OR "self-help group" OR "self-help groups" OR "self-help reading" OR "signposting guidance" OR "signposting information" OR "supported education" OR "supported employment" OR "swimming" OR "team sport" OR "team sports" OR "time bank" OR "time banks" OR "volunteer" OR "volunteering" OR "volunteers" OR "walk" OR "walking" OR "walks")) AND ("Primary Health Care" OR "Primary Health Care" OR "Primary Healthcare" OR "Primary Care" OR "General Practice" OR "Family Practice" OR "General Practice" OR "General Practitioner" OR "General Practitioner" OR "General Practitioners" OR "GP" OR "Family Physician" OR "Family Physicians" OR "Physiotherapist" OR "Physical Therapist" OR "Physical Therapists" OR "Physiotherapist" OR "Physiotherapists" OR "Physiotherapy" OR "Social Worker" OR "Social Worker" OR "Social Workers" OR "Social Work" OR "Social Work" OR "health center" OR "health centres" OR "health centre" OR "health centers" OR "health center" OR "health care centres" OR "health care centre" OR "health care centers" OR "health care center" OR "healthcare centres" OR "healthcare centre" OR "healthcare centers" OR "healthcare center" OR "health services" OR "health service" OR "health care services" OR "health care service" OR "healthcare services" OR "healthcare service" OR "social care"))

AND

(TI("review" OR "systematic review" OR "review" OR "overview" OR "review*" OR "overview*" OR "appraisal" OR "synthesis") OR SU("review" OR "systematic review" OR "review" OR "overview" OR "review*" OR "overview*" OR "appraisal" OR "synthesis") OR KW("review" OR "systematic review" OR "review" OR "overview" OR "review*" OR "overview*" OR "appraisal" OR "synthesis") OR AB("review" OR "systematic review" OR "review" OR "overview" OR "review*" OR "overview*" OR "appraisal" OR "synthesis"))

**Social Services Abstracts**

<https://search.proquest.com/socialservices?accountid=12045>

<http://databases.library.leiden.edu/?bibid=990027333030302711&redirect=true>

Limit to scholarly journals

3 queries

NOFT("social prescribing" OR "social prescri*" OR "community prescribing" OR "community prescri*" OR "community referral" OR "community referrals" OR "community referr*" OR "social referral" OR "social referrals" OR "social referr*" OR "linking scheme" OR "linking schemes" OR "referral scheme" OR "referral schemes")

**(**TI("social prescribing" OR "social prescri*" OR "community prescribing" OR "community prescri*" OR "community referral" OR "community referrals" OR "community referr*" OR "social referral" OR "social referrals" OR "social referr*" OR "linking scheme" OR "linking schemes" OR "referral scheme" OR "referral schemes") **OR** TI(("social" **NEAR/1** "prescri*") OR ("community" **NEAR/1** "prescri*")) **OR** TI(("social intervention" OR "social interventions" OR "welfare intervention" OR "welfare interventions") AND ("link worker" OR "link workers" OR "link work*" OR "community navigators" OR "community navigator" OR "community liaisons" OR "community liaison" OR "well being coach" OR "wellbeing coach" OR "well being coaches" OR "wellbeing coaches" OR "welfare coordinators" OR "welfare coordinator") AND ("Primary Health Care" OR "Primary Health Care" OR "Primary Healthcare" OR "Primary Care" OR "General Practice" OR "Family Practice" OR "General Practice" OR "General Practitioner" OR "General Practitioner" OR "General Practitioners" OR "GP" OR "Family Physician" OR "Family Physicians" OR "Physiotherapist" OR "Physical Therapist" OR "Physical Therapists" OR "Physiotherapist" OR "Physiotherapists" OR "Physiotherapy" OR "Social Worker" OR "Social Worker" OR "Social Workers" OR "Social Work" OR "Social Work" OR "health center" OR "health centres" OR "health centre" OR "health centers" OR "health center" OR "health care centres" OR "health care centre" OR "health care centers" OR "health care center" OR "healthcare centres" OR "healthcare centre" OR "healthcare centers" OR "healthcare center" OR "health services" OR "health service" OR "health care services" OR "health care service" OR "healthcare services" OR "healthcare service" OR "social care")) **OR** TI("Arts on Prescription" OR "Books on Prescription" OR "Education on Prescription" OR "exercise on prescription" OR "exercise referral") **OR** TI((("referral" OR "referr*" OR "prescription" OR "prescrib*" OR "prescrip*") **NEAR/1** ("adult learning" OR "aqua therapy" OR "art" OR "art therapy" OR "arts" OR "befriending" OR "bibliotherap*" OR "bibliotherapy" OR "books" OR "community education group" OR "community education groups" OR "computerised cbt" OR "computerised cognitive behavioural therapy" OR "computerized cbt" OR "computerized cognitive behavioural therapy" OR "creativity" OR "cycling" OR "dance" OR "dance class" OR "dance classes" OR "dancing" OR "education" OR "exercise" OR "exercise class" OR "exercise classes" OR "exercising " OR "fishing" OR "fishing club" OR "fishing clubs" OR "gardening" OR "gardening club" OR "gardening clubs" OR "green gym" OR "group activities" OR "group activity" OR "guided walk" OR "guided walking" OR "guided walks" OR "gym" OR "gym-based activities" OR "gym-based activity" OR "gymnastic" OR "gymnastics" OR "health walk" OR "health walks" OR "knit club" OR "knit clubs" OR "knitting" OR "knitting club" OR "knitting clubs" OR "learning" OR "learning new skill" OR "learning new skills" OR "mutual aid" OR "natter club" OR "natter clubs" OR "physical activities" OR "physical activity" OR "self-help group" OR "self-help groups" OR "self-help reading" OR "signposting guidance" OR "signposting information" OR "supported education" OR "supported employment" OR "swimming" OR "team sport" OR "team sports" OR "time bank" OR "time banks" OR "volunteer" OR "volunteering" OR "volunteers" OR "walk" OR "walking" OR "walks")) AND ("Primary Health Care" OR "Primary Health Care" OR "Primary Healthcare" OR "Primary Care" OR "General Practice" OR "Family Practice" OR "General Practice" OR "General Practitioner" OR "General Practitioner" OR "General Practitioners" OR "GP" OR "Family Physician" OR "Family Physicians" OR "Physiotherapist" OR "Physical Therapist" OR "Physical Therapists" OR "Physiotherapist" OR "Physiotherapists" OR "Physiotherapy" OR "Social Worker" OR "Social Worker" OR "Social Workers" OR "Social Work" OR "Social Work" OR "health center" OR "health centres" OR "health centre" OR "health centers" OR "health center" OR "health care centres" OR "health care centre" OR "health care centers" OR "health care center" OR "healthcare centres" OR "healthcare centre" OR "healthcare centers" OR "healthcare center" OR "health services" OR "health service" OR "health care services" OR "health care service" OR "healthcare services" OR "healthcare service" OR "social care")) **OR** TI(("referral" OR "referr*" OR "prescription" OR "prescrib*" OR "prescrip*") **NEAR/1** ("adult learning" OR "aqua therapy" OR "art" OR "art therapy" OR "arts" OR "befriending" OR "bibliotherap*" OR "bibliotherapy" OR "books" OR "community education group" OR "community education groups" OR "computerised cbt" OR "computerised cognitive behavioural therapy" OR "computerized cbt" OR "computerized cognitive behavioural therapy" OR "creativity" OR "cycling" OR "dance" OR "dance class" OR "dance classes" OR "dancing" OR "education" OR "exercise" OR "exercise class" OR "exercise classes" OR "exercising " OR "fishing" OR "fishing club" OR "fishing clubs" OR "gardening" OR "gardening club" OR "gardening clubs" OR "green gym" OR "group activities" OR "group activity" OR "guided walk" OR "guided walking" OR "guided walks" OR "gym" OR "gym-based activities" OR "gym-based activity" OR "gymnastic" OR "gymnastics" OR "health walk" OR "health walks" OR "knit club" OR "knit clubs" OR "knitting" OR "knitting club" OR "knitting clubs" OR "learning" OR "learning new skill" OR "learning new skills" OR "mutual aid" OR "natter club" OR "natter clubs" OR "physical activities" OR "physical activity" OR "self-help group" OR "self-help groups" OR "self-help reading" OR "signposting guidance" OR "signposting information" OR "supported education" OR "supported employment" OR "swimming" OR "team sport" OR "team sports" OR "time bank" OR "time banks" OR "volunteer" OR "volunteering" OR "volunteers" OR "walk" OR "walking" OR "walks"))**)**

**(**NOFT("social prescribing" OR "social prescri*" OR "community prescribing" OR "community prescri*" OR "community referral" OR "community referrals" OR "community referr*" OR "social referral" OR "social referrals" OR "social referr*" OR "linking scheme" OR "linking schemes" OR "referral scheme" OR "referral schemes") **OR** NOFT(("social" **NEAR/1** "prescri*") OR ("community" **NEAR/1** "prescri*")) **OR** NOFT(("social intervention" OR "social interventions" OR "welfare intervention" OR "welfare interventions") AND ("link worker" OR "link workers" OR "link work*" OR "community navigators" OR "community navigator" OR "community liaisons" OR "community liaison" OR "well being coach" OR "wellbeing coach" OR "well being coaches" OR "wellbeing coaches" OR "welfare coordinators" OR "welfare coordinator") AND ("Primary Health Care" OR "Primary Health Care" OR "Primary Healthcare" OR "Primary Care" OR "General Practice" OR "Family Practice" OR "General Practice" OR "General Practitioner" OR "General Practitioner" OR "General Practitioners" OR "GP" OR "Family Physician" OR "Family Physicians" OR "Physiotherapist" OR "Physical Therapist" OR "Physical Therapists" OR "Physiotherapist" OR "Physiotherapists" OR "Physiotherapy" OR "Social Worker" OR "Social Worker" OR "Social Workers" OR "Social Work" OR "Social Work" OR "health center" OR "health centres" OR "health centre" OR "health centers" OR "health center" OR "health care centres" OR "health care centre" OR "health care centers" OR "health care center" OR "healthcare centres" OR "healthcare centre" OR "healthcare centers" OR "healthcare center" OR "health services" OR "health service" OR "health care services" OR "health care service" OR "healthcare services" OR "healthcare service" OR "social care")) **OR** NOFT("Arts on Prescription" OR "Books on Prescription" OR "Education on Prescription" OR "exercise on prescription" OR "exercise referral") **OR** NOFT((("referral" OR "referr*" OR "prescription" OR "prescrib*" OR "prescrip*") **NEAR/1** ("adult learning" OR "aqua therapy" OR "art" OR "art therapy" OR "arts" OR "befriending" OR "bibliotherap*" OR "bibliotherapy" OR "books" OR "community education group" OR "community education groups" OR "computerised cbt" OR "computerised cognitive behavioural therapy" OR "computerized cbt" OR "computerized cognitive behavioural therapy" OR "creativity" OR "cycling" OR "dance" OR "dance class" OR "dance classes" OR "dancing" OR "education" OR "exercise" OR "exercise class" OR "exercise classes" OR "exercising " OR "fishing" OR "fishing club" OR "fishing clubs" OR "gardening" OR "gardening club" OR "gardening clubs" OR "green gym" OR "group activities" OR "group activity" OR "guided walk" OR "guided walking" OR "guided walks" OR "gym" OR "gym-based activities" OR "gym-based activity" OR "gymnastic" OR "gymnastics" OR "health walk" OR "health walks" OR "knit club" OR "knit clubs" OR "knitting" OR "knitting club" OR "knitting clubs" OR "learning" OR "learning new skill" OR "learning new skills" OR "mutual aid" OR "natter club" OR "natter clubs" OR "physical activities" OR "physical activity" OR "self-help group" OR "self-help groups" OR "self-help reading" OR "signposting guidance" OR "signposting information" OR "supported education" OR "supported employment" OR "swimming" OR "team sport" OR "team sports" OR "time bank" OR "time banks" OR "volunteer" OR "volunteering" OR "volunteers" OR "walk" OR "walking" OR "walks")) AND ("Primary Health Care" OR "Primary Health Care" OR "Primary Healthcare" OR "Primary Care" OR "General Practice" OR "Family Practice" OR "General Practice" OR "General Practitioner" OR "General Practitioner" OR "General Practitioners" OR "GP" OR "Family Physician" OR "Family Physicians" OR "Physiotherapist" OR "Physical Therapist" OR "Physical Therapists" OR "Physiotherapist" OR "Physiotherapists" OR "Physiotherapy" OR "Social Worker" OR "Social Worker" OR "Social Workers" OR "Social Work" OR "Social Work" OR "health center" OR "health centres" OR "health centre" OR "health centers" OR "health center" OR "health care centres" OR "health care centre" OR "health care centers" OR "health care center" OR "healthcare centres" OR "healthcare centre" OR "healthcare centers" OR "healthcare center" OR "health services" OR "health service" OR "health care services" OR "health care service" OR "healthcare services" OR "healthcare service" OR "social care"))**)**

AND NOFT("review" OR "systematic review" OR "review" OR "overview" OR "review*" OR "overview*" OR "appraisal" OR "synthesis")

**Sociological Abstracts**

<https://search.proquest.com/socabs/index>

<http://databases.library.leiden.edu/?bibid=990024621960302711&redirect=true>

Limit to scholarly journals

3 queries

NOFT("social prescribing" OR "social prescri*" OR "community prescribing" OR "community prescri*" OR "community referral" OR "community referrals" OR "community referr*" OR "social referral" OR "social referrals" OR "social referr*" OR "linking scheme" OR "linking schemes" OR "referral scheme" OR "referral schemes")

**(**TI("social prescribing" OR "social prescri*" OR "community prescribing" OR "community prescri*" OR "community referral" OR "community referrals" OR "community referr*" OR "social referral" OR "social referrals" OR "social referr*" OR "linking scheme" OR "linking schemes" OR "referral scheme" OR "referral schemes") **OR** TI(("social" **NEAR/1** "prescri*") OR ("community" **NEAR/1** "prescri*")) **OR** TI(("social intervention" OR "social interventions" OR "welfare intervention" OR "welfare interventions") AND ("link worker" OR "link workers" OR "link work*" OR "community navigators" OR "community navigator" OR "community liaisons" OR "community liaison" OR "well being coach" OR "wellbeing coach" OR "well being coaches" OR "wellbeing coaches" OR "welfare coordinators" OR "welfare coordinator") AND ("Primary Health Care" OR "Primary Health Care" OR "Primary Healthcare" OR "Primary Care" OR "General Practice" OR "Family Practice" OR "General Practice" OR "General Practitioner" OR "General Practitioner" OR "General Practitioners" OR "GP" OR "Family Physician" OR "Family Physicians" OR "Physiotherapist" OR "Physical Therapist" OR "Physical Therapists" OR "Physiotherapist" OR "Physiotherapists" OR "Physiotherapy" OR "Social Worker" OR "Social Worker" OR "Social Workers" OR "Social Work" OR "Social Work" OR "health center" OR "health centres" OR "health centre" OR "health centers" OR "health center" OR "health care centres" OR "health care centre" OR "health care centers" OR "health care center" OR "healthcare centres" OR "healthcare centre" OR "healthcare centers" OR "healthcare center" OR "health services" OR "health service" OR "health care services" OR "health care service" OR "healthcare services" OR "healthcare service" OR "social care")) **OR** TI("Arts on Prescription" OR "Books on Prescription" OR "Education on Prescription" OR "exercise on prescription" OR "exercise referral") **OR** TI((("referral" OR "referr*" OR "prescription" OR "prescrib*" OR "prescrip*") **NEAR/1** ("adult learning" OR "aqua therapy" OR "art" OR "art therapy" OR "arts" OR "befriending" OR "bibliotherap*" OR "bibliotherapy" OR "books" OR "community education group" OR "community education groups" OR "computerised cbt" OR "computerised cognitive behavioural therapy" OR "computerized cbt" OR "computerized cognitive behavioural therapy" OR "creativity" OR "cycling" OR "dance" OR "dance class" OR "dance classes" OR "dancing" OR "education" OR "exercise" OR "exercise class" OR "exercise classes" OR "exercising " OR "fishing" OR "fishing club" OR "fishing clubs" OR "gardening" OR "gardening club" OR "gardening clubs" OR "green gym" OR "group activities" OR "group activity" OR "guided walk" OR "guided walking" OR "guided walks" OR "gym" OR "gym-based activities" OR "gym-based activity" OR "gymnastic" OR "gymnastics" OR "health walk" OR "health walks" OR "knit club" OR "knit clubs" OR "knitting" OR "knitting club" OR "knitting clubs" OR "learning" OR "learning new skill" OR "learning new skills" OR "mutual aid" OR "natter club" OR "natter clubs" OR "physical activities" OR "physical activity" OR "self-help group" OR "self-help groups" OR "self-help reading" OR "signposting guidance" OR "signposting information" OR "supported education" OR "supported employment" OR "swimming" OR "team sport" OR "team sports" OR "time bank" OR "time banks" OR "volunteer" OR "volunteering" OR "volunteers" OR "walk" OR "walking" OR "walks")) AND ("Primary Health Care" OR "Primary Health Care" OR "Primary Healthcare" OR "Primary Care" OR "General Practice" OR "Family Practice" OR "General Practice" OR "General Practitioner" OR "General Practitioner" OR "General Practitioners" OR "GP" OR "Family Physician" OR "Family Physicians" OR "Physiotherapist" OR "Physical Therapist" OR "Physical Therapists" OR "Physiotherapist" OR "Physiotherapists" OR "Physiotherapy" OR "Social Worker" OR "Social Worker" OR "Social Workers" OR "Social Work" OR "Social Work" OR "health center" OR "health centres" OR "health centre" OR "health centers" OR "health center" OR "health care centres" OR "health care centre" OR "health care centers" OR "health care center" OR "healthcare centres" OR "healthcare centre" OR "healthcare centers" OR "healthcare center" OR "health services" OR "health service" OR "health care services" OR "health care service" OR "healthcare services" OR "healthcare service" OR "social care")) **OR** TI(("referral" OR "referr*" OR "prescription" OR "prescrib*" OR "prescrip*") **NEAR/1** ("adult learning" OR "aqua therapy" OR "art" OR "art therapy" OR "arts" OR "befriending" OR "bibliotherap*" OR "bibliotherapy" OR "books" OR "community education group" OR "community education groups" OR "computerised cbt" OR "computerised cognitive behavioural therapy" OR "computerized cbt" OR "computerized cognitive behavioural therapy" OR "creativity" OR "cycling" OR "dance" OR "dance class" OR "dance classes" OR "dancing" OR "education" OR "exercise" OR "exercise class" OR "exercise classes" OR "exercising " OR "fishing" OR "fishing club" OR "fishing clubs" OR "gardening" OR "gardening club" OR "gardening clubs" OR "green gym" OR "group activities" OR "group activity" OR "guided walk" OR "guided walking" OR "guided walks" OR "gym" OR "gym-based activities" OR "gym-based activity" OR "gymnastic" OR "gymnastics" OR "health walk" OR "health walks" OR "knit club" OR "knit clubs" OR "knitting" OR "knitting club" OR "knitting clubs" OR "learning" OR "learning new skill" OR "learning new skills" OR "mutual aid" OR "natter club" OR "natter clubs" OR "physical activities" OR "physical activity" OR "self-help group" OR "self-help groups" OR "self-help reading" OR "signposting guidance" OR "signposting information" OR "supported education" OR "supported employment" OR "swimming" OR "team sport" OR "team sports" OR "time bank" OR "time banks" OR "volunteer" OR "volunteering" OR "volunteers" OR "walk" OR "walking" OR "walks"))**)**

**(**NOFT("social prescribing" OR "social prescri*" OR "community prescribing" OR "community prescri*" OR "community referral" OR "community referrals" OR "community referr*" OR "social referral" OR "social referrals" OR "social referr*" OR "linking scheme" OR "linking schemes" OR "referral scheme" OR "referral schemes") **OR** NOFT(("social" **NEAR/1** "prescri*") OR ("community" **NEAR/1** "prescri*")) **OR** NOFT(("social intervention" OR "social interventions" OR "welfare intervention" OR "welfare interventions") AND ("link worker" OR "link workers" OR "link work*" OR "community navigators" OR "community navigator" OR "community liaisons" OR "community liaison" OR "well being coach" OR "wellbeing coach" OR "well being coaches" OR "wellbeing coaches" OR "welfare coordinators" OR "welfare coordinator") AND ("Primary Health Care" OR "Primary Health Care" OR "Primary Healthcare" OR "Primary Care" OR "General Practice" OR "Family Practice" OR "General Practice" OR "General Practitioner" OR "General Practitioner" OR "General Practitioners" OR "GP" OR "Family Physician" OR "Family Physicians" OR "Physiotherapist" OR "Physical Therapist" OR "Physical Therapists" OR "Physiotherapist" OR "Physiotherapists" OR "Physiotherapy" OR "Social Worker" OR "Social Worker" OR "Social Workers" OR "Social Work" OR "Social Work" OR "health center" OR "health centres" OR "health centre" OR "health centers" OR "health center" OR "health care centres" OR "health care centre" OR "health care centers" OR "health care center" OR "healthcare centres" OR "healthcare centre" OR "healthcare centers" OR "healthcare center" OR "health services" OR "health service" OR "health care services" OR "health care service" OR "healthcare services" OR "healthcare service" OR "social care")) **OR** NOFT("Arts on Prescription" OR "Books on Prescription" OR "Education on Prescription" OR "exercise on prescription" OR "exercise referral") **OR** NOFT((("referral" OR "referr*" OR "prescription" OR "prescrib*" OR "prescrip*") **NEAR/1** ("adult learning" OR "aqua therapy" OR "art" OR "art therapy" OR "arts" OR "befriending" OR "bibliotherap*" OR "bibliotherapy" OR "books" OR "community education group" OR "community education groups" OR "computerised cbt" OR "computerised cognitive behavioural therapy" OR "computerized cbt" OR "computerized cognitive behavioural therapy" OR "creativity" OR "cycling" OR "dance" OR "dance class" OR "dance classes" OR "dancing" OR "education" OR "exercise" OR "exercise class" OR "exercise classes" OR "exercising " OR "fishing" OR "fishing club" OR "fishing clubs" OR "gardening" OR "gardening club" OR "gardening clubs" OR "green gym" OR "group activities" OR "group activity" OR "guided walk" OR "guided walking" OR "guided walks" OR "gym" OR "gym-based activities" OR "gym-based activity" OR "gymnastic" OR "gymnastics" OR "health walk" OR "health walks" OR "knit club" OR "knit clubs" OR "knitting" OR "knitting club" OR "knitting clubs" OR "learning" OR "learning new skill" OR "learning new skills" OR "mutual aid" OR "natter club" OR "natter clubs" OR "physical activities" OR "physical activity" OR "self-help group" OR "self-help groups" OR "self-help reading" OR "signposting guidance" OR "signposting information" OR "supported education" OR "supported employment" OR "swimming" OR "team sport" OR "team sports" OR "time bank" OR "time banks" OR "volunteer" OR "volunteering" OR "volunteers" OR "walk" OR "walking" OR "walks")) AND ("Primary Health Care" OR "Primary Health Care" OR "Primary Healthcare" OR "Primary Care" OR "General Practice" OR "Family Practice" OR "General Practice" OR "General Practitioner" OR "General Practitioner" OR "General Practitioners" OR "GP" OR "Family Physician" OR "Family Physicians" OR "Physiotherapist" OR "Physical Therapist" OR "Physical Therapists" OR "Physiotherapist" OR "Physiotherapists" OR "Physiotherapy" OR "Social Worker" OR "Social Worker" OR "Social Workers" OR "Social Work" OR "Social Work" OR "health center" OR "health centres" OR "health centre" OR "health centers" OR "health center" OR "health care centres" OR "health care centre" OR "health care centers" OR "health care center" OR "healthcare centres" OR "healthcare centre" OR "healthcare centers" OR "healthcare center" OR "health services" OR "health service" OR "health care services" OR "health care service" OR "healthcare services" OR "healthcare service" OR "social care"))**)**

AND

NOFT("review" OR "systematic review" OR "review" OR "overview" OR "review*" OR "overview*" OR "appraisal" OR "synthesis")

**ERIC**

<https://eric.ed.gov/>

Via Ebsco**:** <http://databases.library.leiden.edu/?bibid=990024848320302711&redirect=true>

Via OVID: <http://ovidsp.tx.ovid.com/sp-3.21.0a/ovidweb.cgi?&S=FKFBFPLFBNDDFEMKNCIKKBGCMKJJAA00&New+Database=Single%7c4>

Limit to Academic Journals

5 queries

(TI("social prescribing" OR "social prescri*" OR "community prescribing" OR "community prescri*" OR "community referral" OR "community referrals" OR "community referr*" OR "social referral" OR "social referrals" OR "social referr*" OR "linking scheme" OR "linking schemes" OR "referral scheme" OR "referral schemes") OR SU("social prescribing" OR "social prescri*" OR "community prescribing" OR "community prescri*" OR "community referral" OR "community referrals" OR "community referr*" OR "social referral" OR "social referrals" OR "social referr*" OR "linking scheme" OR "linking schemes" OR "referral scheme" OR "referral schemes") OR KW("social prescribing" OR "social prescri*" OR "community prescribing" OR "community prescri*" OR "community referral" OR "community referrals" OR "community referr*" OR "social referral" OR "social referrals" OR "social referr*" OR "linking scheme" OR "linking schemes" OR "referral scheme" OR "referral schemes"))

**(**TI("social prescribing" OR "social prescri*" OR "community prescribing" OR "community prescri*" OR "community referral" OR "community referrals" OR "community referr*" OR "social referral" OR "social referrals" OR "social referr*" OR "linking scheme" OR "linking schemes" OR "referral scheme" OR "referral schemes") **OR** TI(("social" **N1** "prescri*") OR ("community" **N1** "prescri*")) **OR** TI(("social intervention" OR "social interventions" OR "welfare intervention" OR "welfare interventions") AND ("link worker" OR "link workers" OR "link work*" OR "community navigators" OR "community navigator" OR "community liaisons" OR "community liaison" OR "well being coach" OR "wellbeing coach" OR "well being coaches" OR "wellbeing coaches" OR "welfare coordinators" OR "welfare coordinator") AND ("Primary Health Care" OR "Primary Health Care" OR "Primary Healthcare" OR "Primary Care" OR "General Practice" OR "Family Practice" OR "General Practice" OR "General Practitioner" OR "General Practitioner" OR "General Practitioners" OR "GP" OR "Family Physician" OR "Family Physicians" OR "Physiotherapist" OR "Physical Therapist" OR "Physical Therapists" OR "Physiotherapist" OR "Physiotherapists" OR "Physiotherapy" OR "Social Worker" OR "Social Worker" OR "Social Workers" OR "Social Work" OR "Social Work" OR "health center" OR "health centres" OR "health centre" OR "health centers" OR "health center" OR "health care centres" OR "health care centre" OR "health care centers" OR "health care center" OR "healthcare centres" OR "healthcare centre" OR "healthcare centers" OR "healthcare center" OR "health services" OR "health service" OR "health care services" OR "health care service" OR "healthcare services" OR "healthcare service" OR "social care")) **OR** TI("Arts on Prescription" OR "Books on Prescription" OR "Education on Prescription" OR "exercise on prescription" OR "exercise referral") **OR** TI((("referral" OR "referral*" OR "prescription" OR "prescrib*" OR "prescrip*") **N1** ("adult learning" OR "aqua therapy" OR "art" OR "art therapy" OR "arts" OR "befriending" OR "bibliotherap*" OR "bibliotherapy" OR "books" OR "community education group" OR "community education groups" OR "computerised cbt" OR "computerised cognitive behavioural therapy" OR "computerized cbt" OR "computerized cognitive behavioural therapy" OR "creativity" OR "cycling" OR "dance" OR "dance class" OR "dance classes" OR "dancing" OR "education" OR "exercise" OR "exercise class" OR "exercise classes" OR "exercising " OR "fishing" OR "fishing club" OR "fishing clubs" OR "gardening" OR "gardening club" OR "gardening clubs" OR "green gym" OR "group activities" OR "group activity" OR "guided walk" OR "guided walking" OR "guided walks" OR "gym" OR "gym-based activities" OR "gym-based activity" OR "gymnastic" OR "gymnastics" OR "health walk" OR "health walks" OR "knit club" OR "knit clubs" OR "knitting" OR "knitting club" OR "knitting clubs" OR "learning" OR "learning new skill" OR "learning new skills" OR "mutual aid" OR "natter club" OR "natter clubs" OR "physical activities" OR "physical activity" OR "self-help group" OR "self-help groups" OR "self-help reading" OR "signposting guidance" OR "signposting information" OR "supported education" OR "supported employment" OR "swimming" OR "team sport" OR "team sports" OR "time bank" OR "time banks" OR "volunteer" OR "volunteering" OR "volunteers" OR "walk" OR "walking" OR "walks")) AND ("Primary Health Care" OR "Primary Health Care" OR "Primary Healthcare" OR "Primary Care" OR "General Practice" OR "Family Practice" OR "General Practice" OR "General Practitioner" OR "General Practitioner" OR "General Practitioners" OR "GP" OR "Family Physician" OR "Family Physicians" OR "Physiotherapist" OR "Physical Therapist" OR "Physical Therapists" OR "Physiotherapist" OR "Physiotherapists" OR "Physiotherapy" OR "Social Worker" OR "Social Worker" OR "Social Workers" OR "Social Work" OR "Social Work" OR "health center" OR "health centres" OR "health centre" OR "health centers" OR "health center" OR "health care centres" OR "health care centre" OR "health care centers" OR "health care center" OR "healthcare centres" OR "healthcare centre" OR "healthcare centers" OR "healthcare center" OR "health services" OR "health service" OR "health care services" OR "health care service" OR "healthcare services" OR "healthcare service" OR "social care")) **OR** TI(("referral" OR "referral*" OR "prescription" OR "prescrib*" OR "prescrip*") **N1** ("adult learning" OR "aqua therapy" OR "art" OR "art therapy" OR "arts" OR "befriending" OR "bibliotherap*" OR "bibliotherapy" OR "books" OR "community education group" OR "community education groups" OR "computerised cbt" OR "computerised cognitive behavioural therapy" OR "computerized cbt" OR "computerized cognitive behavioural therapy" OR "creativity" OR "cycling" OR "dance" OR "dance class" OR "dance classes" OR "dancing" OR "education" OR "exercise" OR "exercise class" OR "exercise classes" OR "exercising " OR "fishing" OR "fishing club" OR "fishing clubs" OR "gardening" OR "gardening club" OR "gardening clubs" OR "green gym" OR "group activities" OR "group activity" OR "guided walk" OR "guided walking" OR "guided walks" OR "gym" OR "gym-based activities" OR "gym-based activity" OR "gymnastic" OR "gymnastics" OR "health walk" OR "health walks" OR "knit club" OR "knit clubs" OR "knitting" OR "knitting club" OR "knitting clubs" OR "learning" OR "learning new skill" OR "learning new skills" OR "mutual aid" OR "natter club" OR "natter clubs" OR "physical activities" OR "physical activity" OR "self-help group" OR "self-help groups" OR "self-help reading" OR "signposting guidance" OR "signposting information" OR "supported education" OR "supported employment" OR "swimming" OR "team sport" OR "team sports" OR "time bank" OR "time banks" OR "volunteer" OR "volunteering" OR "volunteers" OR "walk" OR "walking" OR "walks"))**)**

**(**SU("social prescribing" OR "social prescri*" OR "community prescribing" OR "community prescri*" OR "community referral" OR "community referrals" OR "community referr*" OR "social referral" OR "social referrals" OR "social referr*" OR "linking scheme" OR "linking schemes" OR "referral scheme" OR "referral schemes") **OR** SU(("social intervention" OR "social interventions" OR "welfare intervention" OR "welfare interventions") AND ("link worker" OR "link workers" OR "link work*" OR "community navigators" OR "community navigator" OR "community liaisons" OR "community liaison" OR "well being coach" OR "wellbeing coach" OR "well being coaches" OR "wellbeing coaches" OR "welfare coordinators" OR "welfare coordinator") AND ("Primary Health Care" OR "Primary Health Care" OR "Primary Healthcare" OR "Primary Care" OR "General Practice" OR "Family Practice" OR "General Practice" OR "General Practitioner" OR "General Practitioner" OR "General Practitioners" OR "GP" OR "Family Physician" OR "Family Physicians" OR "Physiotherapist" OR "Physical Therapist" OR "Physical Therapists" OR "Physiotherapist" OR "Physiotherapists" OR "Physiotherapy" OR "Social Worker" OR "Social Worker" OR "Social Workers" OR "Social Work" OR "Social Work" OR "health center" OR "health centres" OR "health centre" OR "health centers" OR "health center" OR "health care centres" OR "health care centre" OR "health care centers" OR "health care center" OR "healthcare centres" OR "healthcare centre" OR "healthcare centers" OR "healthcare center" OR "health services" OR "health service" OR "health care services" OR "health care service" OR "healthcare services" OR "healthcare service" OR "social care")) **OR** SU("Arts on Prescription" OR "Books on Prescription" OR "Education on Prescription" OR "exercise on prescription" OR "exercise referral") **OR** SU((("referral" OR "referral*" OR "prescription" OR "prescrib*" OR "prescrip*") **N1** ("adult learning" OR "aqua therapy" OR "art" OR "art therapy" OR "arts" OR "befriending" OR "bibliotherap*" OR "bibliotherapy" OR "books" OR "community education group" OR "community education groups" OR "computerised cbt" OR "computerised cognitive behavioural therapy" OR "computerized cbt" OR "computerized cognitive behavioural therapy" OR "creativity" OR "cycling" OR "dance" OR "dance class" OR "dance classes" OR "dancing" OR "education" OR "exercise" OR "exercise class" OR "exercise classes" OR "exercising " OR "fishing" OR "fishing club" OR "fishing clubs" OR "gardening" OR "gardening club" OR "gardening clubs" OR "green gym" OR "group activities" OR "group activity" OR "guided walk" OR "guided walking" OR "guided walks" OR "gym" OR "gym-based activities" OR "gym-based activity" OR "gymnastic" OR "gymnastics" OR "health walk" OR "health walks" OR "knit club" OR "knit clubs" OR "knitting" OR "knitting club" OR "knitting clubs" OR "learning" OR "learning new skill" OR "learning new skills" OR "mutual aid" OR "natter club" OR "natter clubs" OR "physical activities" OR "physical activity" OR "self-help group" OR "self-help groups" OR "self-help reading" OR "signposting guidance" OR "signposting information" OR "supported education" OR "supported employment" OR "swimming" OR "team sport" OR "team sports" OR "time bank" OR "time banks" OR "volunteer" OR "volunteering" OR "volunteers" OR "walk" OR "walking" OR "walks")) AND ("Primary Health Care" OR "Primary Health Care" OR "Primary Healthcare" OR "Primary Care" OR "General Practice" OR "Family Practice" OR "General Practice" OR "General Practitioner" OR "General Practitioner" OR "General Practitioners" OR "GP" OR "Family Physician" OR "Family Physicians" OR "Physiotherapist" OR "Physical Therapist" OR "Physical Therapists" OR "Physiotherapist" OR "Physiotherapists" OR "Physiotherapy" OR "Social Worker" OR "Social Worker" OR "Social Workers" OR "Social Work" OR "Social Work" OR "health center" OR "health centres" OR "health centre" OR "health centers" OR "health center" OR "health care centres" OR "health care centre" OR "health care centers" OR "health care center" OR "healthcare centres" OR "healthcare centre" OR "healthcare centers" OR "healthcare center" OR "health services" OR "health service" OR "health care services" OR "health care service" OR "healthcare services" OR "healthcare service" OR "social care"))**)**

**(**KW("social prescribing" OR "social prescri*" OR "community prescribing" OR "community prescri*" OR "community referral" OR "community referrals" OR "community referr*" OR "social referral" OR "social referrals" OR "social referr*" OR "linking scheme" OR "linking schemes" OR "referral scheme" OR "referral schemes") **OR** KW(("social intervention" OR "social interventions" OR "welfare intervention" OR "welfare interventions") AND ("link worker" OR "link workers" OR "link work*" OR "community navigators" OR "community navigator" OR "community liaisons" OR "community liaison" OR "well being coach" OR "wellbeing coach" OR "well being coaches" OR "wellbeing coaches" OR "welfare coordinators" OR "welfare coordinator") AND ("Primary Health Care" OR "Primary Health Care" OR "Primary Healthcare" OR "Primary Care" OR "General Practice" OR "Family Practice" OR "General Practice" OR "General Practitioner" OR "General Practitioner" OR "General Practitioners" OR "GP" OR "Family Physician" OR "Family Physicians" OR "Physiotherapist" OR "Physical Therapist" OR "Physical Therapists" OR "Physiotherapist" OR "Physiotherapists" OR "Physiotherapy" OR "Social Worker" OR "Social Worker" OR "Social Workers" OR "Social Work" OR "Social Work" OR "health center" OR "health centres" OR "health centre" OR "health centers" OR "health center" OR "health care centres" OR "health care centre" OR "health care centers" OR "health care center" OR "healthcare centres" OR "healthcare centre" OR "healthcare centers" OR "healthcare center" OR "health services" OR "health service" OR "health care services" OR "health care service" OR "healthcare services" OR "healthcare service" OR "social care")) **OR** KW("Arts on Prescription" OR "Books on Prescription" OR "Education on Prescription" OR "exercise on prescription" OR "exercise referral") **OR** KW((("referral" OR "referral*" OR "prescription" OR "prescrib*" OR "prescrip*") **N1** ("adult learning" OR "aqua therapy" OR "art" OR "art therapy" OR "arts" OR "befriending" OR "bibliotherap*" OR "bibliotherapy" OR "books" OR "community education group" OR "community education groups" OR "computerised cbt" OR "computerised cognitive behavioural therapy" OR "computerized cbt" OR "computerized cognitive behavioural therapy" OR "creativity" OR "cycling" OR "dance" OR "dance class" OR "dance classes" OR "dancing" OR "education" OR "exercise" OR "exercise class" OR "exercise classes" OR "exercising " OR "fishing" OR "fishing club" OR "fishing clubs" OR "gardening" OR "gardening club" OR "gardening clubs" OR "green gym" OR "group activities" OR "group activity" OR "guided walk" OR "guided walking" OR "guided walks" OR "gym" OR "gym-based activities" OR "gym-based activity" OR "gymnastic" OR "gymnastics" OR "health walk" OR "health walks" OR "knit club" OR "knit clubs" OR "knitting" OR "knitting club" OR "knitting clubs" OR "learning" OR "learning new skill" OR "learning new skills" OR "mutual aid" OR "natter club" OR "natter clubs" OR "physical activities" OR "physical activity" OR "self-help group" OR "self-help groups" OR "self-help reading" OR "signposting guidance" OR "signposting information" OR "supported education" OR "supported employment" OR "swimming" OR "team sport" OR "team sports" OR "time bank" OR "time banks" OR "volunteer" OR "volunteering" OR "volunteers" OR "walk" OR "walking" OR "walks")) AND ("Primary Health Care" OR "Primary Health Care" OR "Primary Healthcare" OR "Primary Care" OR "General Practice" OR "Family Practice" OR "General Practice" OR "General Practitioner" OR "General Practitioner" OR "General Practitioners" OR "GP" OR "Family Physician" OR "Family Physicians" OR "Physiotherapist" OR "Physical Therapist" OR "Physical Therapists" OR "Physiotherapist" OR "Physiotherapists" OR "Physiotherapy" OR "Social Worker" OR "Social Worker" OR "Social Workers" OR "Social Work" OR "Social Work" OR "health center" OR "health centres" OR "health centre" OR "health centers" OR "health center" OR "health care centres" OR "health care centre" OR "health care centers" OR "health care center" OR "healthcare centres" OR "healthcare centre" OR "healthcare centers" OR "healthcare center" OR "health services" OR "health service" OR "health care services" OR "health care service" OR "healthcare services" OR "healthcare service" OR "social care"))**)**

TI((("referral" OR "referral*" OR "prescription" OR "prescrib*" OR "prescrip*") **N1** ("adult learning" OR "aqua therapy" OR "art" OR "art therapy" OR "arts" OR "befriending" OR "bibliotherap*" OR "bibliotherapy" OR "books" OR "community education group" OR "community education groups" OR "computerised cbt" OR "computerised cognitive behavioural therapy" OR "computerized cbt" OR "computerized cognitive behavioural therapy" OR "creativity" OR "cycling" OR "dance" OR "dance class" OR "dance classes" OR "dancing" OR "education" OR "exercise" OR "exercise class" OR "exercise classes" OR "exercising " OR "fishing" OR "fishing club" OR "fishing clubs" OR "gardening" OR "gardening club" OR "gardening clubs" OR "green gym" OR "group activities" OR "group activity" OR "guided walk" OR "guided walking" OR "guided walks" OR "gym" OR "gym-based activities" OR "gym-based activity" OR "gymnastic" OR "gymnastics" OR "health walk" OR "health walks" OR "knit club" OR "knit clubs" OR "knitting" OR "knitting club" OR "knitting clubs" OR "learning" OR "learning new skill" OR "learning new skills" OR "mutual aid" OR "natter club" OR "natter clubs" OR "physical activities" OR "physical activity" OR "self-help group" OR "self-help groups" OR "self-help reading" OR "signposting guidance" OR "signposting information" OR "supported education" OR "supported employment" OR "swimming" OR "team sport" OR "team sports" OR "time bank" OR "time banks" OR "volunteer" OR "volunteering" OR "volunteers" OR "walk" OR "walking" OR "walks")) AND ("Primary Health Care" OR "Primary Health Care" OR "Primary Healthcare" OR "Primary Care" OR "General Practice" OR "Family Practice" OR "General Practice" OR "General Practitioner" OR "General Practitioner" OR "General Practitioners" OR "GP" OR "Family Physician" OR "Family Physicians" OR "Physiotherapist" OR "Physical Therapist" OR "Physical Therapists" OR "Physiotherapist" OR "Physiotherapists" OR "Physiotherapy" OR "Social Worker" OR "Social Worker" OR "Social Workers" OR "Social Work" OR "Social Work" OR "health center" OR "health centres" OR "health centre" OR "health centers" OR "health center" OR "health care centres" OR "health care centre" OR "health care centers" OR "health care center" OR "healthcare centres" OR "healthcare centre" OR "healthcare centers" OR "healthcare center" OR "health services" OR "health service" OR "health care services" OR "health care service" OR "healthcare services" OR "healthcare service" OR "social care"))

AND

(TI("review" OR "systematic review" OR "review" OR "overview" OR "review*" OR "overview*" OR "appraisal" OR "synthesis") OR SU("review" OR "systematic review" OR "review" OR "overview" OR "review*" OR "overview*" OR "appraisal" OR "synthesis") OR KW("review" OR "systematic review" OR "review" OR "overview" OR "review*" OR "overview*" OR "appraisal" OR "synthesis") OR AB("review" OR "systematic review" OR "review" OR "overview" OR "review*" OR "overview*" OR "appraisal" OR "synthesis"))

**NHS economic evaluation database + Health Technology Assessment database**

<https://www.crd.york.ac.uk/CRDWeb/>

4 queries

("social prescribing" OR "social prescri*" OR "community prescribing" OR "community prescri*" OR "community referral" OR "community referrals" OR "community referr*" OR "social referral" OR "social referrals" OR "social referr*" OR "linking scheme" OR "linking schemes" OR "referral scheme" OR "referral schemes")

(("social intervention" OR "social interventions" OR "welfare intervention" OR "welfare interventions") AND ("link worker" OR "link workers" OR "link work*" OR "community navigators" OR "community navigator" OR "community liaisons" OR "community liaison" OR "well being coach" OR "wellbeing coach" OR "well being coaches" OR "wellbeing coaches" OR "welfare coordinators" OR "welfare coordinator") AND ("Primary Health Care" OR "Primary Health Care" OR "Primary Healthcare" OR "Primary Care" OR "General Practice" OR "Family Practice" OR "General Practice" OR "General Practitioner" OR "General Practitioner" OR "General Practitioners" OR "GP" OR "Family Physician" OR "Family Physicians" OR "Physiotherapist" OR "Physical Therapist" OR "Physical Therapists" OR "Physiotherapist" OR "Physiotherapists" OR "Physiotherapy" OR "Social Worker" OR "Social Worker" OR "Social Workers" OR "Social Work" OR "Social Work" OR "health center" OR "health centres" OR "health centre" OR "health centers" OR "health center" OR "health care centres" OR "health care centre" OR "health care centers" OR "health care center" OR "healthcare centres" OR "healthcare centre" OR "healthcare centers" OR "healthcare center" OR "health services" OR "health service" OR "health care services" OR "health care service" OR "healthcare services" OR "healthcare service" OR "social care"))

**OR**

("Arts on Prescription" OR "Books on Prescription" OR "Education on Prescription" OR "exercise on prescription" OR "exercise referral")

**OR**

Title

(("referral" OR "referral*" OR "prescription" OR "prescrib*" OR "prescrip*") **AND** ("adult learning" OR "aqua therapy" OR "art" OR "art therapy" OR "arts" OR "befriending" OR "bibliotherap*" OR "bibliotherapy" OR "books" OR "community education group" OR "community education groups" OR "computerised cbt" OR "computerised cognitive behavioural therapy" OR "computerized cbt" OR "computerized cognitive behavioural therapy" OR "creativity" OR "cycling" OR "dance" OR "dance class" OR "dance classes" OR "dancing" OR "education" OR "exercise" OR "exercise class" OR "exercise classes" OR "exercising " OR "fishing" OR "fishing club" OR "fishing clubs" OR "gardening" OR "gardening club" OR "gardening clubs" OR "green gym" OR "group activities" OR "group activity" OR "guided walk" OR "guided walking" OR "guided walks" OR "gym" OR "gym-based activities" OR "gym-based activity" OR "gymnastic" OR "gymnastics" OR "health walk" OR "health walks" OR "knit club" OR "knit clubs" OR "knitting" OR "knitting club" OR "knitting clubs" OR "learning" OR "learning new skill" OR "learning new skills" OR "mutual aid" OR "natter club" OR "natter clubs" OR "physical activities" OR "physical activity" OR "self-help group" OR "self-help groups" OR "self-help reading" OR "signposting guidance" OR "signposting information" OR "supported education" OR "supported employment" OR "swimming" OR "team sport" OR "team sports" OR "time bank" OR "time banks" OR "volunteer" OR "volunteering" OR "volunteers" OR "walk" OR "walking" OR "walks") AND ("Primary Health Care" OR "Primary Health Care" OR "Primary Healthcare" OR "Primary Care" OR "General Practice" OR "Family Practice" OR "General Practice" OR "General Practitioner" OR "General Practitioner" OR "General Practitioners" OR "GP" OR "Family Physician" OR "Family Physicians" OR "Physiotherapist" OR "Physical Therapist" OR "Physical Therapists" OR "Physiotherapist" OR "Physiotherapists" OR "Physiotherapy" OR "Social Worker" OR "Social Worker" OR "Social Workers" OR "Social Work" OR "Social Work" OR "health center" OR "health centres" OR "health centre" OR "health centers" OR "health center" OR "health care centres" OR "health care centre" OR "health care centers" OR "health care center" OR "healthcare centres" OR "healthcare centre" OR "healthcare centers" OR "healthcare center" OR "health services" OR "health service" OR "health care services" OR "health care service" OR "healthcare services" OR "healthcare service" OR "social care"))
