## AppendixC_BCTTaxonomy_v1 for "Intervention Characteristics and Mechanisms and their Relationship with the Influence of Social Prescribing: a Systematic Review"

BCT Taxonomy (v1): 93 hierarchically-clustered techniques


| Page | Grouping and BCTs |
| --- | --- |
| 1 | **1. Goals and planning**<br>1.1. Goal setting (behavior)<br>1.2. Problem solving<br>1.3. Goal setting (outcome)<br>1.4. Action planning<br>1.5. Review behavior goal(s)<br>1.6. Discrepancy between current behavior and goal<br>1.7. Review outcome goal(s)<br>1.8. Behavioral contract<br>1.9. Commitment |
| 3 | **2. Feedback and monitoring**<br>2.1. Monitoring of behavior by others without feedback<br>2.2. Feedback on behaviour<br>2.3. Self-monitoring of behaviour<br>2.4. Self-monitoring of outcome(s) of behaviour<br>2.5. Monitoring of outcome(s) of behavior without feedback<br>2.6. Biofeedback<br>2.7. Feedback on outcome(s) of behavior |
| 5 | **3. Social support**<br>3.1. Social support (unspecified)<br>3.2. Social support (practical)<br>3.3. Social support (emotional) |
| 6 | **4. Shaping knowledge**<br>4.1. Instruction on how to perform the behavior<br>4.2. Information about Antecedents<br>4.3. Re-attribution<br>4.4. Behavioral experiments |
| 7 | **5. Natural consequences**<br>5.1. Information about health consequences<br>5.2. Salience of consequences<br>5.3. Information about social and environmental consequences<br>5.4. Monitoring of emotional consequences<br>5.5. Anticipated regret<br>5.6. Information about emotional consequences |
| 8 | **6. Comparison of behaviour**<br>6.1. Demonstration of the behavior<br>6.2. Social comparison<br>6.3. Information about others' approval |
| 9 | **7. Associations**<br>7.1. Prompts/cues<br>7.2. Cue signalling reward<br>7.3. Reduce prompts/cues<br>7.4. Remove access to the reward<br>7.5. Remove aversive stimulus<br>7.6. Satiation<br>7.7. Exposure<br>7.8. Associative learning |
| 10 | **8. Repetition and substitution**<br>8.1. Behavioral practice/rehearsal<br>8.2. Behavior substitution<br>8.3. Habit formation<br>8.4. Habit reversal<br>8.5. Overcorrection<br>8.6. Generalisation of target behavior<br>8.7. Graded tasks |
| 11 | **9. Comparison of outcomes**<br>9.1. Credible source<br>9.2. Pros and cons<br>9.3. Comparative imagining of future outcomes |
| 12 | **10. Reward and threat**<br>10.1. Material incentive (behavior)<br>10.2. Material reward (behavior)<br>10.3. Non-specific reward<br>10.4. Social reward<br>10.5. Social incentive<br>10.6. Non-specific incentive<br>10.7. Self-incentive<br>10.8. Incentive (outcome)<br>10.9. Self-reward<br>10.10. Reward (outcome)<br>10.11. Future punishment |
| 15 | **11. Regulation**<br>11.1. Pharmacological support<br>11.2. Reduce negative emotions<br>11.3. Conserving mental resources<br>11.4. Paradoxical instructions |
| 16 | **12. Antecedents**<br>12.1. Restructuring the physical environment<br>12.2. Restructuring the social environment<br>12.3. Avoidance/reducing exposure to cues for the behavior<br>12.4. Distraction<br>12.5. Adding objects to the environment<br>12.6. Body changes |
| 17 | **13. Identity**<br>13.1. Identification of self as role model<br>13.2. Framing/reframing<br>13.3. Incompatible beliefs<br>13.4. Valued self-identify<br>13.5. Identity associated with changed behavior |
| 18 | **14. Scheduled consequences**<br>14.1. Behavior cost<br>14.2. Punishment<br>14.3. Remove reward<br>14.4. Reward approximation<br>14.5. Rewarding completion<br>14.6. Situation-specific reward<br>14.7. Reward incompatible behavior<br>14.8. Reward alternative behavior<br>14.9. Reduce reward frequency<br>14.10. Remove punishment |
| 19 | **15. Self-belief**<br>15.1. Verbal persuasion about capability<br>15.2. Mental rehearsal of successful performance<br>15.3. Focus on past success<br>15.4. Self-talk |
| 19 | **16. Covert learning**<br>16.1. Imaginary punishment<br>16.2. Imaginary reward<br>16.3. Vicarious consequences |

**BCT Taxonomy (v1): 93 hierarchically-clustered techniques**

**Note for Users**

**The definitions of Behavior Change Techniques (BCTs):**

i) contain verbs (e.g., provide, advise, arrange, prompt) that refer to the action(s) taken by the person/s delivering the technique. BCTs can be delivered by an 'interventionist' or self-delivered
ii) contain the term **"behavior"** referring to a single action or sequence of actions that includes the performance of **wanted** behavior(s) and/or **inhibition** (non-performance) of **unwanted** behavior(s)
iii) note alternative or additional coding where relevant
iv) note the technical terms associated with particular theoretical frameworks where relevant (e.g. 'including implementation intentions)

| No. | Label | Definition | Examples |
|---|---|---|---|
| **1. Goals and planning** | | | |
| **1.1** | ***Goal setting (behavior)*** | Set or agree on a goal defined in terms of the behavior to be achieved<br>*Note: only code goal-setting if there is sufficient evidence that goal set as part of intervention; if goal unspecified or a behavioral outcome, code **1.3, Goal setting (outcome)**; if the goal defines a specific context, frequency, duration or intensity for the behavior, also code **1.4, Action planning*** | Agree on a daily walking goal (e.g. 3 miles) with the person and reach agreement about the goal<br><br>Set the goal of eating 5 pieces of fruit per day as specified in public health guidelines |
| **1.2** | ***Problem solving*** | Analyse , or prompt the person to analyse, factors influencing the behavior and generate or select strategies that include overcoming barriers and/or increasing facilitators (includes '**Relapse Prevention**' *and* '**Coping Planning**')<br>*Note: barrier identification without solutions is not sufficient. If the BCT does not include analysing the behavioral problem, consider **12.3, Avoidance/changing exposure to cues for the behavior, 12.1, Restructuring the physical environment, 12.2, Restructuring the social environment,** or **11.2, Reduce negative emotions*** | Identify specific triggers (e.g. being in a pub, feeling anxious) that generate the urge/want/need to drink and develop strategies for avoiding environmental triggers or for managing negative emotions, such as anxiety, that motivate drinking<br><br>Prompt the patient to identify barriers preventing them from starting a new exercise regime e.g., lack of motivation, and discuss ways in which they could help overcome them e.g., going to the gym with a buddy |

Back to index page

| | | | |
|---|---|---|---|
| **1.3** | ***Goal setting (outcome)*** | Set or agree on a goal defined in terms of a positive **outcome** of wanted behavior<br>*Note: only code guidelines if set as a goal in an intervention context; if goal is a behavior, code **1.1, Goal setting (behavior)**; if goal unspecified code **1.3, Goal setting (outcome)*** | Set a weight loss goal (e.g. 0.5 kilogram over one week) as an outcome of changed eating patterns |
| **1.4** | ***Action planning*** | Prompt detailed planning of performance of the behavior (must include at least one of context, frequency, duration and intensity). Context may be environmental (physical or social) or internal (physical, emotional or cognitive) (includes *'***Implementation Intentions***'*)<br>*Note: evidence of action planning does not necessarily imply goal setting, only code latter if sufficient evidence* | Encourage a plan to carry condoms when going out socially at weekends<br><br>Prompt planning the performance of a particular physical activity (e.g. running) at a particular time (e.g. before work) on certain days of the week |
| **1.5** | ***Review behavior goal(s)*** | Review behavior goal(s) jointly with the person and consider modifying goal(s) or behavior change strategy in light of achievement. This may lead to re-setting the same goal, a small change in that goal or setting a new goal instead of (or in addition to) the first, or no change<br>*Note: if goal specified in terms of behavior, code **1.5, Review behavior goal(s)**, if goal unspecified, code **1.7, Review outcome goal(s);** if discrepancy created consider also **1.6, Discrepancy between current behavior and goal*** | Examine how well a person's performance corresponds to agreed goals e.g. whether they consumed less than one unit of alcohol per day, and consider modifying future behavioral goals accordingly e.g. by increasing or decreasing alcohol target or changing type of alcohol consumed |
| **1.6** | ***Discrepancy between current behavior and goal*** | Draw attention to discrepancies between a person's current behavior (in terms of the *form, frequency, duration, or intensity* of that behavior) and the person's previously set outcome goals, behavioral goals or action plans (goes beyond self-monitoring of behavior)<br>*Note: if discomfort is created only code **13.3, Incompatible beliefs** and <u>not</u> **1.6, Discrepancy between current behavior and goal**; if goals are modified, also code **1.5, Review behavior goal(s)** and/or **1.7, Review outcome goal(s)**; if feedback is provided, <u>also</u> code **2.2, Feedback on behaviour*** | Point out that the recorded exercise fell short of the goal set |

Back to index page

| | | | |
|---|---|---|---|
| **1.7** | ***Review outcome goal(s)*** | Review outcome goal(s) jointly with the person and consider modifying goal(s) in light of achievement. This may lead to re-setting the same goal, a small change in that goal or setting a new goal instead of, or in addition to the first<br>*Note: if goal specified in terms of behavior, code* ***1.5, Review behavior goal(s)****, if goal unspecified, code* ***1.7, Review outcome goal(s);*** *if discrepancy created consider also* ***1.6, Discrepancy between current behavior and goal*** | Examine how much weight has been lost and consider modifying outcome goal(s) accordingly e.g., by increasing or decreasing subsequent weight loss targets |
| **1.8** | ***Behavioral contract*** | Create a written specification of the behavior to be performed, agreed on by the person, and witnessed by another<br>*Note: also code* ***1.1, Goal setting (behavior)*** | Sign a contract with the person e.g. specifying that they will not drink alcohol for one week |
| **1.9** | ***Commitment*** | Ask the person to affirm or reaffirm statements indicating commitment to change the behavior<br>*Note: if defined in terms of the behavior to be achieved also code* ***1.1, Goal setting (behavior)*** | Ask the person to use an "I will" statement to affirm or reaffirm a strong commitment (i.e. using the words "strongly", "committed" or "high priority") to start, continue or restart the attempt to take medication as prescribed |
| **2. Feedback and monitoring** | | | |
| **2.1** | ***Monitoring of behavior by others without feedback*** | Observe or record behavior with the person's knowledge as part of a behavior change strategy<br>*Note: if monitoring is part of a data collection procedure rather than a strategy aimed at changing behavior, do not code; if feedback given, code only* ***2.2, Feedback on behavior****, and not* ***2.1, Monitoring of behavior by others without feedback****; if monitoring outcome(s) code* ***2.5, Monitoring outcome(s) of behavior by others without feedback****; if self-monitoring behavior, code* ***2.3, Self-monitoring of behaviour*** | Watch hand washing behaviors among health care staff and make notes on context, frequency and technique used |
| Back to index page | | | |

| | | | |
|---|---|---|---|
| **2.2** | ***Feedback on behavior*** | Monitor and provide informative or evaluative feedback on performance of the behavior *(e.g. form, frequency, duration, intensity)*<br>*Note: if Biofeedback, code only* ***2.6, Biofeedback*** *and not* ***2.2, Feedback on behavior****; if feedback is on* ***outcome(s)*** *of behavior, code* ***2.7, Feedback on outcome(s) of behavior****; if there is no clear evidence that feedback was given, code* ***2.1, Monitoring of behavior by others without feedback****; if feedback on behaviour is evaluative e.g. praise, also code* ***10.4, Social reward*** | Inform the person of how many steps they walked each day (as recorded on a pedometer) or how many calories they ate each day (based on a food consumption questionnaire). |
| **2.3** | ***Self-monitoring of behavior*** | Establish a method for the person to monitor and record their behavior(s) as part of a behavior change strategy<br>*Note: if monitoring is part of a data collection procedure rather than a strategy aimed at changing behavior, do not code; if monitoring of outcome of behavior, code* ***2.4, Self-monitoring of outcome(s) of behavior****; if monitoring is by someone else (without feedback), code* ***2.1, Monitoring of behavior by others without feedback*** | Ask the person to record daily, in a diary, whether they have brushed their teeth for at least two minutes before going to bed<br><br>Give patient a pedometer and a form for recording daily total number of steps |
| **2.4** | ***Self-monitoring of outcome(s) of behavior*** | Establish a method for the person to monitor and record the **outcome(s)** of their behavior as part of a behavior change strategy<br>*Note: if monitoring is part of a data collection procedure rather than a strategy aimed at changing behavior, do not code ; if monitoring behavior, code* ***2.3, Self-monitoring of behavior****; if monitoring is by someone else (without feedback), code* ***2.5, Monitoring outcome(s) of behavior by others without feedback*** | Ask the person to weigh themselves at the end of each day, over a two week period, and record their daily weight on a graph to increase exercise behaviors |

Back to index page

| | | | |
|---|---|---|---|
| **2.5** | ***Monitoring outcome(s) of behavior by others without feedback*** | Observe or record outcomes of behavior with the person's knowledge as part of a behavior change strategy<br>*Note: if monitoring is part of a data collection procedure rather than a strategy aimed at changing behavior, do not code; if feedback given, code only* ***2.7, Feedback on outcome(s) of behavior****; if monitoring behavior code* ***2.1, Monitoring of behavior by others without feedback****; if self-monitoring outcome(s), code* ***2.4, Self-monitoring of outcome(s) of behavior*** | Record blood pressure, blood glucose, weight loss, or physical fitness |
| **2.6** | ***Biofeedback*** | Provide feedback about the body *(e.g. physiological or biochemical state)* using an external monitoring device as part of a behavior change strategy<br>*Note: if Biofeedback, code only* ***2.6, Biofeedback*** *and <u>not</u>* ***2.2, Feedback on behavior*** *or* ***2.7, Feedback on outcome(s) of behaviour*** | Inform the person of their blood pressure reading to improve adoption of health behaviors |
| **2.7** | ***Feedback on outcome(s) of behavior*** | Monitor and provide feedback on the outcome of performance of the behavior<br>*Note: if Biofeedback, code only* ***2.6, Biofeedback*** *and <u>not</u>* ***2.7, Feedback on outcome(s) of behavior****; if feedback is on* ***behavior*** *code* ***2.2, Feedback on behavior****; if there is no clear evidence that feedback was given code* ***2.5, Monitoring outcome(s) of behavior by others without feedback****; if feedback on behaviour is evaluative e.g. praise, also code* ***10.4, Social reward*** | Inform the person of how much weight they have lost following the implementation of a new exercise regime |
| **3. Social support** | | | |
| **3.1** | ***Social support (unspecified)*** | Advise on, arrange or provide social support *(e.g. from friends, relatives, colleagues,' buddies' or staff)* or non-contingent praise or reward for performance of the behavior. It includes encouragement and counselling, but only when it is directed at the **behavior**<br>*Note: attending a group class and/or mention of 'follow-up' does not necessarily apply this BCT, support must be explicitly mentioned; if practical, code* ***3.2, Social support (practical)****; if emotional, code* ***3.3, Social support (emotional)*** (includes '**<u>Motivational interviewing</u>**' and **<u>'Cognitive Behavioral Therapy'</u>**) | Advise the person to call a 'buddy' when they experience an urge to smoke<br><br>Arrange for a housemate to encourage continuation with the behavior change programme<br><br>Give information about a self-help group that offers support for the behaviour<br><br>Back to index page |

| | | | |
|---|---|---|---|
| **3.2** | ***Social support (practical)*** | Advise on, arrange, or provide **practical** help *(e.g. from friends, relatives, colleagues, 'buddies' or staff)* for performance of the behavior<br>*Note: if emotional, code **3.3, Social support (emotional)**; if general or unspecified, code **3.1, Social support (unspecified)** If only restructuring the physical environment or adding objects to the environment, code **12.1, Restructuring the physical environment** or **12.5, Adding objects to the environment;** attending a group or class and/or mention of 'follow-up' does not necessarily apply this BCT, support must be explicitly mentioned.* | Ask the partner of the patient to put their tablet on the breakfast tray so that the patient remembers to take it |
| **3.3** | ***Social support (emotional)*** | Advise on, arrange, or provide **emotional** social support *(e.g. from friends, relatives, colleagues, 'buddies' or staff)* for performance of the behavior<br>*Note: if practical, code **3.2, Social support (practical)**; if unspecified, code **3.1, Social support (unspecified)*** | Ask the patient to take a partner or friend with them to their colonoscopy appointment |
| **4. Shaping knowledge** | | | |
| **4.1** | ***Instruction on how to perform a behavior*** | Advise or agree on how to perform the behavior (includes '**Skills training**')<br>*Note: when the person attends classes such as exercise or cookery, code **4.1, Instruction on how to perform the behavior, 8.1, Behavioral practice/rehearsal** and **6.1, Demonstration of the behavior*** | Advise the person how to put a condom on a model of a penis correctly |
| **4.2** | ***Information about antecedents*** | Provide information about antecedents (*e.g. social and environmental situations and events, emotions, cognitions)* that reliably predict performance of the behaviour | Advise to keep a record of snacking and of situations or events occurring prior to snacking |
| **4.3** | ***Re-attribution*** | Elicit perceived causes of behavior and suggest alternative explanations *(e.g. external or internal and stable or unstable)* | If the person attributes their over-eating to the frequent presence of delicious food, suggest that the 'real' cause may be the person's inattention to bodily signals of hunger and satiety |

Back to index page

| | | | |
|---|---|---|---|
| **4.4** | ***Behavioral experiments*** | Advise on how to identify and test hypotheses about the behavior, its causes and consequences, by collecting and interpreting data | Ask a family physician to give evidence-based advice rather than prescribe antibiotics and to note whether the patients are grateful or annoyed |
| **5. Natural consequences** | | | |
| **5.1** | ***Information about health consequences*** | Provide information (e.g. written, verbal, visual) about health consequences of performing the behavior<br>*Note: consequences can be for any target, not just the recipient(s) of the intervention; emphasising importance of consequences is not sufficient; if information about emotional consequences, code* ***5.6, Information about emotional consequences****; if about social, environmental or unspecified consequences code* ***5.3, Information about social and environmental consequences*** | Explain that not finishing a course of antibiotics can increase susceptibility to future infection<br><br>Present the likelihood of contracting a sexually transmitted infection following unprotected sexual behavior |
| **5.2** | ***Salience of consequences*** | Use methods specifically designed to **emphasise** the consequences of performing the behaviour with the aim of making them more memorable (goes beyond informing about consequences)<br>*Note: if information about consequences, also code* ***5.1, Information about health consequences****,* ***5.6, Information about emotional consequences*** *or* ***5.3, Information about social and environmental consequences*** | Produce cigarette packets showing pictures of health consequences e.g. diseased lungs, to highlight the dangers of continuing to smoke |
| **5.3** | ***Information about social and environmental consequences*** | Provide information (e.g. written, verbal, visual) about social and environmental consequences of performing the behavior<br>*Note: consequences can be for any target, not just the recipient(s) of the intervention; if information about health or consequences, code* ***5.1, Information about health consequences****; if about emotional consequences, code* ***5.6, Information about emotional consequences****; if unspecified, code* ***5.3, Information about social and environmental consequences*** | Tell family physician about financial remuneration for conducting health screening<br><br>Inform a smoker that the majority of people disapprove of smoking in public places |
| **5.4** | ***Monitoring of emotional consequences*** | Prompt assessment of **feelings** after attempts at performing the behavior | Agree that the person will record how they feel after taking their daily walk |

Back to index page

| | | | |
|---|---|---|---|
| **5.5** | ***Anticipated regret*** | Induce or raise awareness of expectations of future regret about performance of the unwanted behavior<br>*Note: not including **5.6, Information about emotional consequences**; if suggests adoption of a perspective or new perspective in order to change cognitions also code **13.2, Framing/reframing*** | Ask the person to assess the degree of regret they will feel if they do not quit smoking |
| **5.6** | ***Information about emotional consequences*** | Provide information (e.g. written, verbal, visual) about emotional consequences of performing the behavior<br>*Note: consequences can be related to emotional health disorders (e.g. depression, anxiety) and/or states of mind (e.g. low mood, stress); not including **5.5, Anticipated regret**; consequences can be for any target, not just the recipient(s) of the intervention; if information about health consequences code **5.1, Information about health consequences**; if about social, environmental or unspecified code **5.3, Information about social and environmental consequences*** | Explain that quitting smoking increases happiness and life satisfaction |
| **6. Comparison of behaviour** | | | |
| **6.1** | ***Demonstration of the behavior*** | Provide an observable sample of the performance of the behaviour, directly in person or indirectly e.g. via film, pictures, for the person to aspire to or imitate (includes '**Modelling**'). *Note:* if advised to practice, also code, ***8.1, Behavioural practice and rehearsal;*** *If provided with instructions on how to perform, also code **4.1, Instruction on how to perform the behaviour*** | Demonstrate to nurses how to raise the issue of excessive drinking with patients via a role-play exercise |
| **6.2** | ***Social comparison*** | Draw attention to others' performance to allow comparison with the person's own performance *Note: being in a group setting does not necessarily mean that social comparison is actually taking place* | Show the doctor the proportion of patients who were prescribed antibiotics for a common cold by other doctors and compare with their own data |
| **6.3** | ***Information about others' approval*** | Provide information about what other people think about the behavior. The information clarifies whether others will like, approve or disapprove of what the person is doing or will do | Tell the staff at the hospital ward that staff at all other wards approve of washing their hands according to the guidelines |

Back to index page

| 7. Associations | | | |
|---|---|---|---|
| **7.1** | ***Prompts/cues*** | Introduce or define environmental or social stimulus with the purpose of prompting or cueing the behavior. The prompt or cue would normally occur at the time or place of performance<br>*Note: when a stimulus is linked to a specific action in an if-then plan including one or more of frequency, duration or intensity also code* ***1.4, Action planning****.* | Put a sticker on the bathroom mirror to remind people to brush their teeth |
| **7.2** | ***Cue signalling reward*** | Identify an environmental stimulus that reliably predicts that reward will follow the behavior (includes ***'Discriminative cue'***) | Advise that a fee will be paid to dentists for a particular dental treatment of 6-8 year old, but not older, children to encourage delivery of that treatment (the 6-8 year old children are the environmental stimulus) |
| **7.3** | ***Reduce prompts/cues*** | Withdraw gradually prompts to perform the behavior (includes ***'Fading'***) | Reduce gradually the number of reminders used to take medication |
| **7.4** | ***Remove access to the reward*** | Advise or arrange for the person to be separated from situations in which unwanted behavior can be rewarded in order to reduce the behavior (includes ***'Time out'***) | Arrange for cupboard containing high calorie snacks to be locked for a specified period to reduce the consumption of sugary foods in between meals |
| **7.5** | ***Remove aversive stimulus*** | Advise or arrange for the removal of an aversive stimulus to facilitate behavior change (includes ***'Escape learning'***) | Arrange for a gym-buddy to stop nagging the person to do more exercise in order to increase the desired exercise behaviour |
| **7.6** | ***Satiation*** | Advise or arrange repeated exposure to a stimulus that reduces or extinguishes a drive for the unwanted behavior | Arrange for the person to eat large quantities of chocolate, in order to reduce the person's appetite for sweet foods |
| **7.7** | ***Exposure*** | Provide systematic confrontation with a feared stimulus to reduce the response to a later encounter | Agree a schedule by which the person who is frightened of surgery will visit the hospital where they are scheduled to have surgery |

Back to index page

| | | | |
|---|---|---|---|
| **7.8** | ***Associative learning*** | Present a neutral stimulus jointly with a stimulus that already elicits the behavior repeatedly until the neutral stimulus elicits that behavior (includes **'Classical/Pavlovian Conditioning'**)<br>*Note: when a BCT involves reward or punishment, code one or more of:* ***10.2, Material reward (behavior); 10.3, Non-specific reward; 10.4, Social reward, 10.9, Self-reward; 10.10, Reward (outcome)*** | Present repeatedly fatty foods with a disliked sauce to discourage the consumption of fatty foods |
| **8. Repetition and substitution** | | | |
| **8.1** | ***Behavioral practice/ rehearsal*** | Prompt practice or rehearsal of the performance of the behavior one or more times in a context or at a time when the performance may not be necessary, in order to increase habit and skill<br>*Note: if aiming to associate performance with the context, also code* ***8.3, Habit formation*** | Prompt asthma patients to practice measuring their peak flow in the nurse's consulting room |
| **8.2** | ***Behavior substitution*** | Prompt substitution of the unwanted behavior with a wanted or neutral behavior<br>*Note: if this occurs regularly, also code* ***8.4, Habit reversal*** | Suggest that the person goes for a walk rather than watches television |
| **8.3** | ***Habit formation*** | Prompt rehearsal and repetition of the behavior in the same context repeatedly so that the context elicits the behavior<br>*Note: also code* ***8.1, Behavioral practice/rehearsal*** | Prompt patients to take their statin tablet before brushing their teeth every evening |
| **8.4** | ***Habit reversal*** | Prompt rehearsal and repetition of an alternative behavior to **replace** an unwanted habitual behavior<br>*Note: also code* ***8.2, Behavior substitution*** | Ask the person to walk up stairs at work where they previously always took the lift |
| **8.5** | ***Overcorrection*** | Ask to repeat the wanted behavior in an exaggerated way following an unwanted behaviour | Ask to eat only fruit and vegetables the day after a poor diet |
| **8.6** | ***Generalisation of a target behavior*** | Advise to perform the wanted behaviour, which is already performed in a particular situation, in another situation | Advise to repeat toning exercises learned in the gym when at home |

Back to index page

| | | | |
|---|---|---|---|
| **8.7** | ***Graded tasks*** | Set easy-to-perform tasks, making them increasingly difficult, but achievable, until behavior is performed | Ask the person to walk for 100 yards a day for the first week, then half a mile a day after they have successfully achieved 100 yards, then two miles a day after they have successfully achieved one mile |
| **9. Comparison of outcomes** | | | |
| **9.1** | ***Credible source*** | Present verbal or visual communication from a **credible source** in favour of or against the behavior<br>*Note: code this BCT if source generally agreed on as credible e.g., health professionals, celebrities or words used to indicate expertise or leader in field and if the communication has the aim of persuading; if information about health consequences, also code* ***5.1, Information about health consequences****, if about emotional consequences, also code* ***5.6, Information about emotional consequences****; if about social, environmental or unspecified consequences also code* ***5.3, Information about social and environmental consequences*** | Present a speech given by a high status professional to emphasise the importance of not exposing patients to unnecessary radiation by ordering x-rays for back pain |
| **9.2** | ***Pros and cons*** | Advise the person to identify and compare reasons for wanting (pros) and not wanting to (cons) change the behavior (includes '**Decisional balance'**)<br>*Note: if providing information about health consequences, also code* ***5.1, Information about health consequences****; if providing information about emotional consequences, also code* ***5.6, Information about emotional consequences****; if providing information about social, environmental or unspecified consequences also code* ***5.3, Information about social and environmental consequences*** | Advise the person to list and compare the advantages and disadvantages of prescribing antibiotics for upper respiratory tract infections |
| **9.3** | ***Comparative imagining of future outcomes*** | Prompt or advise the imagining and comparing of future outcomes of changed versus unchanged behaviour | Prompt the person to imagine and compare likely or possible outcomes following attending versus not attending a screening appointment |
| Back to index page | | | |

| 10. Reward and threat | | | |
|---|---|---|---|
| **10.1** | ***Material incentive (behavior)*** | Inform that money, vouchers or other valued objects ***will be*** delivered if and only if there has been effort and/or progress in performing the behavior (includes **'<u>Positive reinforcement'</u>**)<br>*Note: if incentive is social, code* ***10.5, Social incentive*** *if unspecified code* ***10.6, Non-specific incentive,*** *and <u>not</u>* ***10.1, Material incentive (behavior)****; if incentive is for* ***outcome,*** *code* ***10.8, Incentive (outcome).*** *If reward is delivered also code one of:* ***10.2, Material reward (behavior); 10.3, Non-specific reward; 10.4, Social reward, 10.9, Self-reward; 10.10, Reward (outcome)*** | Inform that a financial payment will be made each month in pregnancy that the woman has not smoked |
| **10.2** | ***Material reward (behavior)*** | Arrange for the delivery of money, vouchers or other valued objects if and only if there ***has been*** effort and/or progress in performing the behavior (includes '**<u>Positive reinforcement'</u>**)<br>*Note: If reward is social, code* ***10.4, Social reward****, if unspecified code* ***10.3, Non-specific reward****, and <u>not</u>* ***10.1, Material reward (behavior)****; if reward is for* ***outcome****, code* ***10.10, Reward (outcome).*** *If informed of reward in advance of rewarded behaviour, also code one of:* ***10.1, Material incentive (behaviour); 10.5, Social incentive; 10.6, Non-specific incentive; 10.7, Self-incentive; 10.8, Incentive (outcome)*** | Arrange for the person to receive money that would have been spent on cigarettes if and only if the smoker has not smoked for one month |
| **10.3** | ***Non-specific reward*** | Arrange delivery of a reward if and only if there ***has been*** effort and/or progress in performing the behavior (includes '**<u>Positive reinforcement'</u>**)<br>*Note: if reward is material, code* ***10.2, Material reward (behavior)****, if social, code* ***10.4, Social reward****, and <u>not</u>* ***10.3, Non-specific reward****; if reward is for* ***outcome*** *code* ***10.10, Reward (outcome).*** *If informed of reward in advance of rewarded behaviour, also code one of:* ***10.1, Material incentive (behaviour); 10.5, Social incentive; 10.6, Non-specific incentive; 10.7, Self-incentive; 10.8, Incentive (outcome)*** | Identify something (e.g. an activity such as a visit to the cinema) that the person values and arrange for this to be delivered if and only if they attend for health screening |

Back to index page

| | | | |
|---|---|---|---|
| **10.4** | ***Social reward*** | Arrange verbal or non-verbal reward if and only if there ***has been*** effort and/or progress in performing the behavior (includes '**Positive reinforcement**') *Note: if reward is material, code* ***10.2, Material reward (behavior)****, if unspecified code* ***10.3, Non-specific reward****, and not* ***10.4, Social reward****; if reward is for* ***outcome*** *code* ***10.10, Reward (outcome).*** *If informed of reward in advance of rewarded behaviour, also code one of:* ***10.1, Material incentive (behaviour); 10.5, Social incentive; 10.6, Non-specific incentive; 10.7, Self-incentive; 10.8, Incentive (outcome)*** | Congratulate the person for each day they eat a reduced fat diet |
| **10.5** | ***Social incentive*** | Inform that a verbal or non-verbal reward ***will be*** delivered if and only if there has been effort and/or progress in performing the behavior (includes '**Positive reinforcement'**) *Note: if incentive is material, code* ***10.1, Material incentive (behavior)****, if unspecified code* ***10.6, Non-specific incentive****, and not* ***10.5, Social incentive****; if incentive is for* ***outcome*** *code* ***10.8, Incentive (outcome).*** *If reward is delivered also code one of:* ***10.2, Material reward (behavior); 10.3, Non-specific reward; 10.4, Social reward, 10.9, Self-reward; 10.10, Reward (outcome)*** | Inform that they will be congratulated for each day they eat a reduced fat diet |
| **10.6** | ***Non-specific incentive*** | Inform that a reward ***will be*** delivered if and only if there has been effort and/or progress in performing the behavior (includes '**Positive reinforcement'**) *Note: if incentive is material, code* ***10.1, Material incentive (behavior)****, if social, code* ***10.5, Social incentive*** *and not* ***10.6, Non-specific incentive****; if incentive is for* ***outcome*** *code* ***10.8, Incentive (outcome).*** *If reward is delivered also code one of:* ***10.2, Material reward (behavior); 10.3, Non-specific reward; 10.4, Social reward, 10.9, Self-reward; 10.10, Reward (outcome)*** | Identify an activity that the person values and inform them that this will happen if and only if they attend for health screening |

Back to index page

| | | | |
|---|---|---|---|
| **10.7** | ***Self-incentive*** | Plan to reward self in future if and only if there has been effort and/or progress in performing the behavior<br>*Note: if self-reward is material, also code **10.1, Material incentive (behavior)**, if social, also code **10.5, Social incentive**, if unspecified, also code **10.6, Non-specific incentive**; if incentive is for **outcome** code **10.8, Incentive (outcome).** If reward is delivered also code one of: **10.2, Material reward (behavior); 10.3, Non-specific reward; 10.4, Social reward, 10.9, Self-reward; 10.10, Reward (outcome)*** | Encourage to provide self with material (e.g., new clothes) or other valued objects if and only if they have adhered to a healthy diet |
| **10.8** | ***Incentive (outcome)*** | Inform that a reward ***will be*** delivered if and only if there has been effort and/or progress in achieving the behavioural **outcome** (*includes* **'Positive reinforcement'**)<br>*Note: this includes social, material, self- and non-specific incentives for outcome; if incentive is for the **behavior** code **10.5, Social incentive**, **10.1, Material incentive (behavior)**, **10.6, Non-specific incentive** or **10.7**, **Self-incentive** and not **10.8, Incentive (outcome).** If reward is delivered also code one of: **10.2, Material reward (behavior); 10.3, Non-specific reward; 10.4, Social reward, 10.9, Self-reward; 10.10, Reward (outcome)*** | Inform the person that they will receive money if and only if a certain amount of weight is lost |
| **10.9** | ***Self-reward*** | Prompt self-praise or self-reward if and only if there ***has been*** effort and/or progress in performing the behavior<br>*Note: if self-reward is material, also code **10.2, Material reward (behavior)**, if social, also code **10.4, Social reward**, if unspecified, also code **10.3, Non-specific reward**; if reward is for **outcome** code **10.10, Reward (outcome).** If informed of reward in advance of rewarded behaviour, also code one of: **10.1, Material incentive (behaviour); 10.5, Social incentive; 10.6, Non-specific incentive; 10.7, Self-incentive; 10.8, Incentive (outcome)*** | Encourage to reward self with material (e.g., new clothes) or other valued objects if and only if they have adhered to a healthy diet |

Back to index page

| | | | |
|---|---|---|---|
| 10.10 | ***Reward (outcome)*** | Arrange for the delivery of a reward if and only if there ***has been*** effort and/or progress in achieving the behavioral **outcome** (includes '**<u>Positive reinforcement</u>**')<br>*Note: this includes social, material, self- and non-specific rewards for outcome; if reward is for the* ***behavior*** *code* ***10.4, Social reward, 10.2, Material reward (behavior), 10.3, Non-specific reward*** *or* ***10.9, Self-reward*** *and <u>not</u>* ***10.10, Reward (outcome).*** *If informed of reward in advance of rewarded behaviour, also code one of****: 10.1, Material incentive (behaviour); 10.5, Social incentive; 10.6, Non-specific incentive; 10.7, Self-incentive; 10.8, Incentive (outcome)*** | Arrange for the person to receive money if and only if a certain amount of weight is lost |
| 10.11 | ***Future punishment*** | Inform that future punishment or removal of reward will be a consequence of performance of an unwanted behavior (may include fear arousal) (includes '**<u>Threat</u>**') | Inform that continuing to consume 30 units of alcohol per day is likely to result in loss of employment if the person continues |
| **11. Regulation** | | | |
| **11.1** | ***Pharmacological support*** | Provide, or encourage the use of or adherence to, drugs to facilitate behavior change<br>*Note: if pharmacological support to reduce negative emotions (i.e. anxiety) then <u>also</u> code* ***11.2, Reduce negative emotions*** | Suggest the patient asks the family physician for nicotine replacement therapy to facilitate smoking cessation |
| **11.2** | ***Reduce negative emotions*** [b] | Advise on ways of reducing negative emotions to facilitate performance of the behavior (includes '**<u>Stress Management</u>**')<br>*Note: if includes analysing the behavioural problem, <u>also</u> code* ***1.2, Problem solving*** | Advise on the use of stress management skills, e.g. to reduce anxiety about joining Alcoholics Anonymous |
| **11.3** | ***Conserving mental resources*** | Advise on ways of minimising demands on mental resources to facilitate behavior change | Advise to carry food calorie content information to reduce the burden on memory in making food choices |
| **11.4** | ***Paradoxical instructions*** | Advise to engage in some form of the unwanted behavior with the aim of reducing motivation to engage in that behaviour | Advise a smoker to smoke twice as many cigarettes a day as they usually do<br><br>Tell the person to stay awake as long as possible in order to reduce insomnia |

Back to index page

| 12. Antecedents | | | |
|---|---|---|---|
| **12.1** | ***Restructuring the physical environment*** | Change, or advise to change the **physical** environment in order to facilitate performance of the wanted behavior or create barriers to the unwanted behavior (other than prompts/cues, rewards and punishments)<br>*Note: this may also involve **12.3, Avoidance/reducing exposure to cues for the behavior**; if restructuring of the social environment code **12.2, Restructuring the social environment;***<br>*if only adding objects to the environment, code **12.5, Adding objects to the environment*** | Advise to keep biscuits and snacks in a cupboard that is inconvenient to get to<br><br>Arrange to move vending machine out of the school |
| **12.2** | ***Restructuring the social environment*** | Change, or advise to change the **social** environment in order to facilitate performance of the wanted behavior or create barriers to the unwanted behavior (other than prompts/cues, rewards and punishments)<br>*Note: this may also involve **12.3, Avoidance/reducing exposure to cues for the behavior**; if also restructuring of the physical environment also code **12.1, Restructuring the physical environment*** | Advise to minimise time spent with friends who drink heavily to reduce alcohol consumption |
| **12.3** | ***Avoidance/reducing exposure to cues for the behavior*** | Advise on how to avoid exposure to specific social and contextual/physical cues for the behavior, including changing daily or weekly routines<br>*Note: this may also involve **12.1, Restructuring the physical environment*** and/or ***12.2, Restructuring the social environment***; if the BCT includes analysing the behavioral problem, <u>only</u> code ***1.2, Problem solving*** | Suggest to a person who wants to quit smoking that their social life focus on activities other than pubs and bars which have been associated with smoking |
| **12.4** | ***Distraction*** | Advise or arrange to use an alternative focus for attention to avoid triggers for unwanted behaviour | Suggest to a person who is trying to avoid between-meal snacking to focus on a topic they enjoy (e.g. holiday plans) instead of focusing on food |

Back to index page

| | | | |
|---|---|---|---|
| **12.5** | ***Adding objects to the environment*** | Add objects to the environment in order to facilitate performance of the behavior *Note: Provision of information (e.g. written, verbal, visual) in a booklet or leaflet is insufficient. If this is accompanied by social support, also code* ***3.2, Social support (practical)****; if the environment is changed beyond the addition of objects, also code* ***12.1, Restructuring the physical environment*** | Provide free condoms to facilitate safe sex<br><br>Provide attractive toothbrush to improve tooth brushing technique |
| **12.6** | ***Body changes*** | Alter body structure, functioning or support **directly** to facilitate behavior change | Prompt strength training, relaxation training or provide assistive aids (e.g. a hearing aid) |
| **13. Identity** | | | |
| **13.1** | ***Identification of self as role model*** | Inform that one's own behavior may be an example to others | Inform the person that if they eat healthily, that may be a good example for their children |
| **13.2** | ***Framing/reframing*** | Suggest the deliberate adoption of a perspective or new perspective on behavior (e.g. its purpose) in order to change cognitions or emotions about performing the behavior (includes '**Cognitive structuring**'); *If information about consequences then code* ***5.1, Information about health consequences, 5.6, Information about emotional consequences*** *or* ***5.3, Information about social and environmental consequences*** *instead of* ***13.2, Framing/reframing*** | Suggest that the person might think of the tasks as reducing sedentary behavior (rather than increasing activity) |
| **13.3** | ***Incompatible beliefs*** | Draw attention to discrepancies between current or past behavior and self-image, in order to create discomfort (includes **'Cognitive dissonance'**) | Draw attention to a doctor's liberal use of blood transfusion and their self-identification as a proponent of evidence-based medical practice |
| **13.4** | ***Valued self-identity*** | Advise the person to write or complete rating scales about a cherished value or personal strength as a means of affirming the person's identity as part of a behavior change strategy (includes **'Self-affirmation'**) | Advise the person to write about their personal strengths before they receive a message advocating the behavior change |
| **13.5** | ***Identity associated with changed behavior*** | Advise the person to construct a new self-identity as someone who 'used to engage with the unwanted behavior' | Ask the person to articulate their new identity as an 'ex-smoker' |

Back to index page

| 14. Scheduled consequences | | | |
|---|---|---|---|
| **14.1** | ***Behavior cost*** | Arrange for withdrawal of something valued if and only if an unwanted behavior is performed (includes '**Response cost'**). Note if withdrawal of contingent reward code, ***14.3, Remove reward*** | Subtract money from a prepaid refundable deposit when a cigarette is smoked |
| **14.2** | ***Punishment*** | Arrange for aversive consequence contingent on the performance of the unwanted behavior | Arrange for the person to wear unattractive clothes following consumption of fatty foods |
| **14.3** | ***Remove reward*** | Arrange for discontinuation of contingent reward following performance of the unwanted behavior (includes **'Extinction'**) | Arrange for the other people in the household to ignore the person every time they eat chocolate (rather than attending to them by criticising or persuading) |
| **14.4** | ***Reward approximation*** | Arrange for reward following any approximation to the target behavior, gradually rewarding only performance closer to the wanted behavior (includes **'Shaping'**)<br>*Note: also code one of* ***59-63*** | Arrange reward for any reduction in daily calories, gradually requiring the daily calorie count to become closer to the planned calorie intake |
| **14.5** | ***Rewarding completion*** | Build up behavior by arranging reward following final component of the behavior; gradually add the components of the behavior that occur earlier in the behavioral sequence (includes **'Backward chaining'**)<br>*Note: also code one of* ***10.2, Material reward (behavior); 10.3, Non-specific reward; 10.4, Social reward, 10.9, Self-reward; 10.10, Reward (outcome)*** | Reward eating a supplied low calorie meal; then make reward contingent on cooking and eating the meal; then make reward contingent on purchasing, cooking and eating the meal |
| **14.6** | ***Situation-specific reward*** | Arrange for reward following the behavior in one situation but not in another (includes **'Discrimination training'**)<br>*Note: also code one of* ***10.2, Material reward (behavior); 10.3, Non-specific reward; 10.4, Social reward, 10.9, Self-reward; 10.10, Reward (outcome)*** | Arrange reward for eating at mealtimes but not between meals |
| **14.7** | ***Reward incompatible behavior*** | Arrange reward for responding in a manner that is incompatible with a previous response to that situation (includes **'Counter-conditioning'**)<br>*Note: also code one of* ***10.2, Material reward (behavior); 10.3, Non-specific reward; 10.4, Social reward, 10.9, Self-reward; 10.10, Reward (outcome)*** | Arrange reward for ordering a soft drink at the bar rather than an alcoholic beverage |

Back to index page

| | | | |
|---|---|---|---|
| 14.8 | ***Reward alternative behavior*** | Arrange reward for performance of an alternative to the unwanted behavior (includes ***'*Differential reinforcement*'***)<br>*Note: also code one of* ***10.2, Material reward (behavior); 10.3, Non-specific reward; 10.4, Social reward, 10.9, Self-reward; 10.10, Reward (outcome);*** *consider also coding* ***1.2, Problem solving*** | Reward for consumption of low fat foods but not consumption of high fat foods |
| 14.9 | ***Reduce reward frequency*** | Arrange for rewards to be made contingent on increasing duration or frequency of the behavior (includes ***'*Thinning*'***)<br>*Note: also code one of* ***10.2, Material reward (behavior); 10.3, Non-specific reward; 10.4, Social reward, 10.9, Self-reward; 10.10, Reward (outcome)*** | Arrange reward for each day without smoking, then each week, then each month, then every 2 months and so on |
| 14.10 | ***Remove punishment*** | Arrange for removal of an unpleasant consequence contingent on performance of the wanted behavior (includes **'Negative reinforcement'**) | Arrange for someone else to do housecleaning only if the person has adhered to the medication regimen for a week |
| **15. Self-belief** | | | |
| 15.1 | ***Verbal persuasion about capability*** | Tell the person that they can successfully perform the wanted behavior, arguing against self-doubts and asserting that they can and will succeed | Tell the person that they can successfully increase their physical activity, despite their recent heart attack. |
| 15.2 | ***Mental rehearsal of successful performance*** | Advise to practise imagining performing the behavior successfully in relevant contexts | Advise to imagine eating and enjoying a salad in a work canteen |
| 15.3 | ***Focus on past success*** | Advise to think about or list previous successes in performing the behavior (or parts of it) | Advise to describe or list the occasions on which the person had ordered a non-alcoholic drink in a bar |
| 15.4 | ***Self-talk*** | Prompt positive self-talk (aloud or silently) before and during the behavior | Prompt the person to tell themselves that a walk will be energising |
| **16. Covert learning** | | | |
| 16.1 | ***Imaginary punishment*** | Advise to imagine performing the **unwanted** behavior in a real-life situation followed by imagining an unpleasant consequence (includes **'Covert sensitisation'**) | Advise to imagine overeating and then vomiting |

Back to index page

| | | | |
|---|---|---|---|
| **16.2** | ***Imaginary reward*** | Advise to imagine performing the **wanted** behavior in a real-life situation followed by imagining a pleasant consequence (includes ***'Covert conditioning'***) | Advise the health professional to imagine giving dietary advice followed by the patient losing weight and no longer being diabetic |
| **16.3** | ***Vicarious consequences*** | Prompt observation of the consequences (including rewards and punishments) for others when they perform the behavior *Note: if observation of health consequences, also code* ***5.1, Information about health consequences****; if of emotional consequences, also code* ***5.6, Information about emotional consequences****, if of social, environmental or unspecified consequences, also code* ***5.3, Information about social and environmental consequences*** | Draw attention to the positive comments other staff get when they disinfect their hands regularly |
| Back to index page | | | |

[a] Notes are provided underneath most BCTs to help distinguish them from similar techniques

[b] An additional technique 'Increase positive emotions' will be included in BCT Taxonomy v2
