## AppendixD_PA-QC-RoB for "Intervention Characteristics and Mechanisms and their Relationship with the Influence of Social Prescribing: a Systematic Review"

**Appendix D:** Percent agreement, quality assessment, and Risk of Bias.

**Percent agreement**

| **Table AC.1.** Percent agreement quality check between EMD, FHA, MB, SGLS, and LAV | | | | | | | |
| --- | --- | --- | --- | --- | --- | --- | --- |
| **Qualitative studies** (*n*=3) | | **Randomied controlled trials** (*n*=1) | | **Non-randomized studies** (*n*=4) | | **Mixed methods studies** (*n*=1) | |
| Item | Agreement | Item | Agreement | Item | Agreement | Item | Agreement |
| 1.1 | 100.0 | 2.1 | 60.0 | 3.1 | 60.0 | 5.1 | 80.0 |
| 1.2 | 93.3 | 2.2 | 100.0 | 3.2 | 90.0 | 5.2 | 60.0 |
| 1.3 | 80.0 | 2.3 | 80.0 | 3.3 | 70.0 | 5.3 | 60.0 |
| 1.4 | 93.3 | 2.4 | 60.0 | 3.4 | 85.0 | 5.4 | 60.0 |
| 1.5 | 93.3 | 2.5 | 100.0 | 3.5 | 53.3 | 5.5 | 80.0 |
| Mean | 92.0 | Mean | 80.0 | Mean | 71.7 | Mean | 68.0 |

| **Table AC.2.** Percent agreement study and intervention characteristics and outcomes between EMD, MB, SGLS, and LAV | | | | | |
| --- | --- | --- | --- | --- | --- |
|  | **Qualitative studies** (*n*=2) | **Randomied controlled trials** (*n*=1) | **Non-randomized studies** (*n*=3) | **Mixed methods studies** (*n*=1) | **All studies** |
| Study characteritics | 96.9 | 100.0 | 94.8 | 81.2 | 94.2 |
| Intervention characteristics | 90.5 | 87.4 | 89.1 | 88.8 | 89.2 |
| Outcomes | 56.3 | 66.7 | 70.6 | 71.4 | 66.0 |
| Mean | 81.2 | 84.7 | 84.8 | 80.5 | 83.1 |

| **Table AC.3.** Quality assessment for all included studies* | | | | | | | | | | | | | | | | | | | | | | | | | | | | | | |
| --- | --- | --- | --- | --- | --- | --- | --- | --- | --- | --- | --- | --- | --- | --- | --- | --- | --- | --- | --- | --- | --- | --- | --- | --- | --- | --- | --- | --- | --- | --- |
|  | Qualitative studies | | | | | Randomized controlled trials | | | | | Non-randomized studies | | | | | | Descriptive studies | | | | | | Mixed methods studies | | | | | | **Quality**  **Score** | |
|  | 1.1 | 1.2 | 1.3 | 1.4 | 1.5 | 2.1 | 2.2 | 2.3 | 2.4 | 2.5 | | 3.1 | 3.2 | 3.3 | 3.4 | 3.5 | | 4.1 | 4.2 | 4.3 | 4.4 | 4.5 | | 5.1 | 5.2 | 5.3 | 5.4 | 5.5 | |  |
| Aggar et al., 2021 |  |  |  |  |  |  |  |  |  |  | | 0 | 1 | 0 | 0 | 1 | |  |  |  |  |  | |  |  |  |  |  | | Low |
| Bertotti et al., 2017 | 1 | 0 | 0 | 1 | 0 |  |  |  |  |  | | 0 | 1 | 0 | 0 | 0 | |  |  |  |  |  | | 1 | 1 | 1 | 1 | 0 | | Low |
| Bertotti et al., 2018 | 1 | 0 | 0 | 1 | 0 |  |  |  |  |  | |  |  |  |  |  | | 1 | 1 | 0 | 1 | 0 | | 1 | 1 | 1 | 0 | 0 | | Low |
| Bertotti et al., 2020 | 1 | 0 | 0 | 1 | 0 |  |  |  |  |  | | 0 | 1 | 0 | 0 | 0 | |  |  |  |  |  | | 0 | 0 | 0 | 0 | 0 | | Low |
| Bhatti et al., 2021 | 1 | 1 | 1 | 1 | 1 |  |  |  |  |  | |  |  |  |  |  | |  |  |  |  |  | |  |  |  |  |  | | High |
| Brettell et al., 2022 |  |  |  |  |  |  |  |  |  |  | | 0 | 1 | 0 | 0 | 0 | |  |  |  |  |  | |  |  |  |  |  | | Low |
| Carnes et al., 2017 | 1 | 1 | 1 | 1 | 1 |  |  |  |  |  | | 0 | 0 | 1 | 1 | 0 | |  |  |  |  |  | | 1 | 0 | 0 | 1 | 0 | | Low |
| Crone et al., 2012 | 1 | 1 | 1 | 1 | 1 |  |  |  |  |  | |  |  |  |  |  | |  |  |  |  |  | |  |  |  |  |  | | High |
| Crone et al., 2018 |  |  |  |  |  |  |  |  |  |  | | 1 | 1 | 1 | 1 | 0 | |  |  |  |  |  | |  |  |  |  |  | | High |
| Dayson et al., 2015 | 0 | 0 | 0 | 1 | 0 |  |  |  |  |  | | 0 | 0 | 0 | 0 | 0 | |  |  |  |  |  | | 0 | 0 | 0 | 0 | 0 | | Low |
| Dayson et al., 2016 | 0 | 0 | 0 | 1 | 0 |  |  |  |  |  | | 0 | 1 | 0 | 0 | 0 | | 0 | 0 | 0 | 0 | 0 | | 0 | 0 | 0 | 0 | 0 | | Low |
| Dayson et al., 2017 | 1 | 0 | 0 | 1 | 0 |  |  |  |  |  | | 0 | 0 | 0 | 0 | 0 | |  |  |  |  |  | | 1 | 1 | 1 | 1 | 0 | | Low |
| Envoy Partnership 2018 | 1 | 0 | 0 | 1 | 0 |  |  |  |  |  | | 0 | 1 | 0 | 0 | 0 | |  |  |  |  |  | | 1 | 1 | 1 | 1 | 0 | | Low |
| Farenden et al., 2015 |  |  |  |  |  |  |  |  |  |  | |  |  |  |  |  | | 1 | 1 | 0 | 1 | 0 | |  |  |  |  |  | | Moderate |
| Foster et al., 2021 | 1 | 1 | 1 | 1 | 1 |  |  |  |  |  | | 1 | 1 | 1 | 0 | 1 | |  |  |  |  |  | | 1 | 1 | 1 | 1 | 1 | | High |
| Friedli et al., 2012 | 0 | 0 | 0 | 1 | 0 |  |  |  |  |  | | 0 | 0 | 0 | 0 | 0 | |  |  |  |  |  | | 0 | 0 | 0 | 0 | 0 | | Low |
| Hanlon et al., 2021 | 1 | 1 | 1 | 1 | 1 |  |  |  |  |  | |  |  |  |  |  | |  |  |  |  |  | |  |  |  |  |  | | High |
| Heijnders et al., 2018 | 1 | 1 | 1 | 1 | 1 |  |  |  |  |  | |  |  |  |  |  | |  |  |  |  |  | |  |  |  |  |  | | High |
| Holt et al., 2020 |  |  |  |  |  |  |  |  |  |  | | 1 | 1 | 1 | 0 | 1 | |  |  |  |  |  | |  |  |  |  |  | | High |
| Howarth et al., 2019 | 0 | 0 | 0 | 1 | 0 |  |  |  |  |  | |  |  |  |  |  | | 0 | 1 | 1 | 0 | 0 | | 0 | 0 | 0 | 0 | 0 | | Low |
| Hughes et al., 2019 | 1 | 1 | 1 | 1 | 1 |  |  |  |  |  | | 1 | 1 | 0 | 1 | 0 | |  |  |  |  |  | | 1 | 1 | 1 | 1 | 0 | | Moderate |
| Jones and Lynch 2019 | 0 | 0 | 0 | 1 | 0 |  |  |  |  |  | | 0 | 1 | 0 | 0 | 0 | |  |  |  |  |  | | 0 | 0 | 0 | 0 | 0 | | Low |
| Jones and Lynch 2020 | 0 | 0 | 0 | 1 | 0 |  |  |  |  |  | | 0 | 1 | 0 | 0 | 0 | |  |  |  |  |  | | 0 | 0 | 0 | 0 | 0 | | Low |
| Kellezi et al., 2019 | 1 | 1 | 1 | 1 | 1 |  |  |  |  |  | | 1 | 1 | 0 | 1 | 1 | |  |  |  |  |  | | 1 | 1 | 1 | 1 | 1 | | High |
| Kim et al., 2021 |  |  |  |  |  |  |  |  |  |  | | 0 | 1 | 0 | 0 | 0 | |  |  |  |  |  | |  |  |  |  |  | | Low |
| Kimberlee et al., 2014 | 0 | 0 | 0 | 1 | 0 |  |  |  |  |  | | 1 | 1 | 0 | 0 | 0 | |  |  |  |  |  | | 1 | 1 | 1 | 1 | 0 | | Low |
| Kimberlee et al., 2016 | 0 | 0 | 0 | 1 | 0 |  |  |  |  |  | | 1 | 1 | 0 | 0 | 0 | |  |  |  |  |  | | 0 | 0 | 0 | 0 | 0 | | Low |
| Loftus et al., 2017 |  |  |  |  |  |  |  |  |  |  | | 0 | 1 | 1 | 0 | 0 | |  |  |  |  |  | |  |  |  |  |  | | Low |
| Lynch and Jones, 2022 |  |  |  |  |  |  |  |  |  |  | | 1 | 1 | 1 | 0 | 0 | |  |  |  |  |  | |  |  |  |  |  | | Moderate |
| Maughan et al., 2016 |  |  |  |  |  |  |  |  |  |  | | 1 | 1 | 0 | 0 | 0 | |  |  |  |  |  | |  |  |  |  |  | | Low |
| Mercer et al., 2019 |  |  |  |  |  | 0 | 0 | 0 | 0 | 1 | |  |  |  |  |  | |  |  |  |  |  | |  |  |  |  |  | | Low |
| Morton et al., 2015 |  |  |  |  |  |  |  |  |  |  | | 1 | 1 | 0 | 1 | 1 | |  |  |  |  |  | |  |  |  |  |  | | High |
| Mulligan et al., 2020 | 1 | 0 | 1 | 1 | 0 |  |  |  |  |  | | 1 | 0 | 0 | 0 | 1 | |  |  |  |  |  | | 1 | 1 | 1 | 1 | 0 | | Low |
| Palmer et al., 2017 | 1 | 1 | 1 | 1 | 0 |  |  |  |  |  | | 1 | 1 | 1 | 0 | 0 | |  |  |  |  |  | | 0 | 0 | 1 | 1 | 0 | | Low |
| Payne et al., 2020 | 1 | 1 | 1 | 1 | 1 |  |  |  |  |  | |  |  |  |  |  | |  |  |  |  |  | |  |  |  |  |  | | High |
| Pescheny et al., 2019 |  |  |  |  |  |  |  |  |  |  | | 1 | 1 | 0 | 0 | 0 | |  |  |  |  |  | |  |  |  |  |  | | Low |
| Pescheny et al., 2021 |  |  |  |  |  |  |  |  |  |  | | 0 | 1 | 1 | 1 | 0 | |  |  |  |  |  | |  |  |  |  |  | | Moderate |
| Polley et al., 2019 | 1 | 1 | 1 | 1 | 1 |  |  |  |  |  | | 1 | 1 | 0 | 0 | 0 | |  |  |  |  |  | | 1 | 1 | 1 | 1 | 0 | | Low |
| Pomp, 2015 |  |  |  |  |  |  |  |  |  |  | | 1 | 0 | 1 | 1 | 0 | |  |  |  |  |  | |  |  |  |  |  | | Moderate |
| Potter et al., 2015 | 1 | 1 | 1 | 1 | 1 |  |  |  |  |  | | 0 | 1 | 0 | 0 | 0 | |  |  |  |  |  | | 1 | 1 | 1 | 1 | 0 | | Low |
| Poulos et al., 2019 | 1 | 1 | 1 | 1 | 1 |  |  |  |  |  | | 0 | 1 | 1 | 0 | 0 | |  |  |  |  |  | | 0 | 0 | 1 | 1 | 0 | | Low |
| Redmond et al., 2019 | 1 | 1 | 1 | 1 | 1 |  |  |  |  |  | |  |  |  |  |  | |  |  |  |  |  | |  |  |  |  |  | | High |
| Simpson et al., 2020 | 1 | 1 | 1 | 1 | 1 |  |  |  |  |  | |  |  |  |  |  | |  |  |  |  |  | |  |  |  |  |  | | High |
| Sumner et al., 2020 |  |  |  |  |  |  |  |  |  |  | | 1 | 1 | 1 | 1 | 0 | |  |  |  |  |  | |  |  |  |  |  | | High |
| Sumner et al., 2021 |  |  |  |  |  |  |  |  |  |  | | 1 | 1 | 1 | 1 | 1 | |  |  |  |  |  | |  |  |  |  |  | | High |
| van de Venter et al., 2015 | 1 | 1 | 1 | 1 | 1 |  |  |  |  |  | | 1 | 1 | 1 | 0 | 0 | |  |  |  |  |  | | 1 | 1 | 1 | 1 | 0 | | Moderate |
| Vogelpoel et al., 2014 | 1 | 1 | 0 | 1 | 1 |  |  |  |  |  | | 1 | 1 | 0 | 0 | 0 | |  |  |  |  |  | | 1 | 1 | 1 | 1 | 0 | | Low |
| Wakefield et al., 2020 |  |  |  |  |  |  |  |  |  |  | | 0 | 1 | 0 | 1 | 0 | |  |  |  |  |  | |  |  |  |  |  | | Low |
| Woodall et al., 2018 | 1 | 1 | 1 | 1 | 1 |  |  |  |  |  | | 0 | 1 | 0 | 1 | 0 | |  |  |  |  |  | | 1 | 1 | 1 | 1 | 0 | | Low |
| * 0=no/can’t tell; 1=yes. | | | | | | | | | | | | | | | | | | | | | | | | | | | | | | |

**Quality assessment using the Mixed Methods Appraisal Tool (MMAT).**

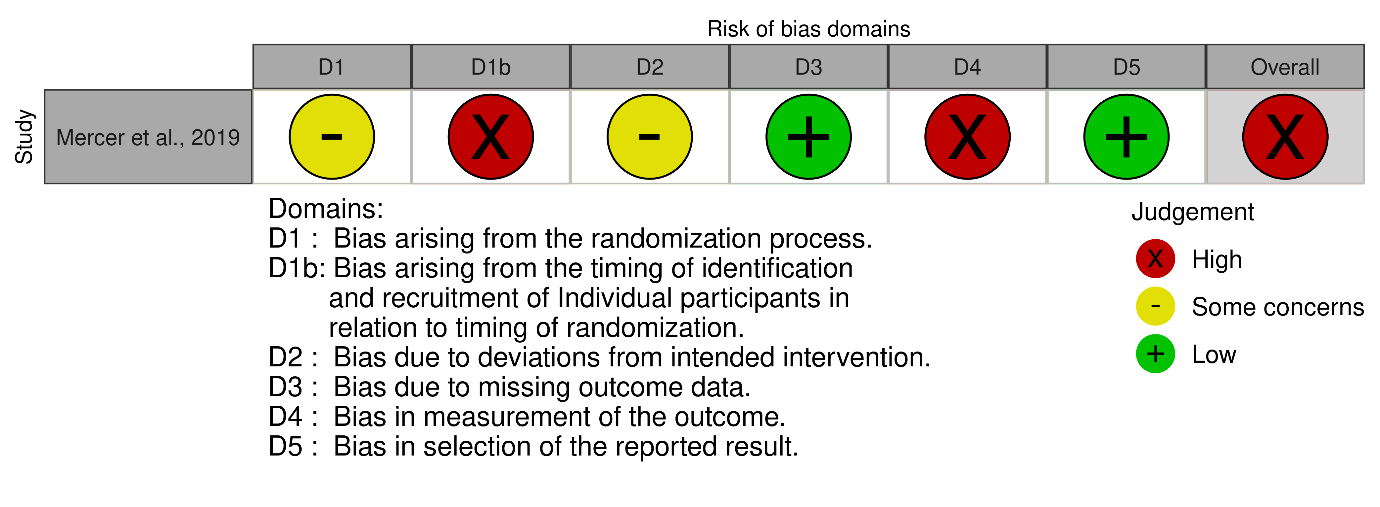
  **Risk of bias**

**Figure AC.2.** Risk of bias table for Carnes et al., 2017 and Pomp., 2015

**Figure AC.1.** Risk of bias table for Mercer et al., 2019

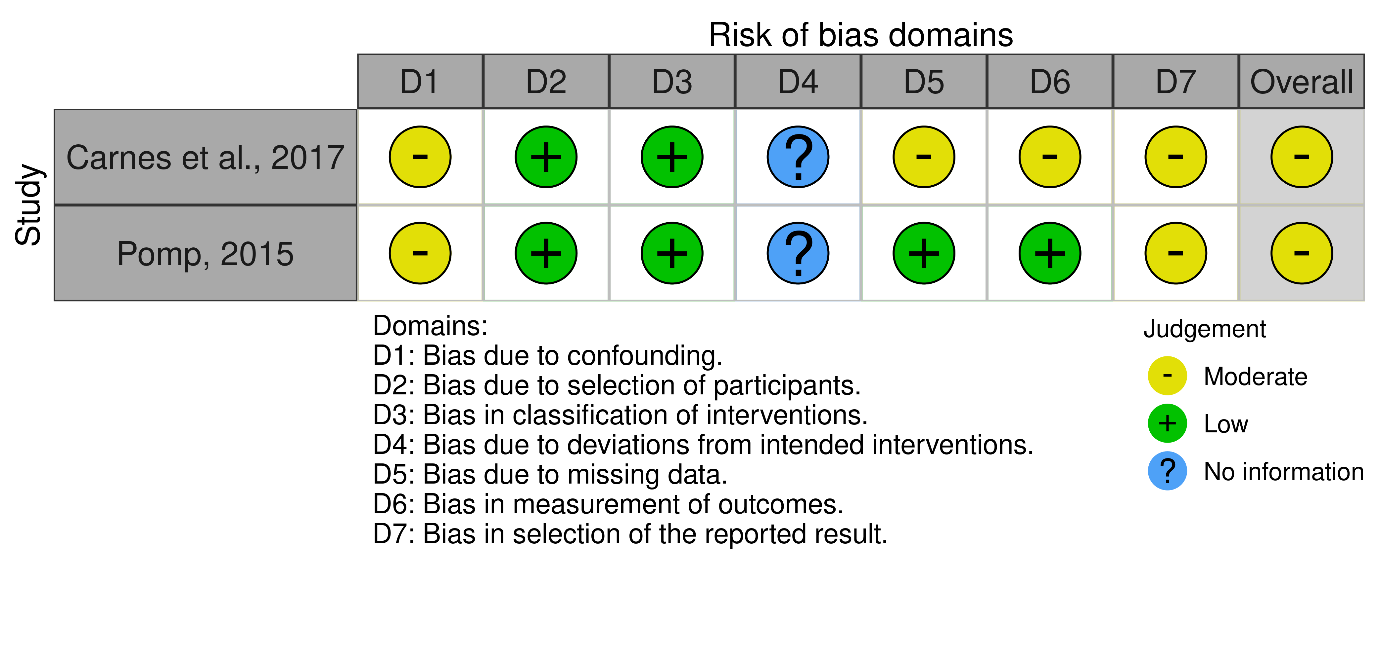
