## AppendixE_ExcludedReferences for "Intervention Characteristics and Mechanisms and their Relationship with the Influence of Social Prescribing: a Systematic Review"

**Appendix E.** Excluded references including reasons for exclusion

| **(first) Author** | **Year** | **Title** | **Reason for exclusion** |
| --- | --- | --- | --- |
| **INITIAL SEARCH** | | | |
| Anonymous | 2019 | Erratum: Correction: Personal Wellbeing Score (PWS)-a short version of ONS4: development and validation in social prescribing | Other |
| Anonymous | 2019 | Erratum: Correction: Personal Wellbeing Score (PWS)-a short version of ONS4: development and validation in social prescribing | Other |
| Augtherson | 2020 | Social prescribing for individuals with mental health problems: a qualitative study of barriers and enablers experienced by general practitioners | Not focused on the influence of SP (on quadruple aim outcomes) |
| Baker | 2016 | Co-producing Approaches to the Management of Dementia through Social Prescribing | Not focused on the influence of SP (on quadruple aim outcomes) |
| Baker | 2012 | Eco Arts on Prescription | Mental health recovery program / exercise referral scheme |
| Baska | 2021 | Social Prescribing and Lifestyle Medicine-A Remedy to Chronic Health Problems? | Study type not suited |
| BattRawden | 2020 | Singing has empowered, enchanted and enthralled me'-choirs for wellbeing? | Patients not referred from primary care |
| Baxter | 2020 | What are the barriers to, and enablers of, working with people with lived experience of mental illness amongst community and voluntary sector organisations? A qualitative study | Not focused on the influence of SP (on quadruple aim outcomes) |
| Beardmore | 2019 | Working in social prescribing services: a qualitative study | Not focused on the influence of SP (on quadruple aim outcomes) |
| Benson | 2019 | Personal Wellbeing Score (PWS)-a short version of ONS4: development and validation in social prescribing | Not focused on the influence of SP (on quadruple aim outcomes) |
| Benson | 2019 | Personal Wellbeing Score (PWS)-a short version of ONS4: development and validation in social prescribing | Not focused on the influence of SP (on quadruple aim outcomes) |
| Bird | 2020 | Reducing chronic stress to promote health in adults: the role of social prescriptions and social movements | Study type not suited |
| Bird | 2019 | General practice referral of ‘at risk’ populations to community leisure services: applying the RE-AIM framework to evaluate the impact of a community-based physical activity programme for inactive adults with long-term conditions | Mental health recovery program / exercise referral scheme |
| Blickem | 2014 | Aligning everyday life priorities with people’s self-management support networks: an exploration of the work and implementation of a needs-led telephone support system | Mental health recovery program / exercise referral scheme |
| Brandborg | 2021 | Physical activity through social prescribing: An interview-based study of Danish general practitioners' opinions | Not focused on the influence of SP (on quadruple aim outcomes) |
| Brandling | 2009 | Social prescribing in general practice: adding meaning to medicine | Study type not suited |
| Burns | 2021 | Happy Hookers: findings from an international study exploring the effects of crochet on wellbeing | Patients not referred from primary care |
| Camic | 2021 | Subjective wellbeing in people living with dementia: exploring processes of multiple object handling sessions in a museum setting | Patients not referred from primary care |
| Carty | 2016 | Books on Prescription – community-based health initiative to increase access to mental health treatment: an evaluation | Mental health recovery program / exercise referral scheme |
| Chatterjee | 2018 | Social prescribing: community-based referral in public health | Study type not suited |
| Chesterman | 2018 | Report on some action research in the implementation of social prescription in Crawley. Paths to greater wellbeing: ‘sometimes you have to be in it to get it’ | Referral reasons not suited |
| Chitson | 2020 | The impact and potential value for medical students of implementing social prescribing projects in primary care | Not focused on the influence of SP (on quadruple aim outcomes) |
| Chiva Giurca | 2018 | Social prescribing student champion scheme: a novel peer-assisted-learning approach to teaching social prescribing and social determinants of health | Not focused on the influence of SP (on quadruple aim outcomes) |
| Chng | 2021 | Implementing social prescribing in primary care in areas of high socioeconomic deprivation: process evaluation of the 'Deep End' community Links Worker Programme | Not focused on the influence of SP (on quadruple aim outcomes) |
| Costa | 2021 | Developing a social prescribing local system in a European Mediterranean country: a feasibility study to promote active and healthy aging | Not focused on the influence of SP (on quadruple aim outcomes) |
| Crabtree | 2018 | Men's sheds: The perceived health and wellbeing benefits | Patients not referred from primary care |
| Dauenhauer | 2006 | Prescribing exercise for older adults: A needs assessment comparing primary care physicians, nurse practitioners, and physician assistants | Mental health recovery program / exercise referral scheme |
| Davies | 2014 | The art of being healthy: a qualitative study to develop a thematic framework for understanding the relationship between health and the arts | Patients not referred from primary care |
| Dayson | 2020 | A Comparative Analysis of Social Impact Bond and Conventional Financing Approaches to Health Service Commissioning in England: The Case of Social Prescribing | Not focused on the influence of SP (on quadruple aim outcomes) |
| De Rada | 2014 | Community resources prescription for self-care improvement in chronic illnesses. Clinical case management in Primary Health Care | Study type not suited |
| Dowden | 2019 | How social prescribing can benefit patients and prescriber | Study type not suited |
| Drennan | 2018 | Evaluating the use of social prescribing coordinators in general practices | Other |
| Duffin | 2016 | Assessing the benefits of social prescribing | Other |
| Duong | 2015 | Perception of Exercise Lifestyle as a Valid Tool for Prevention and Treatment of Depression in Rural Communities | Mental health recovery program / exercise referral scheme |
| Edwards | 2021 | An exploration of engagement in community based creative activities as an occupation for older adults | Patients not referred from primary care |
| Elston | 2019 | Does a social prescribing ‘holistic’ link-worker for older people with complex, multimorbidity improve well-being and frailty and reduce  health and social care use and costs? A 12-month before-and-after evaluation | Referral reasons not suited |
| Escobar-Roldan | 2021 | Exercise Prescription Practices to Improve Mental Health | Mental health recovery program / exercise referral scheme |
| Esmene | 2020 | Beyond adherence to social prescriptions: How places, social acquaintances and stories help walking group members to thrive | Mental health recovery program / exercise referral scheme |
| Estevao | 2021 | Scaling-up Health-Arts Programmes: the largest study in the world bringing arts-based mental health interventions into a national health service | Study type not suited |
| Fancourt | 2020 | [Letter to the editor] Fixed-Effects Analyses of Time-Varying Associations between Hobbies and Depression in a Longitudinal Cohort Study: Support for Social Prescribing? | Other |
| Fancourt | 2020 | Community engagement and dementia risk: time-to-event analyses from a national cohort study | Patients not referred from primary care |
| Ferez | 2014 | Continuing to engage in recreation after being infected with HIV: between quest for normality and social prescription | Not written in Dutch or English |
| Fixsen | 2020 | Applying critical systems thinking to social prescribing: a relational model of stakeholder "buy-in" | Not focused on the influence of SP (on quadruple aim outcomes) |
| Fixsen | 2021 | Weathering the storm: A qualitative study of social prescribing in urban and rural Scotland during the COVID-19 pandemic | Not focused on the influence of SP (on quadruple aim outcomes) |
| Fleming | 2020 | Collaboration between primary care and a voluntary, community sector organisation: Practical guidance from the parkrun practice initiative | Not focused on the influence of SP (on quadruple aim outcomes) |
| Fleming | 2020 | Engagement with and delivery of the 'parkrun practice initiative' in general practice: a mixed methods study | Not focused on the influence of SP (on quadruple aim outcomes) |
| Frostick | 2021 | The frontline of social prescribing - How do we ensure Link Workers can work safely and effectively within primary care? | Not focused on the influence of SP (on quadruple aim outcomes) |
| Fuller | 2017 | Patient empowerment: a prescription for success | Study type not suited |
| Gallacher | 2021 | Social prescribing for dementia | Study type not suited |
| Galway | 2019 | Adapting Digital Social Prescribing for Suicide Bereavement Support: The Findings of a Consultation Exercise to Explore the Acceptability of Implementing Digital Social Prescribing within an Existing Postvention Service | Not focused on the influence of SP (on quadruple aim outcomes) |
| Garner-Purkis | 2020 | A community-based, sport-led programme to increase physical activity in an area of deprivation: a qualitative case study | Mental health recovery program / exercise referral scheme |
| Gibson | 2021 | Social prescribing and classed inequality: A journey of upward health mobility? | Referral reasons not suited |
| Giebel | 2021 | A socially prescribed community service for people living with dementia and family carers and its long-term effects on well-being | Referral reasons not suited |
| Golubinsky | 2020 | Once is rarely enough: can social prescribing facilitate adherence to nonclinical community and voluntary sector health services? Empirical evidence from Germany | Not focused on the influence of SP (on quadruple aim outcomes) |
| Gomez-Juanes | 2015 | Exercise Prescription for Depression by General Practitioners, Factors Involved | Not written in Dutch or English |
| Goodrich | 2011 | Integrating an internet-mediated walking program into family medicine clinical practice: a pilot feasibility study | Mental health recovery program / exercise referral scheme |
| Gradinger | 2019 | [poster abstract] Implementation and impact of co-locating the voluntary sector with a multidisciplinary, cross-sector community hub at the Integrated Care Organisation (ICO) in Torbay and South Devon, UK | Other |
| Gupta | 1996 | A two-year review of an 'open access' multidisciplinary community psychiatric service for the elderly | Patients not referred from primary care |
| Hassan | 2020 | Social prescribing for people with mental health needs living in disadvantaged communities: the Life Rooms model | Mental health recovery program / exercise referral scheme |
| Hazeldine | 2021 | Link worker perspectives of early implementation of social prescribing: A 'Researcher-in-Residence' study | Patients not referred from primary care |
| Hildson | 1998 | Promoting physical activity: issues in primary health care | Before 2010 |
| Hodge | 2007 | Reading between the lines: the experiences of taking part in a community reading project | Patients not referred from primary care |
| Hodgson | 2021 | Integrating primary care and social services for older adults with multimorbidity: policy implications | Study type not suited |
| Holding | 2020 | Connecting communities: A qualitative investigation of the challenges in delivering a national social prescribing service to reduce loneliness | Not focused on the influence of SP (on quadruple aim outcomes) |
| Howarth | 2018 | Growing spaces: an evaluation of the mental health recovery programme using mixed methods | Mental health recovery program / exercise referral scheme |
| Howarth | 2021 | Creating a transformative space for change: A qualitative evaluation of the RHS Wellbeing Programme for people with long term conditions | Mental health recovery program / exercise referral scheme |
| Howarth | 2021 | Personalised solutions through social prescribing | Study type not suited |
| Husk | 2018 | Prescribing gardening and conservation activities for health and wellbeing in older people | Study type not suited |
| Ivory | 2019 | Perspectives: A refreshing and welcome boost for the arts through social prescribing | Other |
| Jani | 2020 | Use and impact of social prescribing: a mixed-methods feasibility study protocol | Other |
| Jani | 2020 | Investing resources to address social factors affecting health: the essential role of social prescribing | Study type not suited |
| Jaswal | 2018 | [poster abstract] Social Prescribing and Integrated Care: An Evaluability Assessment | Other |
| Jensen | 2018 | The use of arts interventions for mental health and wellbeing in health settings | Study type not suited |
| Jensen | 2020 | An Arts on Prescription programme: Perspectives of the cultural institutions | Not focused on the influence of SP (on quadruple aim outcomes) |
| Jensen | 2019 | Aesthetic engagement as health and wellbeing promotion | Patients not referred from primary care |
| Jensen | 2019 | Culture Vitamins – an Arts on Prescription project in Denmark | Patients not referred from primary care |
| Johansson | 2021 | Let’s Try Social Prescribing in Sweden (SPiS) – an Interventional Project Targeting Loneliness among Older Adults Using a Model for Integrated Care: A Research Protocol | Other |
| Johansson | 2021 | Can an ecological-transactional systems model in occupational therapy contribute to a social prescribing programme? | Not focused on the influence of SP (on quadruple aim outcomes) |
| Jones | 2020 | Social Return on Investment Analysis of the Health Precinct Community Hub for Chronic Conditions | Mental health recovery program / exercise referral scheme |
| Joosten | 2007 | Preferences for Accepting Prescribed Community-Based, Psychosocial, and In-Home Services by Older Adults | Not focused on the influence of SP (on quadruple aim outcomes) |
| Khan | 2021 | Public perspectives of social prescribing | Not focused on the influence of SP (on quadruple aim outcomes) |
| Kiely | 2021 | Primary care-based link workers providing social prescribing to improve health and social care outcomes for people with multimorbidity in socially deprived areas (the LinkMM trial): Pilot study for a pragmatic randomised controlled trial | Not focused on the influence of SP (on quadruple aim outcomes) |
| Kime | 2012 | The delivery and management of telephone befriending services – whose needs are being met? | Patients not referred from primary care |
| Kleemann | 2020 | Exercise prescription for people with mental illness: an evaluation of mental health professionals' knowledge, beliefs, barriers, and behaviors | Mental health recovery program / exercise referral scheme |
| Leavitt | 2017 | Improving Exercise Prescribing in a Rural New England Free Clinic | Mental health recovery program / exercise referral scheme |
| Lindsay | 2016 | Social prescribing: Leg Clubs - a collaborative example | Study type not suited |
| Lu | 2017 | Effect of Solution-Focused Brief Therapy-Based on Exercise Prescription Intervention on Adolescent Mental health | Not written in Dutch or English |
| Maier | 2016 | Promoting Nature-Based Activity for People With Mental Illness Through the US "Exercise Is Medicine" Initiative | Study type not suited |
| Martin-Domench | 2021 | [Evaluation of a pilot program of physical activity prescription in primary care in the Valencian Community (Spain)] | Not written in Dutch or English |
| Maund | 2019 | Wetlands for Wellbeing: Piloting a Nature-Based Health Intervention for the Management of Anxiety and Depression | Patients not referred from primary care |
| McHale | 2020 | Green Health Partnerships in Scotland; Pathways for Social Prescribing and Physical Activity Referral | Not focused on the influence of SP (on quadruple aim outcomes) |
| McLoughlin | 2019 | An integrated care approach to the users of social prescribing in an acutely frail older adult cohort | Not focused on the influence of SP (on quadruple aim outcomes) |
| Mendes | 2021 | Social prescribing in the community | Study type not suited |
| Mercer | 2017 | The Glasgow ‘Deep End’ Links Worker Study Protocol: a quasi-experimental evaluation of a social prescribing intervention for patients with complex needs in areas of high socioeconomic deprivation | Other |
| Miller | 2009 | Quality Zone: Yoga for emotional wellbeing in diabetes and other long-term conditions | Mental health recovery program / exercise referral scheme |
| Moffat | 2017 | Link Worker social prescribing to improve health and well-being for people with long-term conditions: qualitative study of service  user perceptions | Referral reasons not suited |
| Moffat | 2019 | Evaluating the impact of a community-based social prescribing intervention on people with type 2 diabetes in North East England: mixed-methods study protocol | Other |
| Mulligan | 2020 | Social Prescribing: Creating Pathways Towards Better Health and Wellness | Study type not suited |
| Munford | 2020 | Community asset participation and social medicine increases qualities of life | Patients not referred from primary care |
| Munford | 2020 | Effects of participating in community assets on quality of life and costs of care: longitudinal cohort study of older people in England | Patients not referred from primary care |
| Naylor | 2010 | Bibliotherapy as a Treatment for Depression in Primary Care | Mental health recovery program / exercise referral scheme |
| Neville | 2013 | Prose not Prozac? The role of book prescription schemes and healthy reading schemes in the treatment of mental illness in Ireland | Mental health recovery program / exercise referral scheme |
| Neville | 2010 | The Reading Cure? Bibliotherapy, Healthy Reading Schemes and the Treatment of Mental Illness in Ireland | Mental health recovery program / exercise referral scheme |
| NICE | 2021 | Evidence review for social interventions for chronic pain (chronic primary pain and chronic secondary pain) | Study type not suited |
| Nyatanga | 2020 | Social prescribing: combating loneliness is everyone's business | Other |
| Orellana | 2020 | Day centres for older people - attender characteristics, access routes and outcomes of regular attendance: findings of exploratory mixed methods case study research | Patients not referred from primary care |
| O'Toole | 2018 | The Efficacy of Exercise Referral as an Intervention for Irish Male Prisoners Presenting with Mental Health Symptoms | Mental health recovery program / exercise referral scheme |
| Patel | 2021 | Opportunities and Challenges for Digital Social Prescribing in Mental Health: Questionnaire Study | Not focused on the influence of SP (on quadruple aim outcomes) |
| Patel | 2021 | Correction: Opportunities and Challenges for Digital Social Prescribing in Mental Health: Questionnaire Study | Other |
| Patterson | 2013 | Considering referral to art therapy: Responses to referral and experiences of participants in a randomised controlled trial | Patients not referred from primary care |
| Patterson | 2013 | Considering referral to art therapy: Responses to referral and experiences of participants in a randomised controlled trial | Patients not referred from primary care (duplicate) |
| Pescheny | 2018 | Patient uptake and adherence to social prescribing: a qualitative study | Not focused on the influence of SP (on quadruple aim outcomes) |
| Pescheny | 2018 | Social Prescribing: Primary care patient and service user engagement | Not focused on the influence of SP (on quadruple aim outcomes) |
| Pescheny | 2018 | Evaluating the Implementation and Delivery of a Social Prescribing Intervention: A Research Protocol | Other |
| Pescheny | 2018 | Social Prescribing: Implementation and delivery | Not focused on the influence of SP (on quadruple aim outcomes) |
| Pescheny | 2018 | [Poster abstract] Service user outcomes of a social prescribing programme in general practice | Other |
| Pickering | 2021 | Social prescribing: a nurse-led pilot project in a general practice setting | Study type not suited |
| Popay | 2007 | Social problems, primary care and pathways to help and support: addressing health inequalities at the individual level. Part II: lay perspectives | Patients not referred from primary care (duplicate) |
| Popay | 2007 | Social problems, primary care and pathways to help and support: addressing health inequalities at the individual level. Part I: the GP perspective | Patients not referred from primary care (duplicate) |
| Pretty | 2020 | Nature-Based Interventions and Mind–Body Interventions: Saving Public Health Costs Whilst Increasing Life Satisfaction and Happiness | Study type not suited |
| Puebla Fortier | 2021 | Creative cross-sectoral collaboration: a conceptual framework of factors influencing partnerships for arts, health and wellbeing | Not focused on the influence of SP (on quadruple aim outcomes) |
| Radovic | 2018 | Clinician perspectives and practices regarding the use of exercise in the treatment of adolescent depression | Mental health recovery program / exercise referral scheme |
| Rhodes | 2021 | 'It sounded a lot simpler on the job description'': A qualitative study exploring the role of social prescribing link workers and their training and support needs | Not focused on the influence of SP (on quadruple aim outcomes) |
| Roberts | 2021 | The role of social prescribers in wales: a consensus methods study | Not focused on the influence of SP (on quadruple aim outcomes) |
| Roessler | 2011 | A corrective emotional experience - or just a bit of exercise? The relevance of interpersonal learning in Exercise on prescription | Mental health recovery program / exercise referral scheme |
| Rogers | 2012 | Physician-prescribed physical activity in older adults | Study type not suited |
| Shaikh | 2021 | Socio-economic inequalities in arts engagement and depression among older adults in the United Kingdom: evidence from the English Longitudinal Study of Ageing | Not focused on the influence of SP (on quadruple aim outcomes) |
| Skivington | 2018 | Delivering a primary care-based social prescribing initiative: a qualitative study of the benefits and challenges | Not focused on the influence of SP (on quadruple aim outcomes) |
| Smock | 2020 | Assessing Factors That Influence Healthcare Provider Attitudes and Practices regarding Place-Based Exercise Prescriptions: Results of Principal Components Analysis of a Newly Developed Survey Instrument | Other |
| South | 2008 | Can social prescribing provide the missing link? | Before 2010 |
| Southby | 2018 | Factors affecting general practice collaboration with voluntary and community sector organisations | Not focused on the influence of SP (on quadruple aim outcomes) |
| Stanton | 2015 | A Pilot Study of the Views of General Practitioners Regarding Exercise for the Treatment of Depression | Mental health recovery program / exercise referral scheme |
| Stanton | 2013 | An Exercise Prescription Primer for People with Depression | Mental health recovery program / exercise referral scheme |
| Stanton | 2018 | Implementation in action: how Australian Exercise Physiologists approach exercise prescription for people with mental illness | Mental health recovery program / exercise referral scheme |
| Stickley | 2010 | Does prescribing participation in arts help to promote recovery for mental health clients? | Mental health recovery program / exercise referral scheme |
| Stickley | 2012 | Social prescribing through arts on prescription in a UK city: Participants’ perspectives (Part 1) | Patients not referred from primary care |
| Stickley | 2012 | Social prescribing through arts on prescription in a UK city: Participants’ perspectives (Part 2) | Patients not referred from primary care |
| Stickley | 2013 | Arts on prescription: a qualitative outcomes study | Patients not referred from primary care |
| Stuart | 2021 | ‘Oh no, not a group!’ The factors that lonely or isolated people report as barriers to joining groups for health and well-being | Not focused on the influence of SP (on quadruple aim outcomes) |
| Swift | 2017 | People powered primary care: learning from Halton | Study type not suited |
| Taylor | 2019 | Perceptions of Pharmacy Involvement in Social Prescribing Pathways in England, Scotland and Wales | Not focused on the influence of SP (on quadruple aim outcomes) |
| Thomas | 2020 | Stepped-Wedge Cluster Randomised Trial of Social Prescribing of Forest Therapy for Quality of Life and Biopsychosocial Wellbeing in Community-Living Australian Adults with Mental Illness: Protocol | Other |
| Thomson | 2018 | Effects of a museum-based social prescription intervention on quantitative measures of psychological wellbeing in older adults | Mental health recovery program / exercise referral scheme |
| Thomson | 2020 | Art, nature and mental health: assessing the biopsychosocial effects of a ‘creative green prescription’ museum programme involving horticulture, artmaking and collections | Mental health recovery program / exercise referral scheme |
| Tierney | 2019 | Current understanding and implementation of ‘care navigation’ across England: a cross-sectional study of NHS clinical commissioning groups | Not focused on the influence of SP (on quadruple aim outcomes) |
| Todd | 2017 | Museum-based programs for socially isolated older adults: Understanding what works | Not focused on the influence of SP (on quadruple aim outcomes) |
| Unknwon | 2012 | Physical activity is not a panacea for depression after all | Mental health recovery program / exercise referral scheme |
| Unknown | Unknown | A Mental Health Social Prescribing Trial (British Red Cross) | Other |
| Unknown | Unknown | Improving Depressive Symptoms Through Personalised Exercise and Activation | Other |
| Vancampfort | 2019 | A quantitative assessment of the views of mental health professionals on exercise for people with mental illness: perspectives from a low-resource setting | Mental health recovery program / exercise referral scheme |
| Vannier | 2021 | Strengthening community connection and personal well‐being through volunteering in New Zealand | Patients not referred from primary care |
| Waddington-Jones | 2019 | Exploring Wellbeing and Creativity Through Collaborative Composition as Part of Hull 2017 City of Culture. | Patients not referred from primary care |
| Wallace | 2021 | Using consensus methods to develop a Social Prescribing Learning Needs Framework for practitioners in Wales | Not focused on the influence of SP (on quadruple aim outcomes) |
| Ward | 2020 | Social prescribing by students: the design and delivery of a social prescribing scheme by medical students in general practice | Not focused on the influence of SP (on quadruple aim outcomes) |
| White | 2017 | Front-line perspectives on 'joined-up' working relationships: a qualitative study of social prescribing in the west of Scotland | Not focused on the influence of SP (on quadruple aim outcomes) |
| Whitelaw | 2017 | Developing and implementing a social prescribing initiative in primary care: insights into the possibility of normalisation and sustainability from a UK case study | Not focused on the influence of SP (on quadruple aim outcomes) |
| Wildman | 2019 | Link workers' perspectives on factors enabling and preventing client engagement with social prescribing | Referral reasons not suited |
| Wildman | 2019 | Service-users’ perspectives of link worker social prescribing: a qualitative follow-up study | Referral reasons not suited |
| Wildman | 2021 | Evaluation of a Community Health Worker Social Prescribing Program Among UK Patients With Type 2 Diabetes | Referral reasons not suited |
| Wilkinson | 2013 | Visible Voices: Expressive arts with isolated seniors using trained volunteers | Patients not referred from primary care |
| Wilkinson | 2021 | A collaborative, multi-sectoral approach to implementing a social prescribing initiative to alleviate social isolation and enhance well-being amongst older people | Study type not suited |
| Wood | 2021 | Social prescribing for people with complex needs: a realist evaluation | Not focused on the influence of SP (on quadruple aim outcomes) |
| Yates | 2020 | Prescribed exercise for the treatment of depression in a college population: An interprofessional approach | Mental health recovery program / exercise referral scheme |
| Zhu | 2020 | An Evaluation of Connect for Health: A Social Referral Program in RI | Not focused on the influence of SP (on quadruple aim outcomes) |
| **SEARCH UPDATE** | | | |
| Al-Khudairy | 2022 | Evidence and methods required to evaluate the impact for patients who use social prescribing: a rapid systematic review and qualitative interviews | Study type not suited |
| Brettell | 2022 | Linking Leeds: A Social Prescribing Service for Children and Young People | Other |
| Burns | 2022 | Social prescribing pilot costing 12.7m aims to cut medication use through walking and cycling | Study type not suited |
| Cheshire | 2022 | 'Joining a group was inspiring': a qualitative study of service users' experiences of yoga on social prescription | Not focused on the influence of SP (on quadruple aim outcomes) |
| Fixsen | 2022 | Challenges and Approaches to Green Social Prescribing During and in the Aftermath of COVID-19: A Qualitative Study | Not focused on the influence of SP (on quadruple aim outcomes) |
| Gorenberg | 2023 | Understanding and Improving Older People's Well-Being through Social Prescribing Involving the Cultural Sector: Interviews from a Realist Evaluation | Not focused on the influence of SP (on quadruple aim outcomes) |
| Irwin | 2022 | Evaluation of a gallery-based Arts Engagement program for depression | Other |
| Jensen | 2021 | Swedish primary healthcare practitioners' perspectives on the impact of arts on prescription for patients and the wider society: a qualitative interview study | Not focused on the influence of SP (on quadruple aim outcomes) |
| Makanjuola | 2022 | A Social Return on Investment Evaluation of the Pilot Social Prescribing EmotionMind Dynamic Coaching Programme to Improve Mental Wellbeing and Self-Confidence | Mental health recovery program / exercise referral scheme |
| Morris | 2022 | Social prescribing during the COVID-19 pandemic: a qualitative study of service providers' and clients' experiences | Patients not referred from primary care |
| Ohta | 2022 | A Solution for Loneliness in Rural Populations: The Effects of Osekkai Conferences during the COVID-19 Pandemic | Patients not referred from primary care |
| Pretty | 2020 | Nature-Based Interventions and Mind–Body Interventions: Saving Public Health Costs Whilst Increasing Life Satisfaction and Happiness | Patients not referred from primary care |
| Smyth | 2022 | Increased Wellbeing following Engagement in a Group Nature-Based Programme: The Green Gym Programme Delivered by the Conservation Volunteers | Patients not referred from primary care |
| Thomson | 2023 | Service Users’ Perspectives of a National Social Prescribing Programme to Address Loneliness and Social Isolation: A Qualitative Study | Other |
| Unknown | 2022 | Testing a Nature-based Socila Intevention on Loneliness: the RECETAS-PRG Trial | Other |
| Unknwon | 2022 | Testing a Nature-based Social Intervention on Loneliness: the RECETAS-BCN Trial | Other |
| Wachuku | 2021 | Social Prescribing for Health: Voucher Adherence and Measured Health Outcomes in a Prescription for Health Program | Patients not referred from primary care |
| White | 2022 | More than signposting: Findings from an evaluation of a social prescribing service | Not focused on the influence of SP (on quadruple aim outcomes) |
| Wildman and Wildman | 2023 | Impact of a link worker social prescribing intervention on non-elective admitted patient care costs: A quasi-experimental study | Referral reasons not suited |
