## AppendixF_StratifiedResults for "Intervention Characteristics and Mechanisms and their Relationship with the Influence of Social Prescribing: a Systematic Review"

**Appendix F.** Stratified results

Wellbeing outcomes

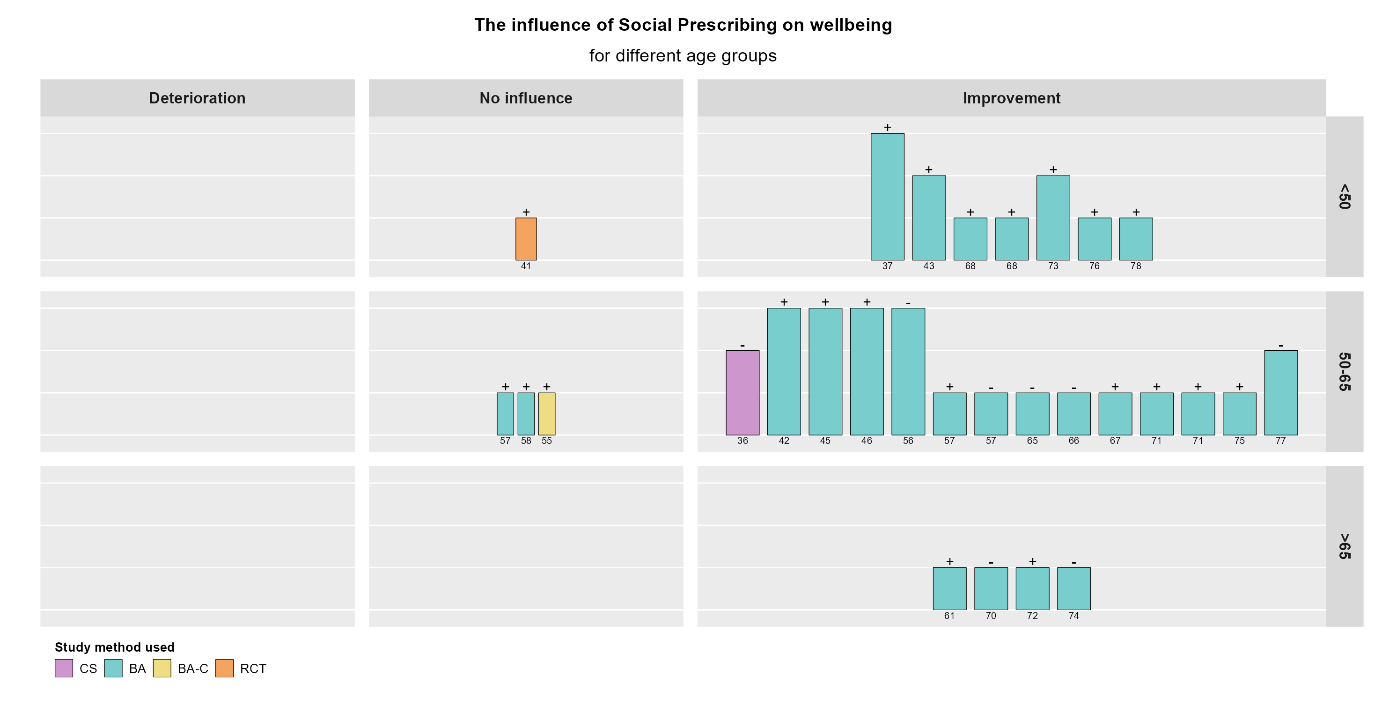

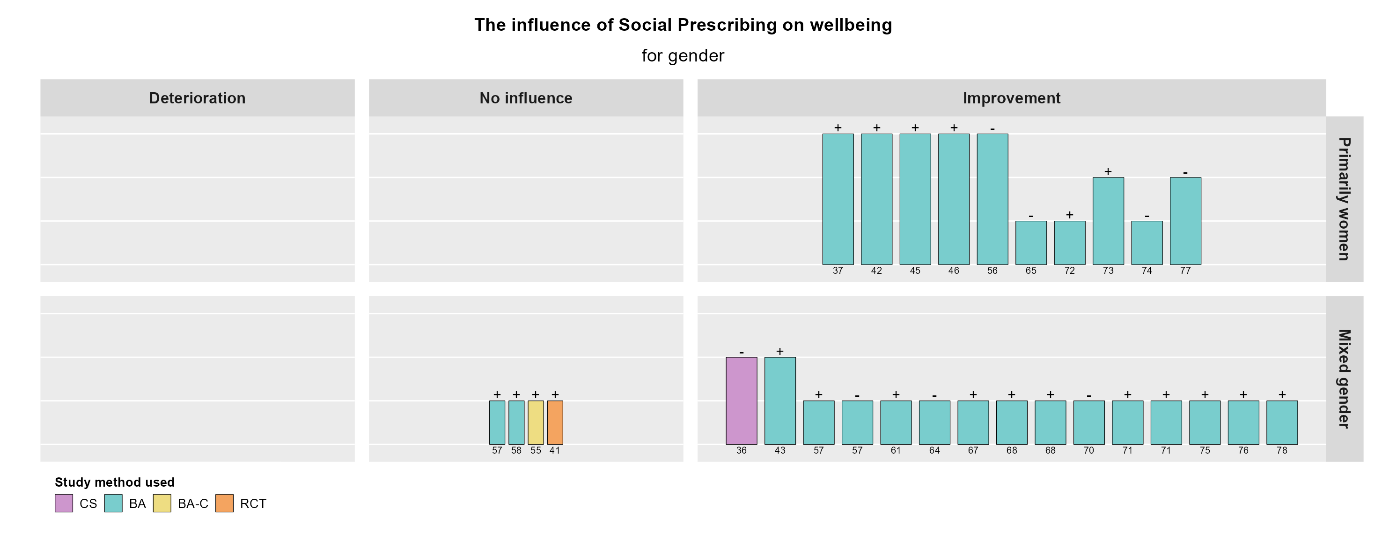

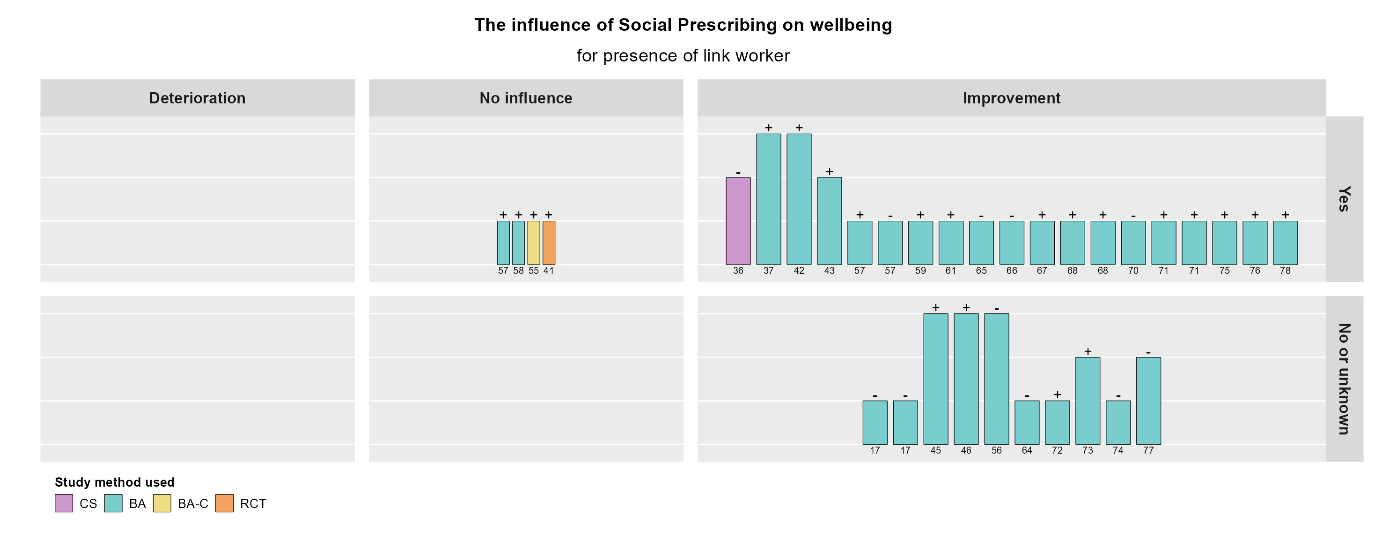

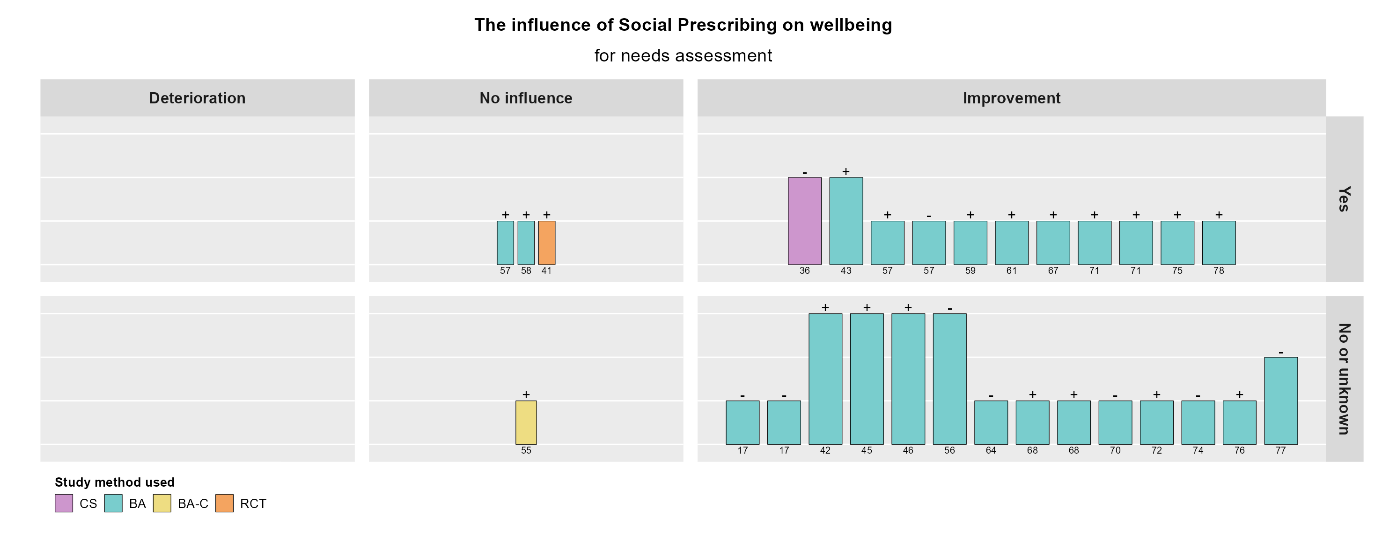

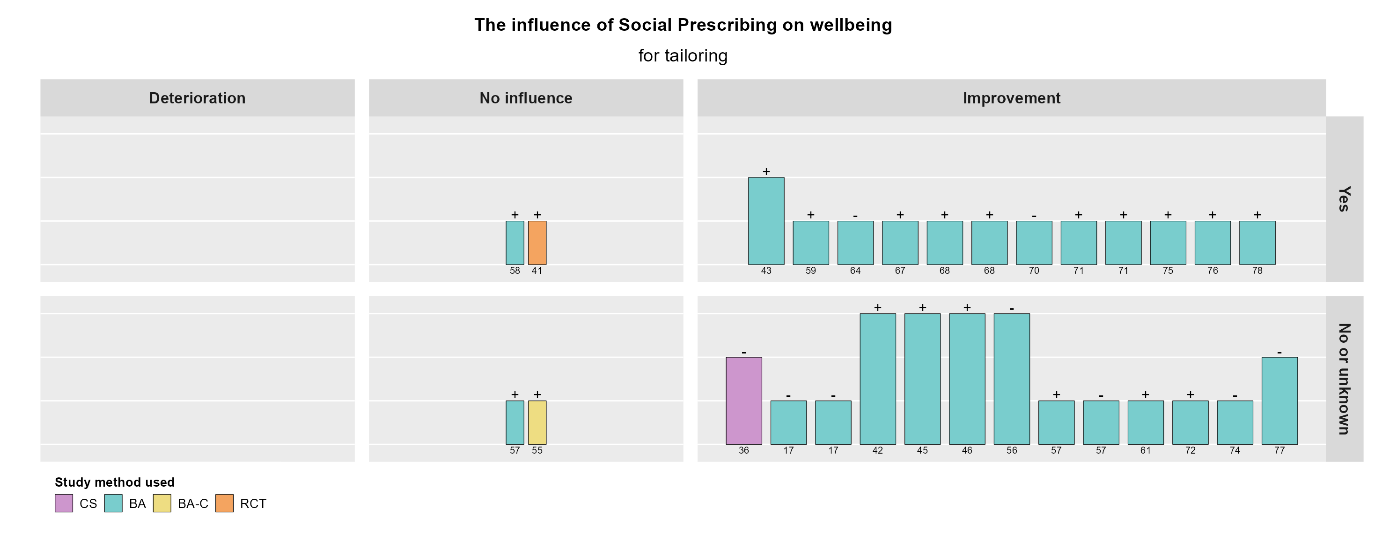

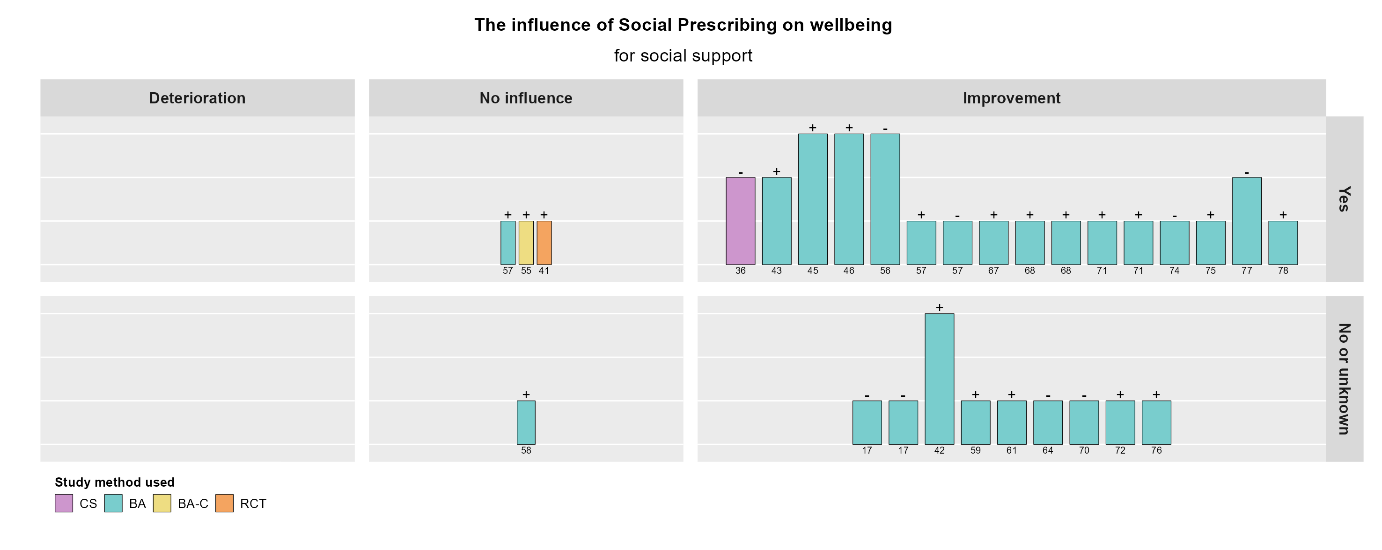

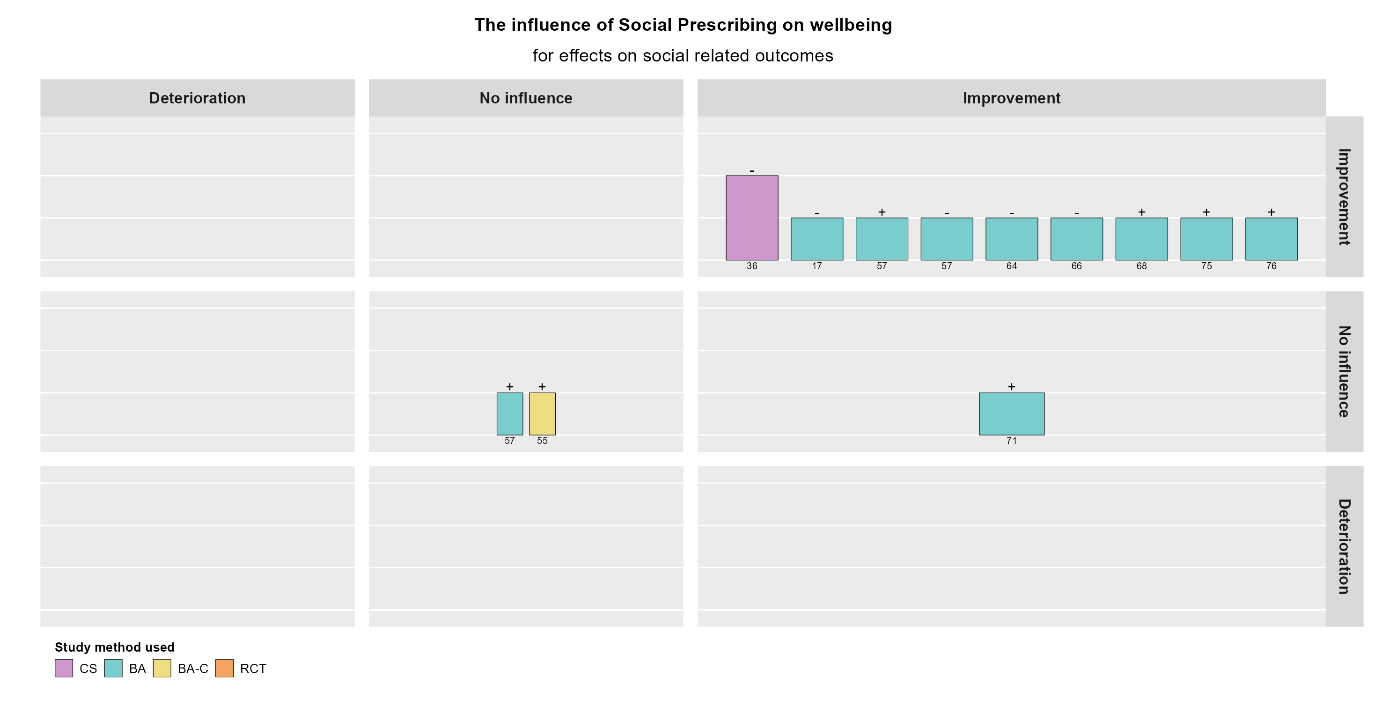

Mental health outcomes

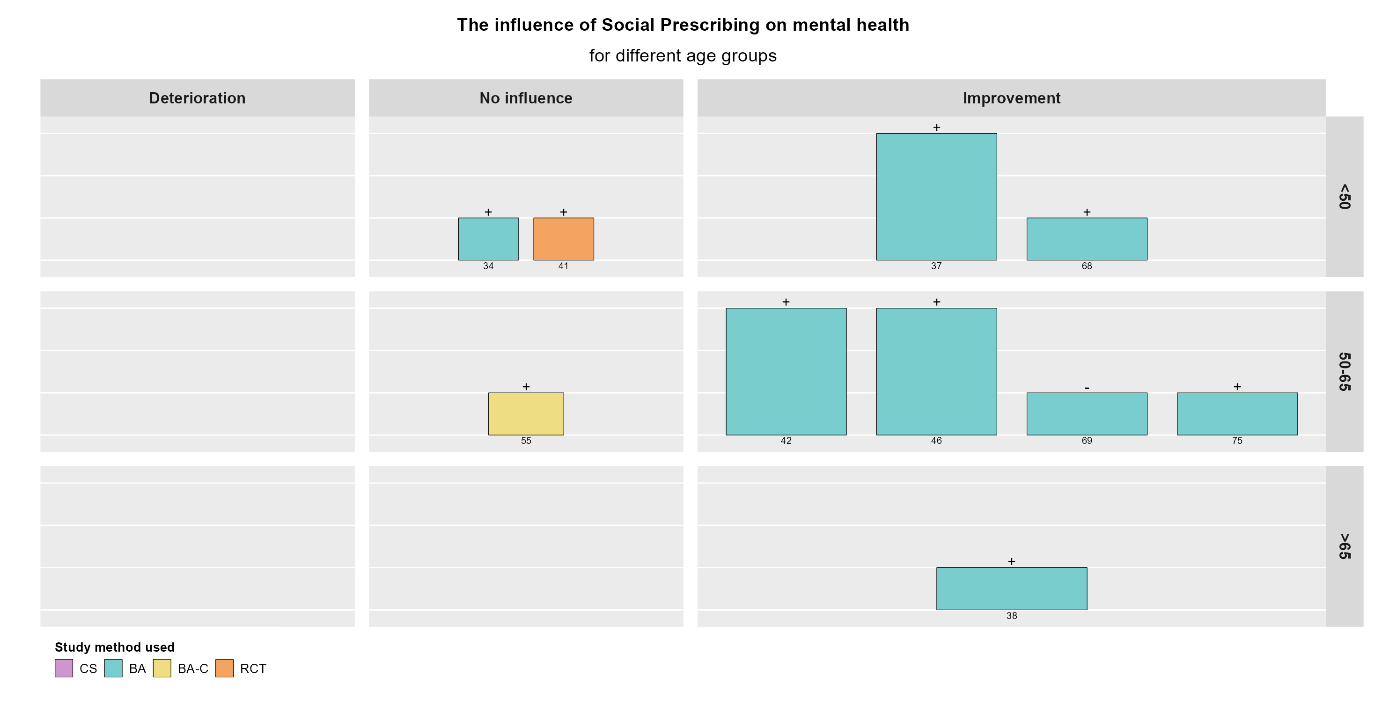

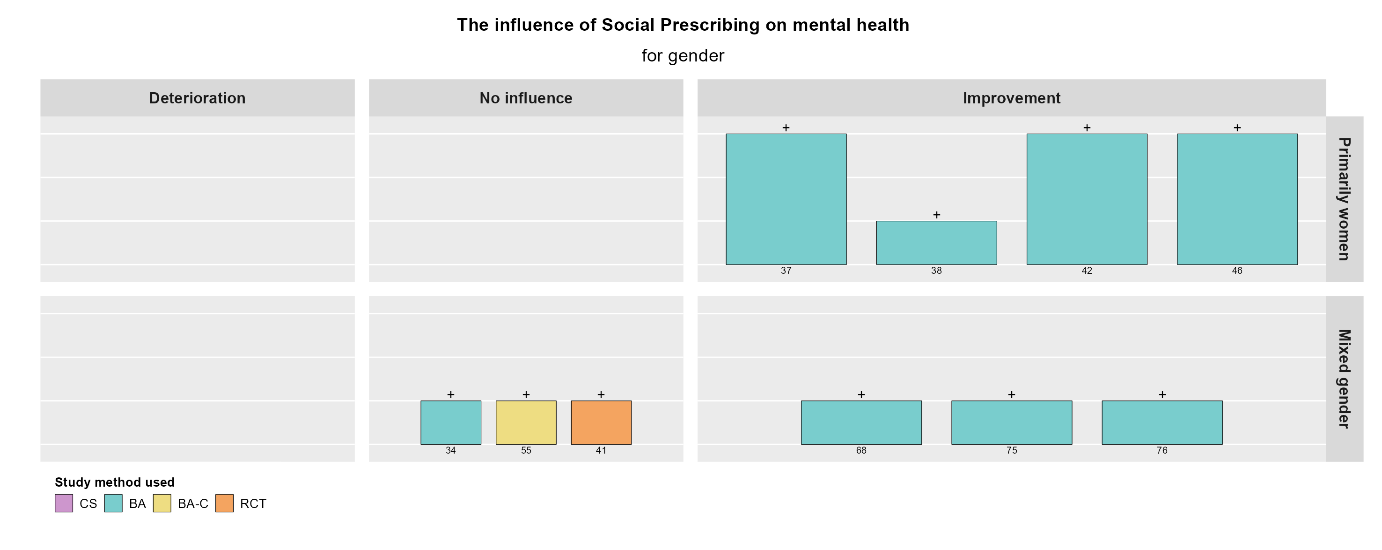

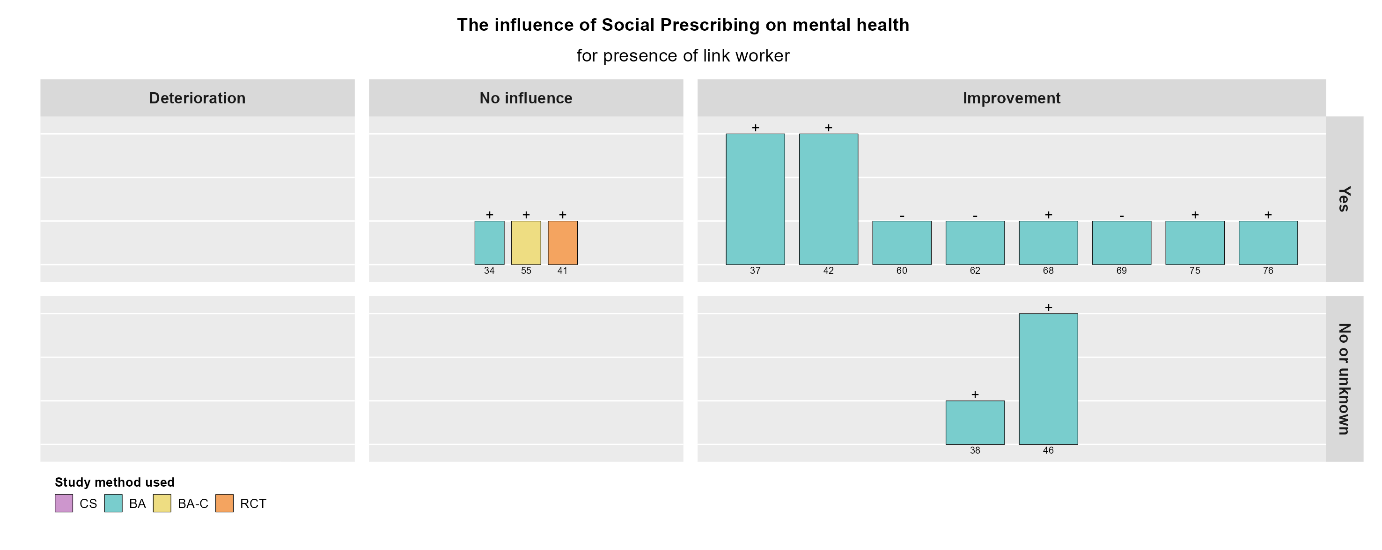

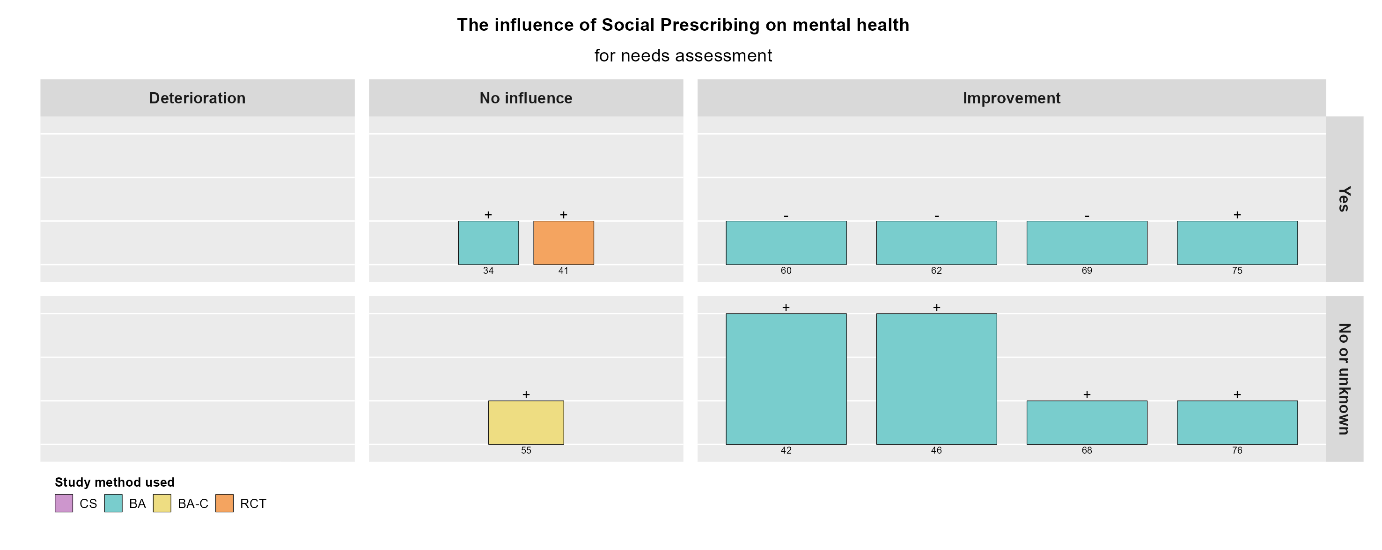

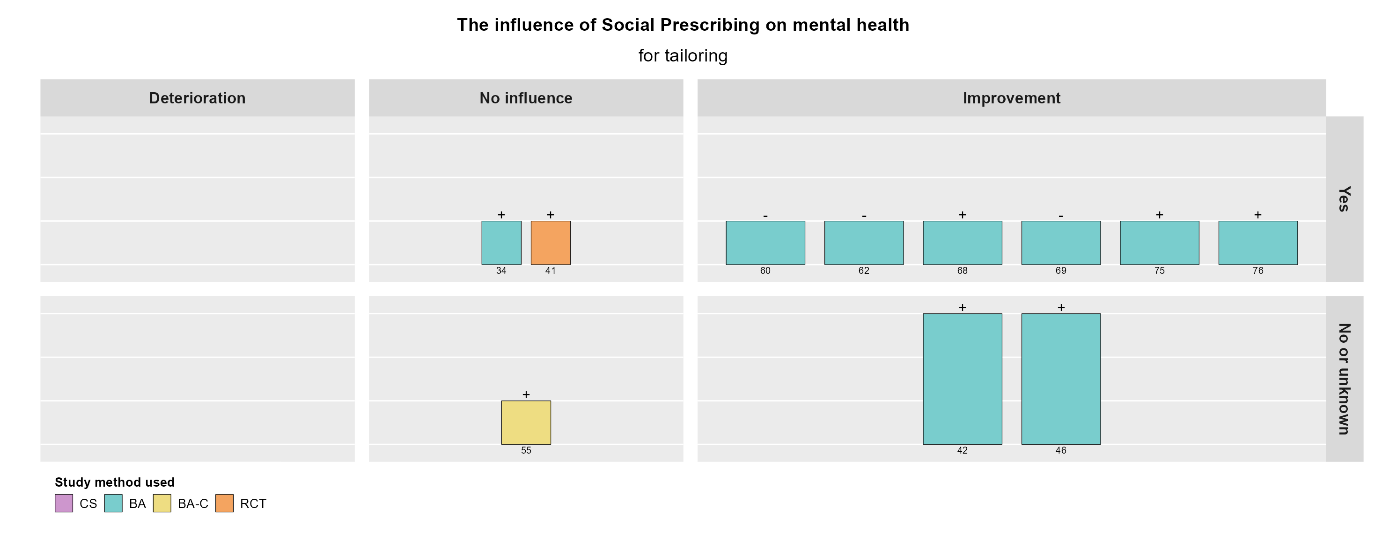

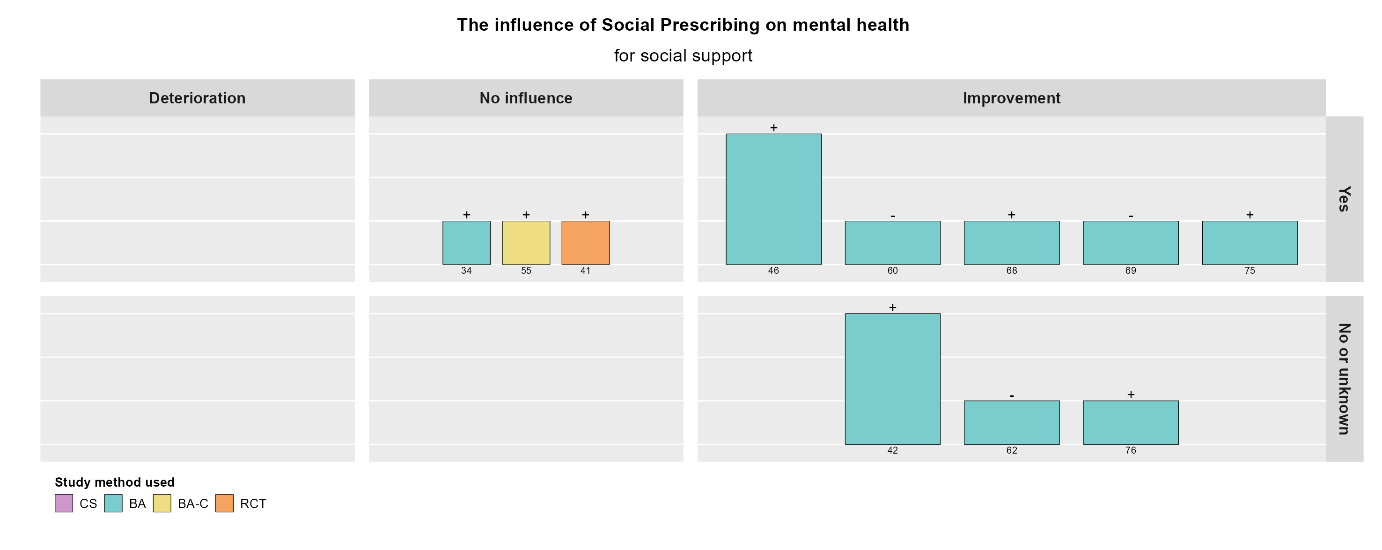

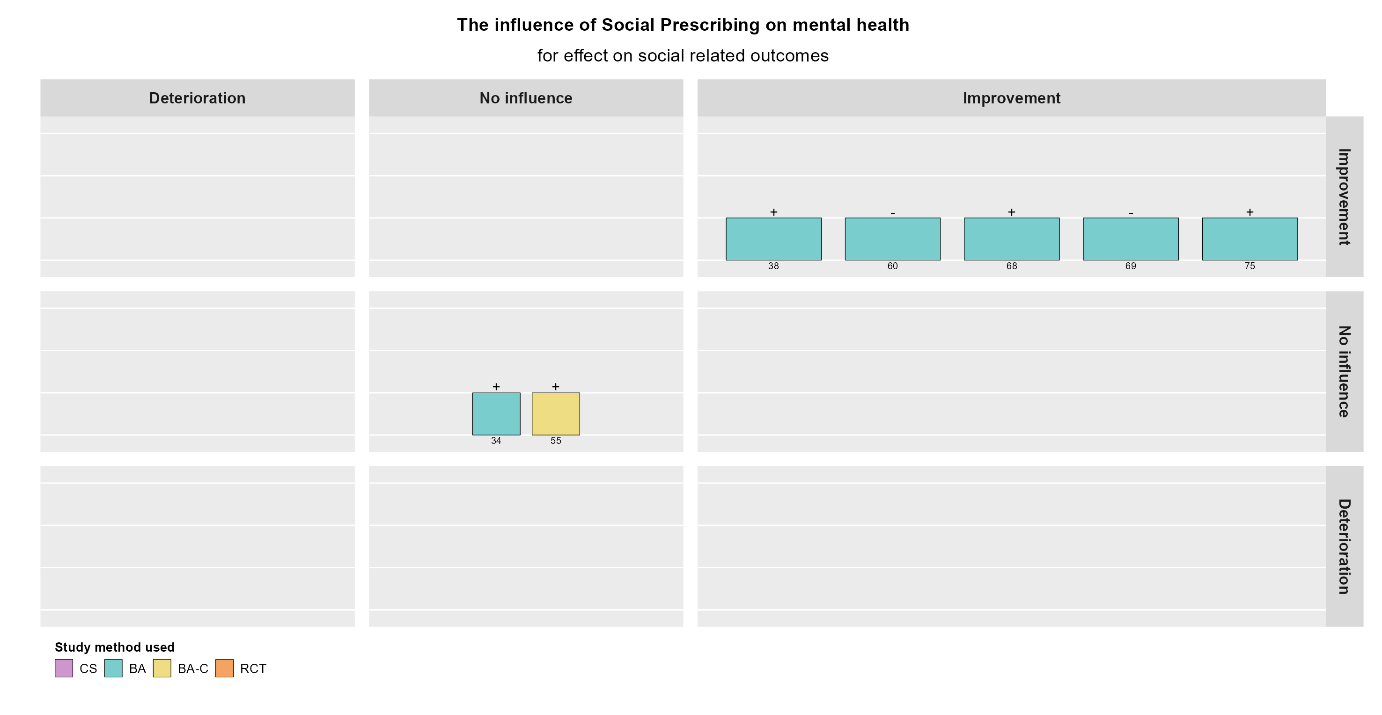

General and physical health and health behavior outcomes

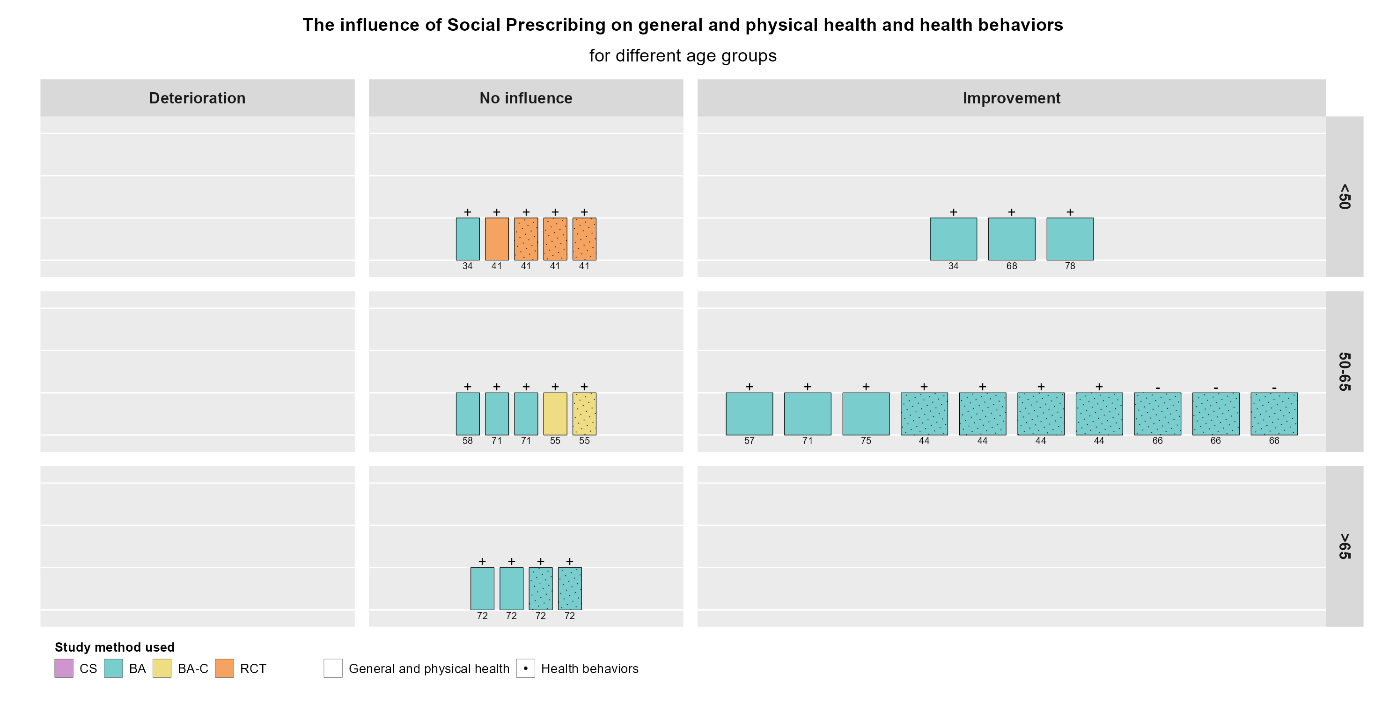

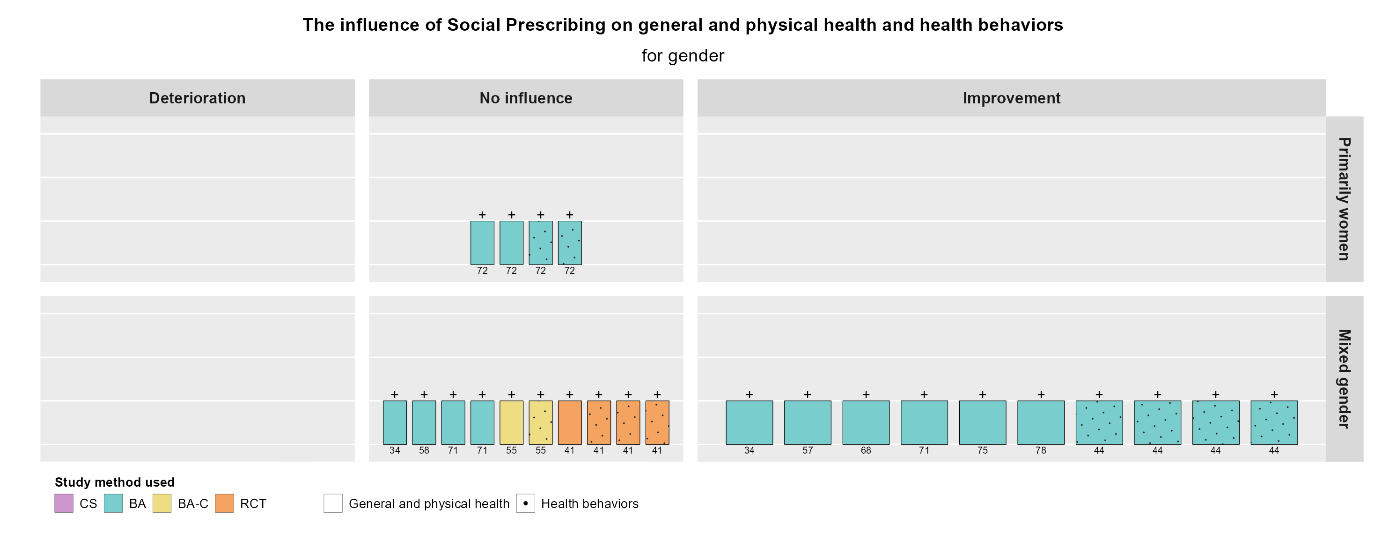

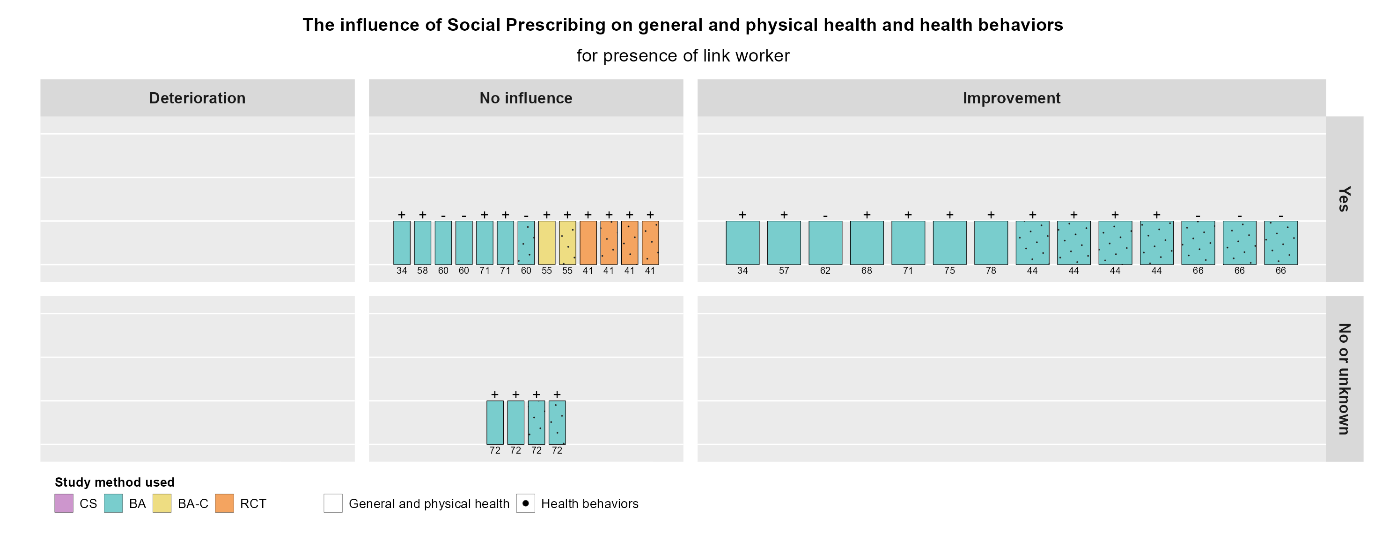

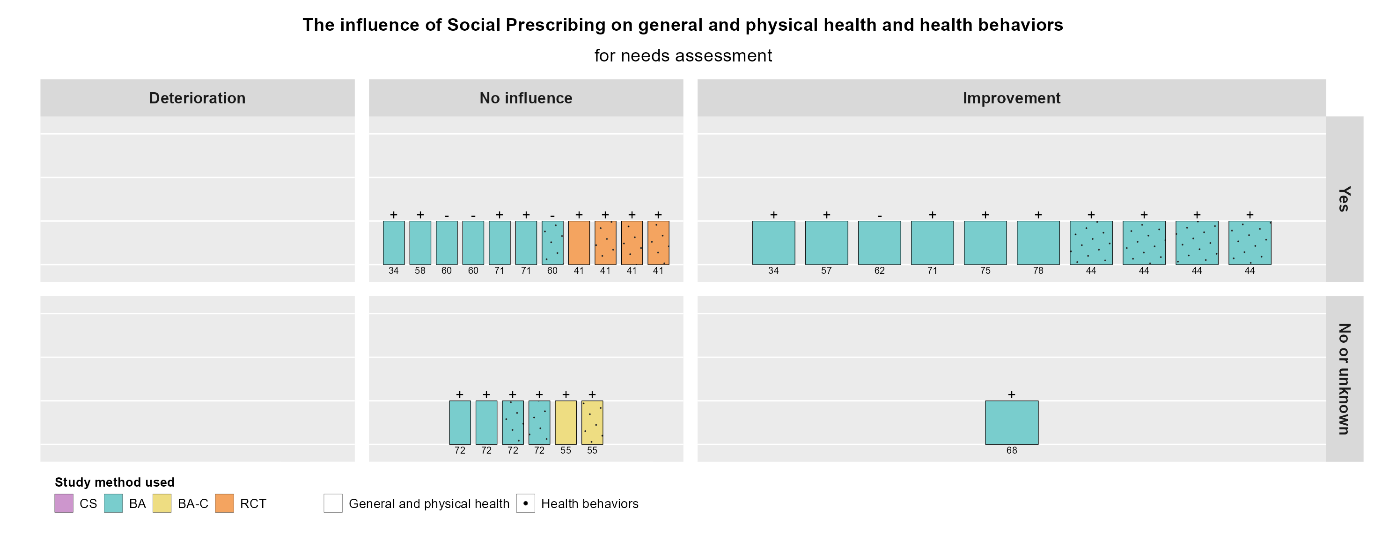

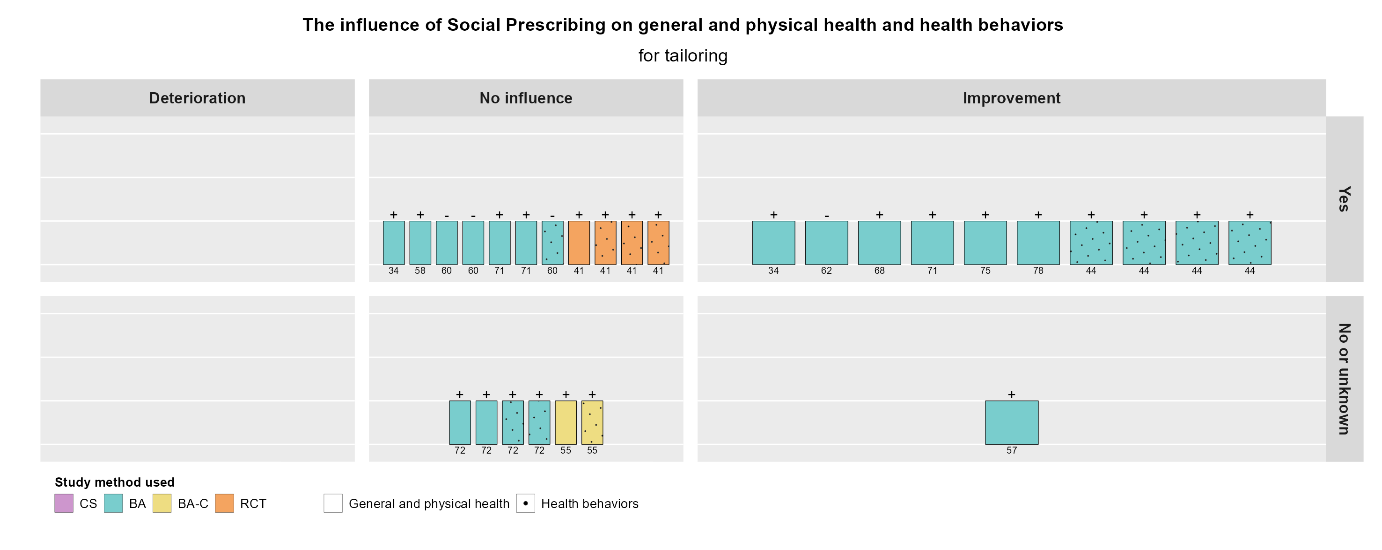

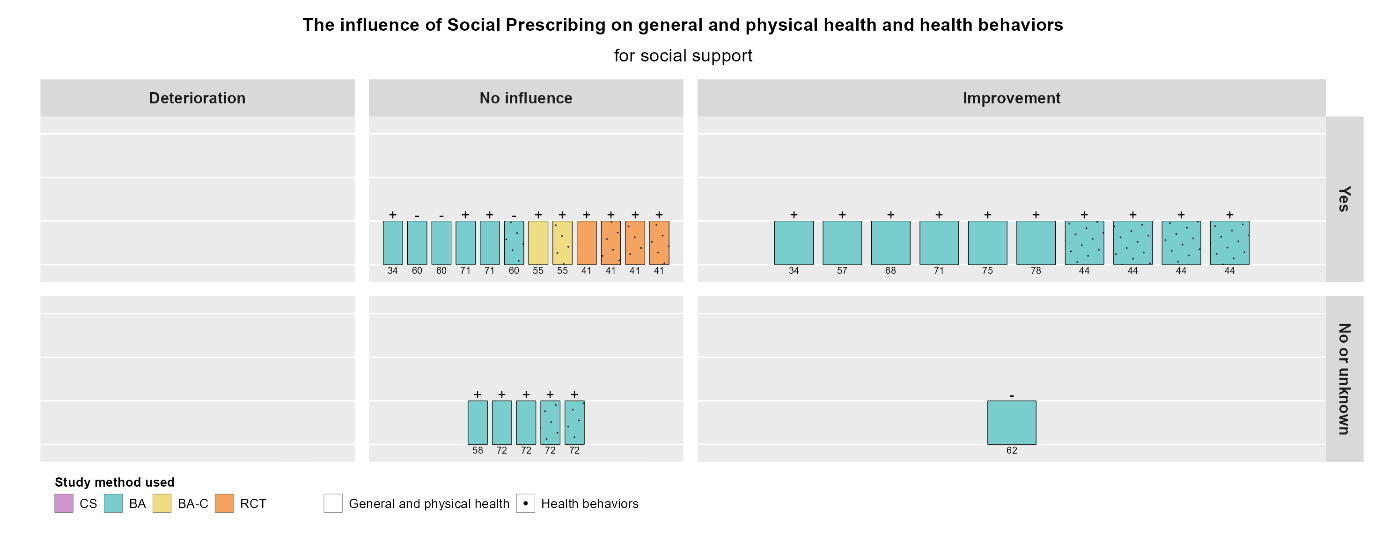

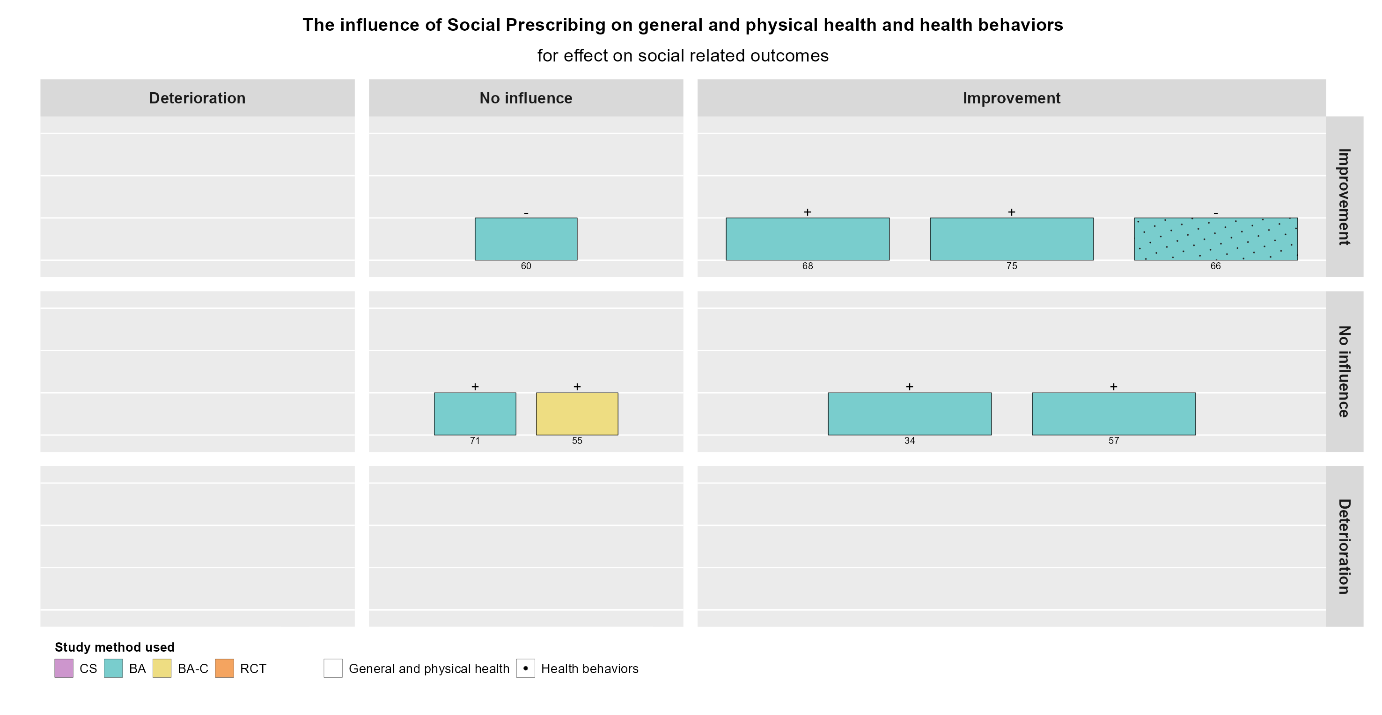

Primary care use

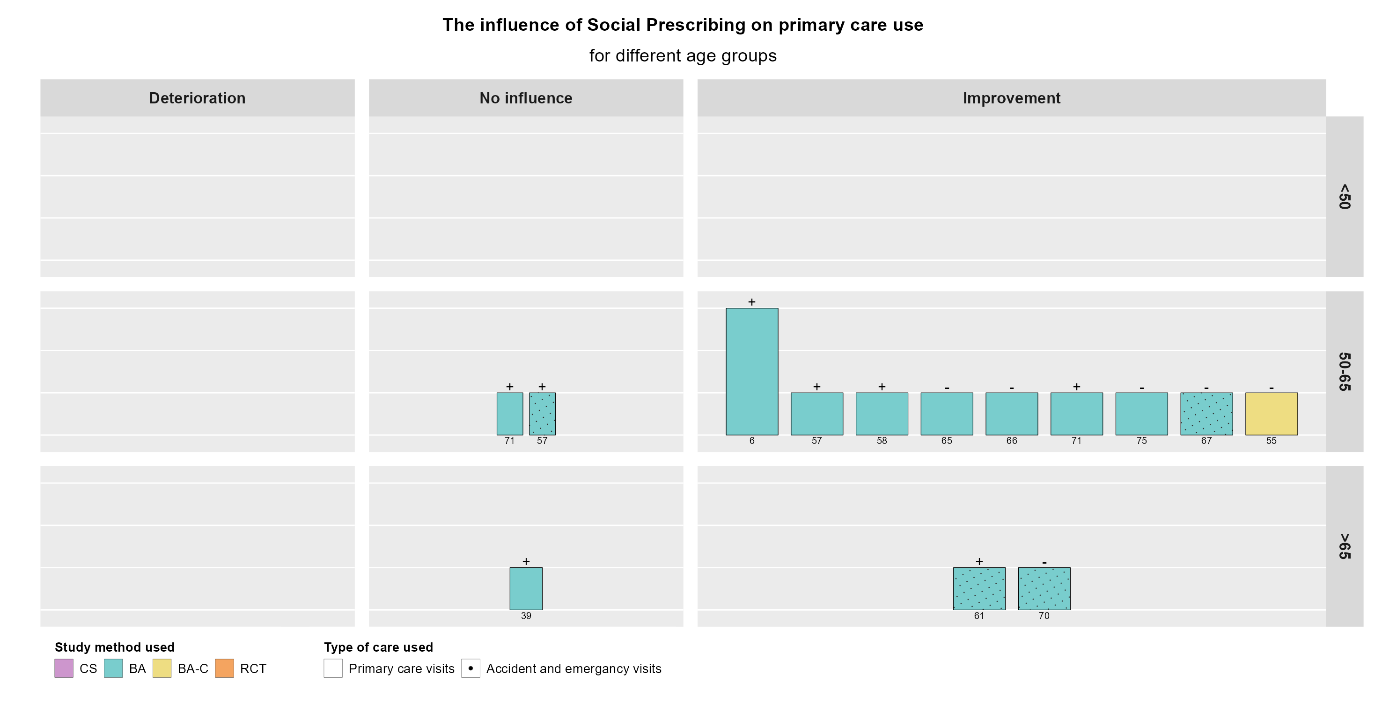

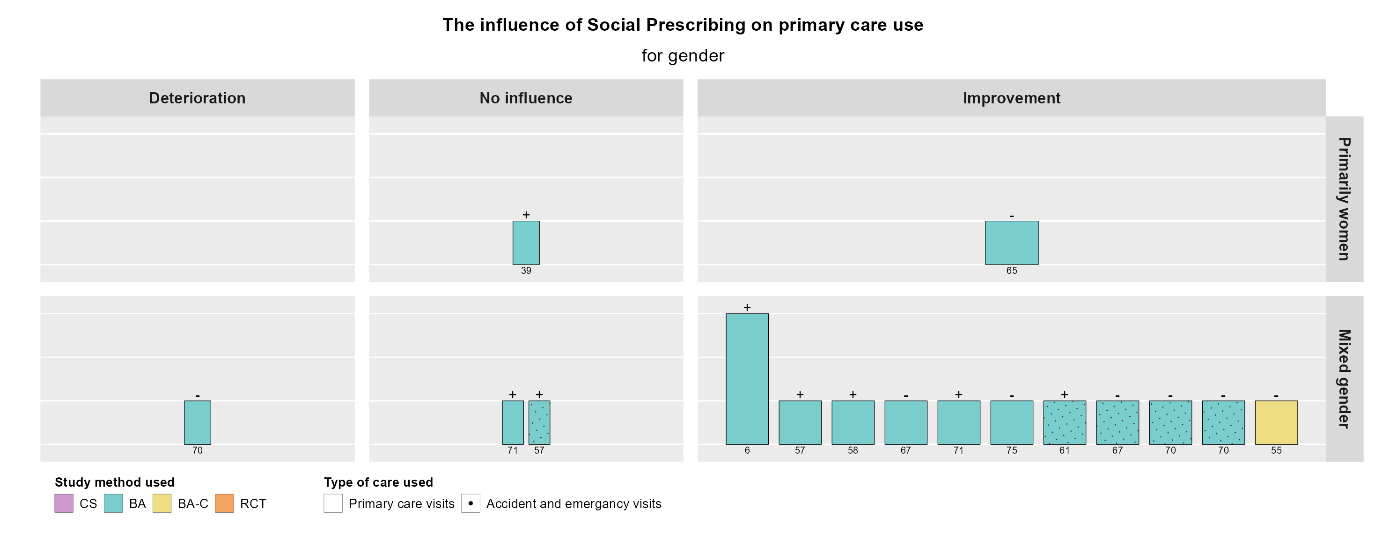

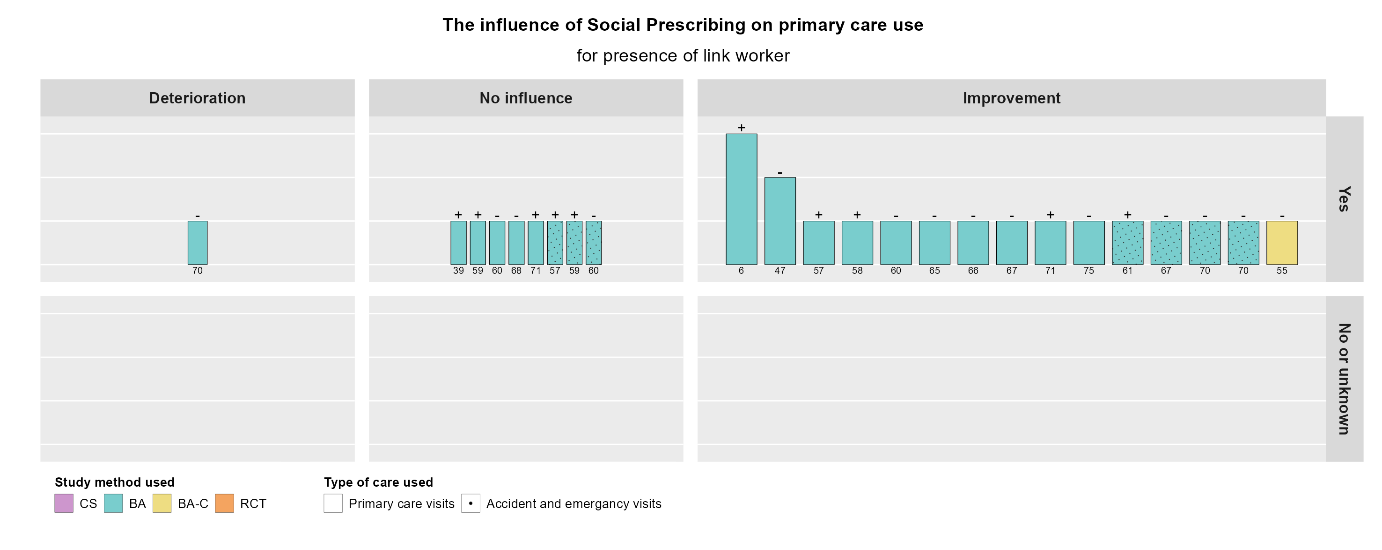

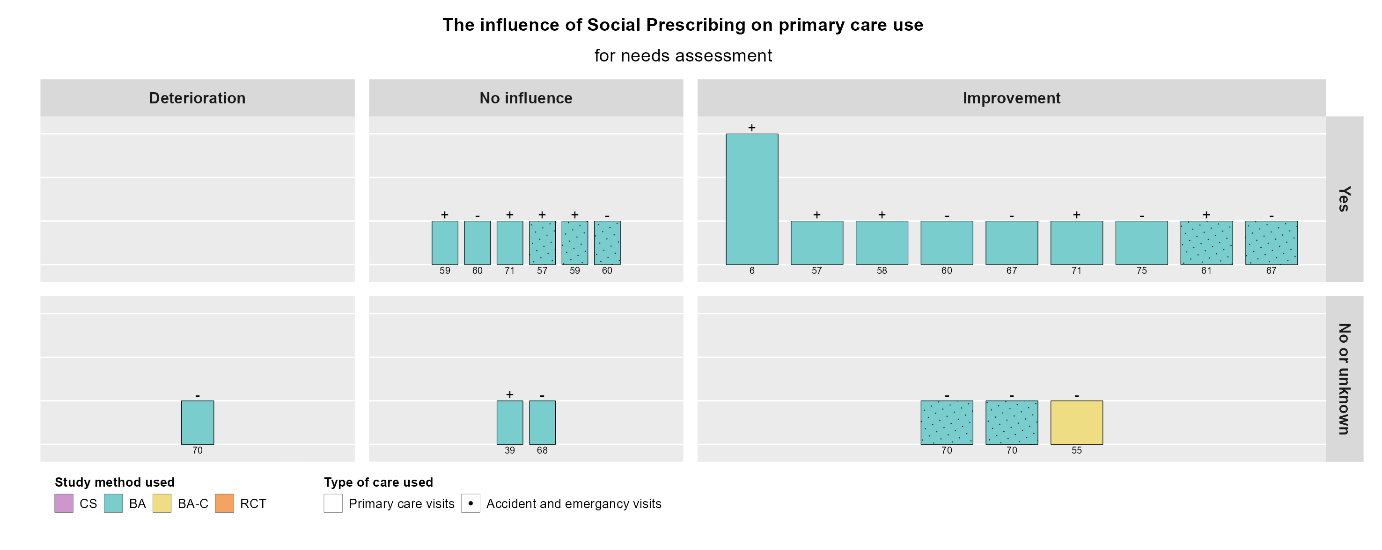

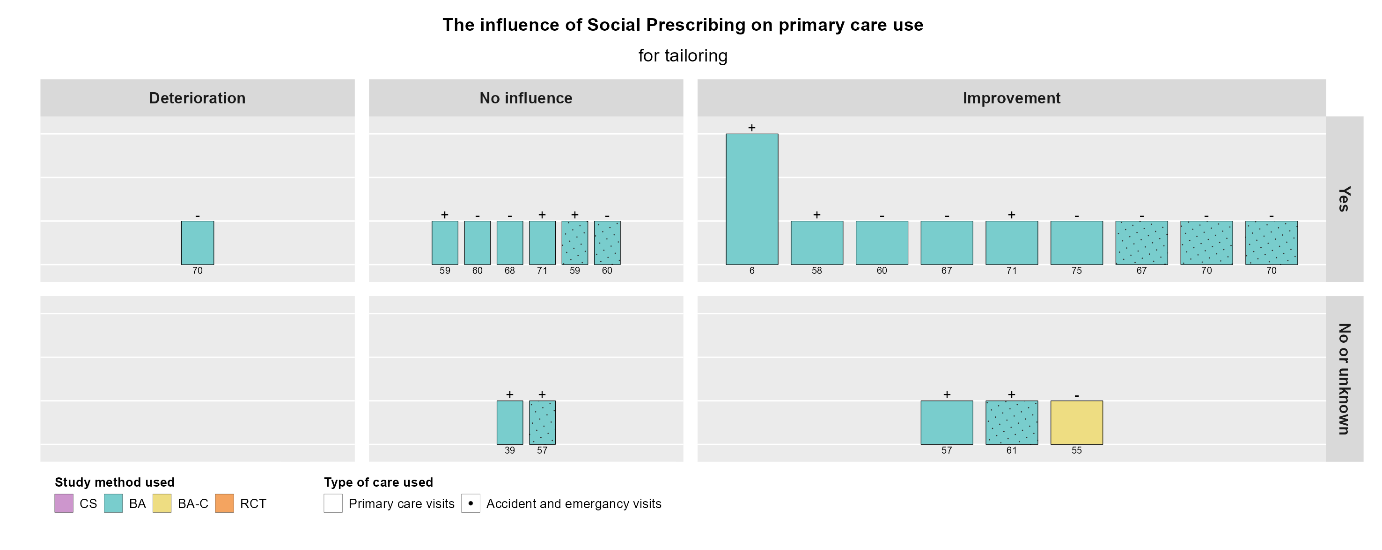

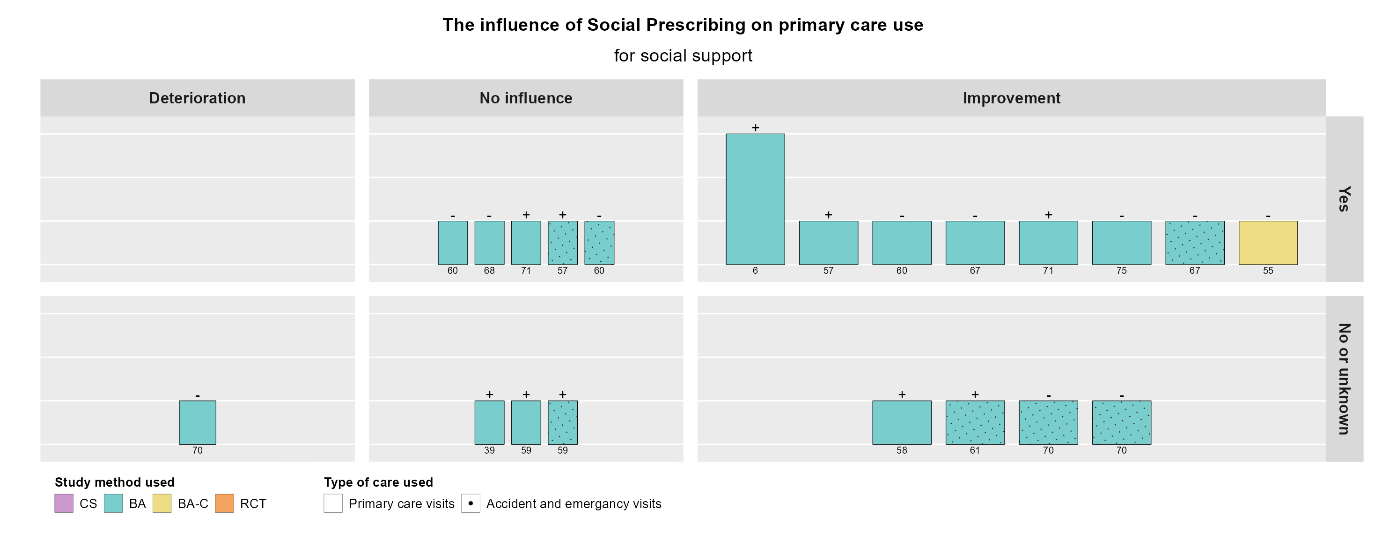

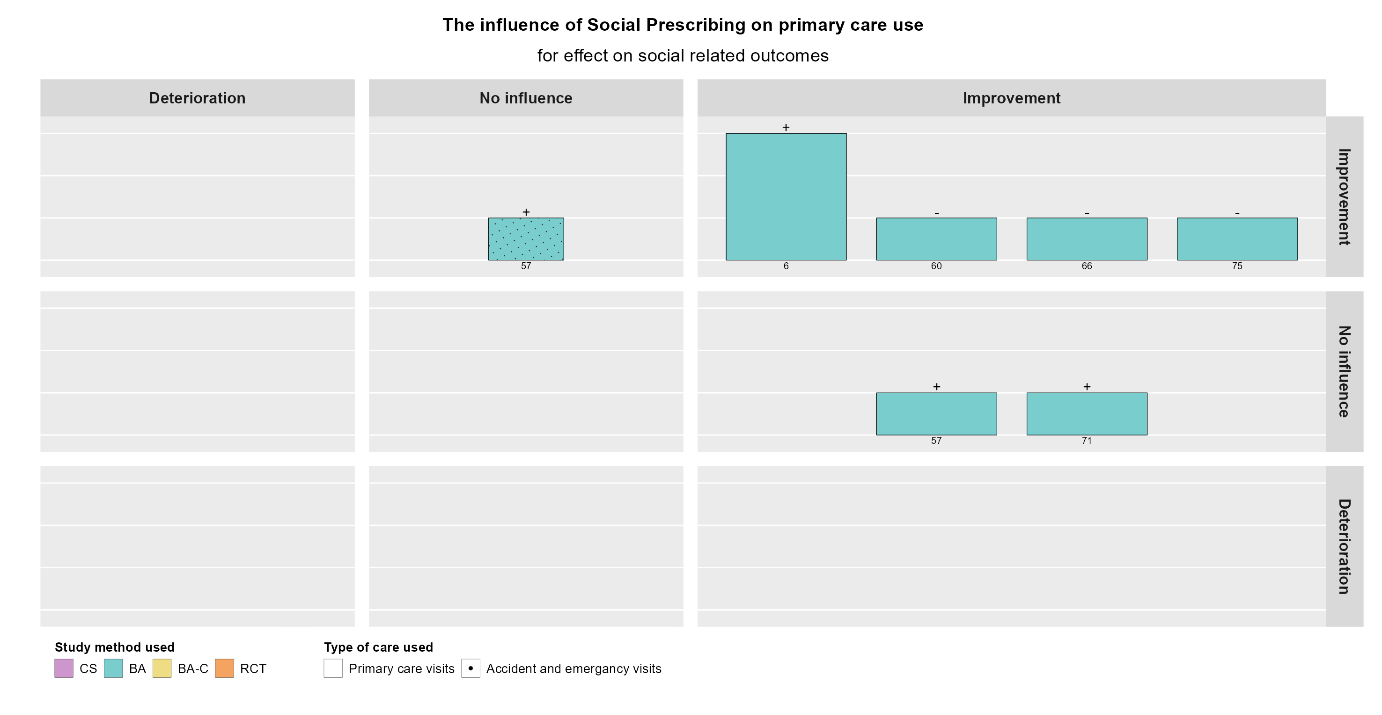

Social-related outcomes

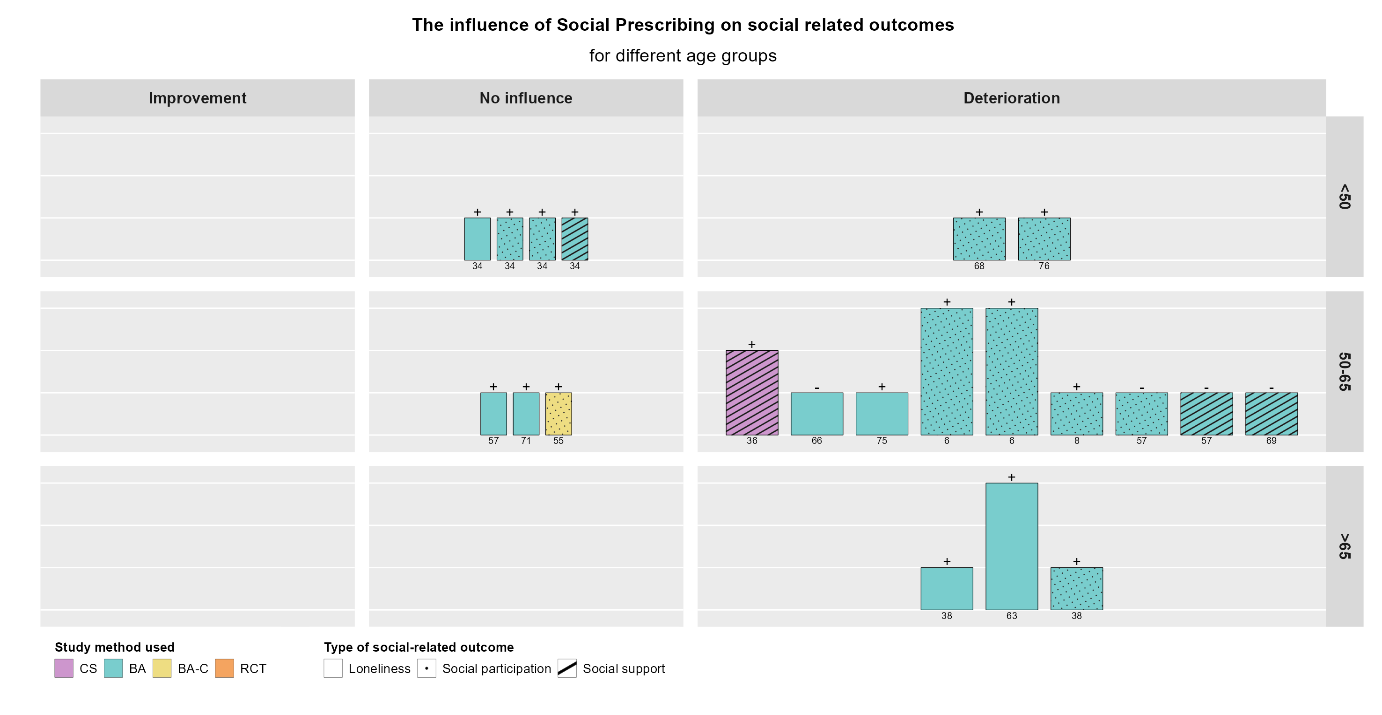

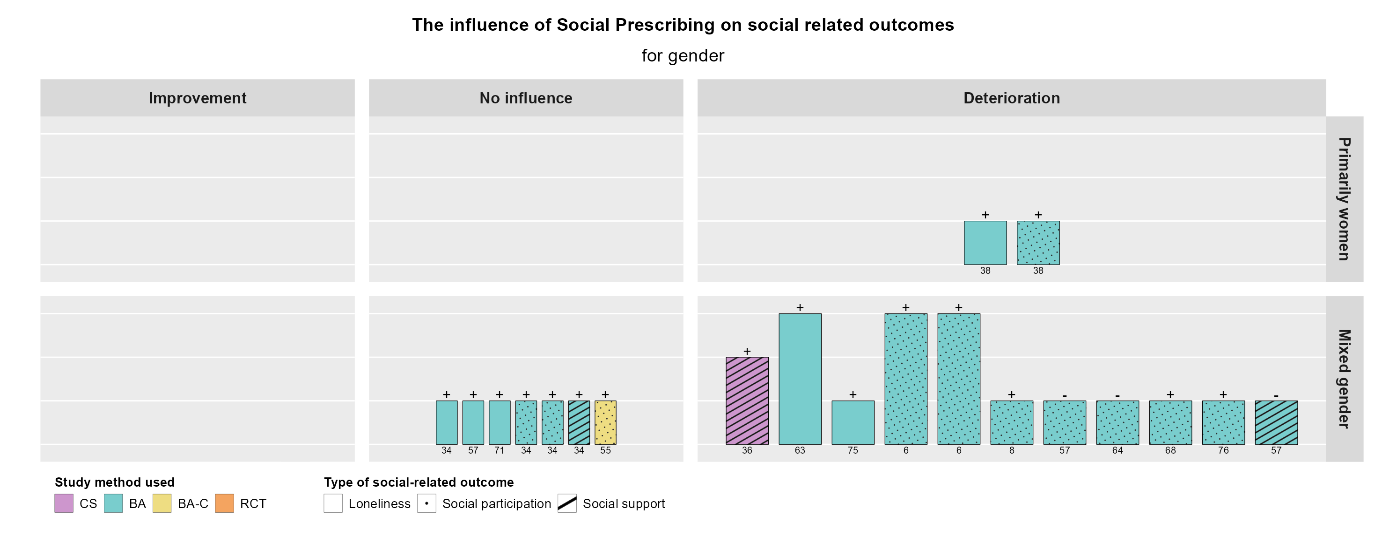

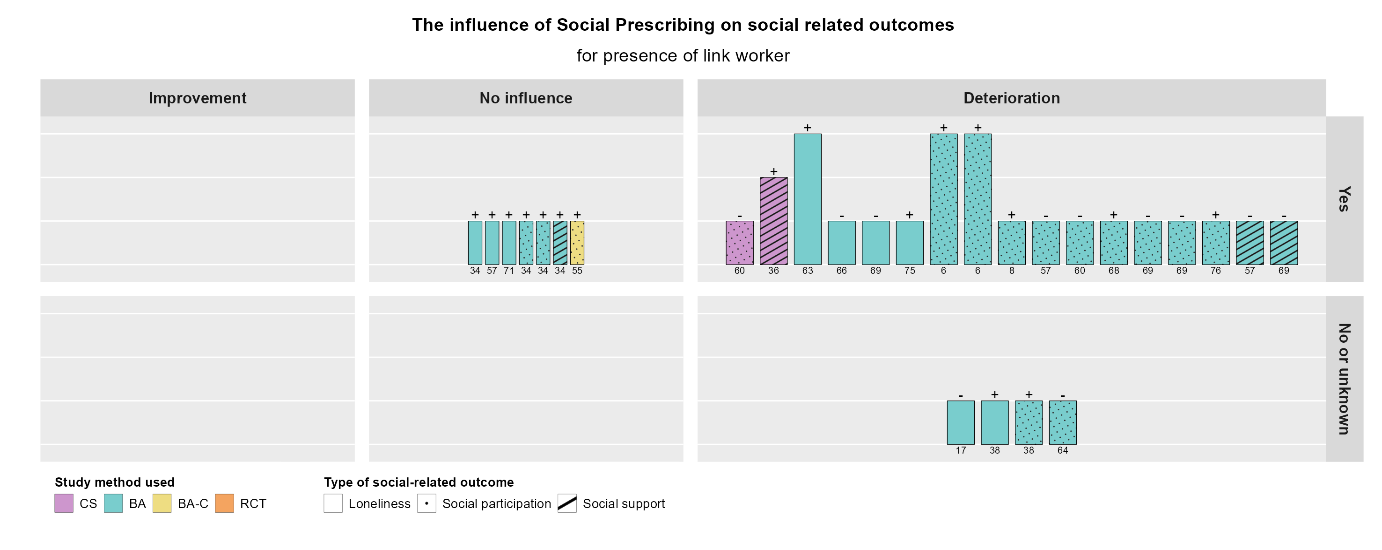

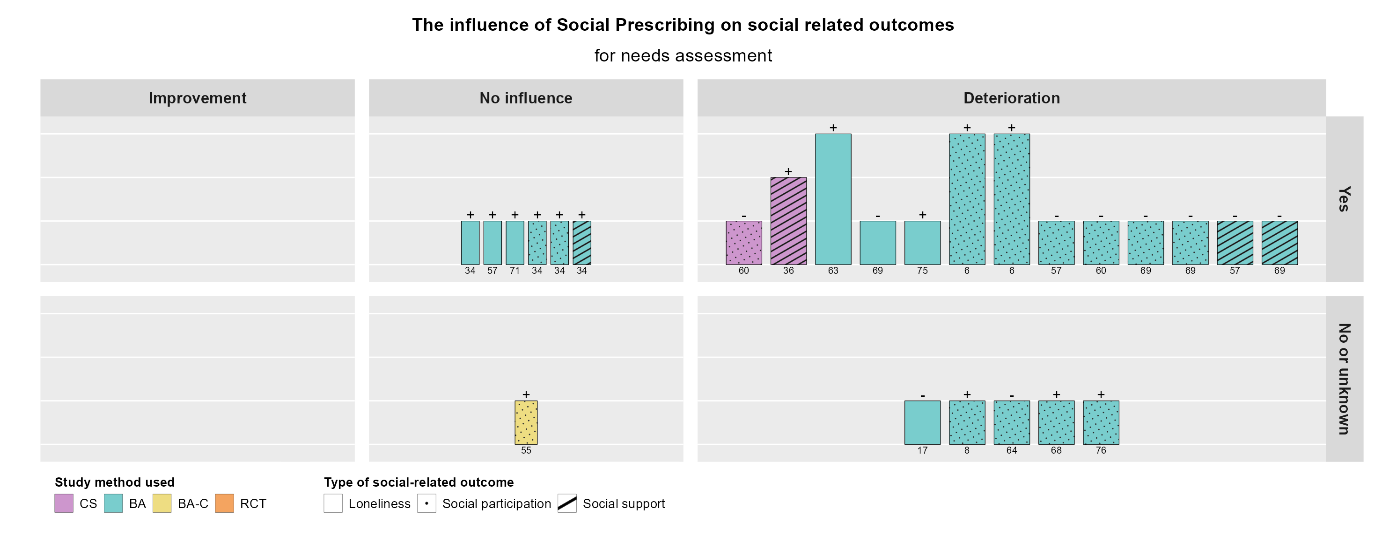

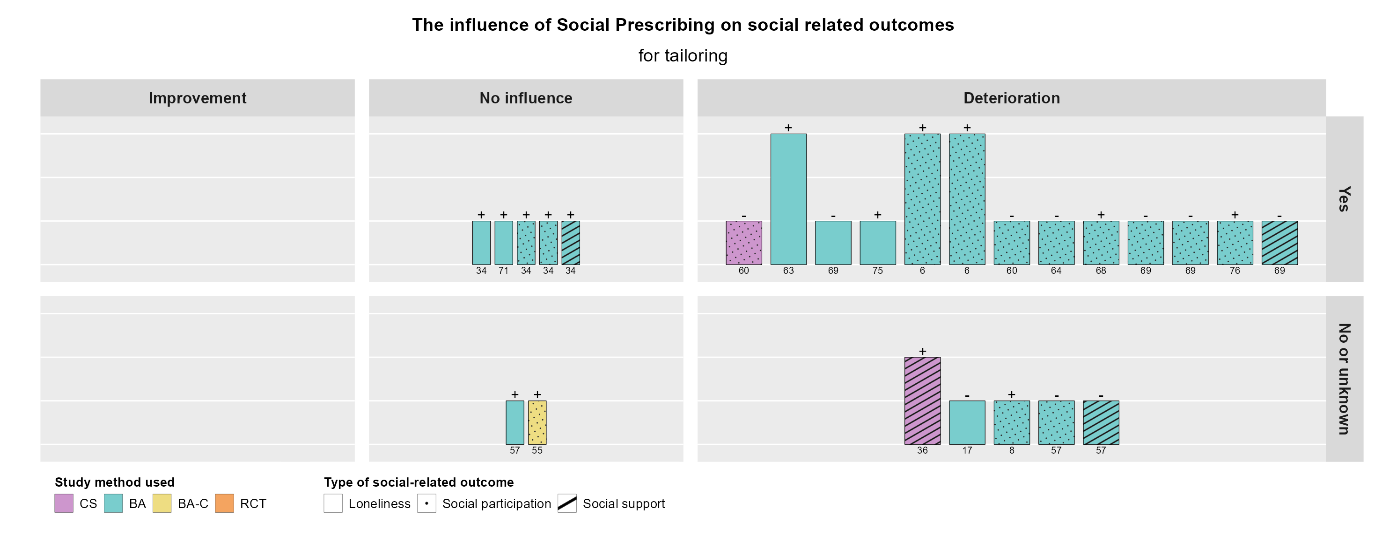

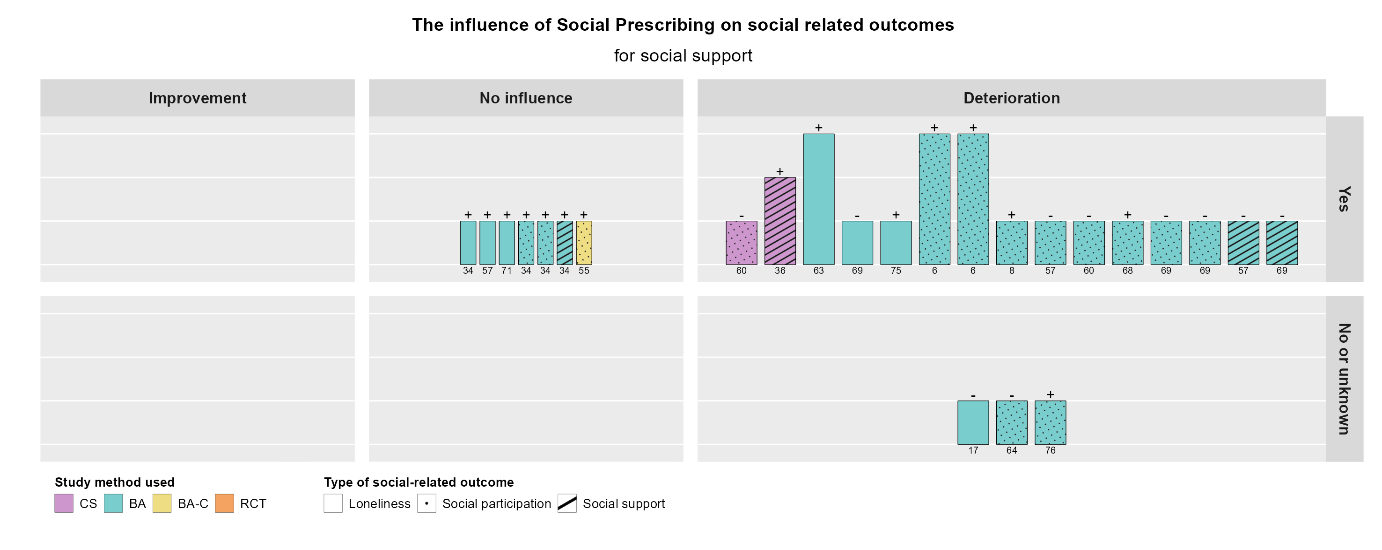
